# *NOTCH3* p.Arg1231Cys is Markedly Enriched in South Asians and Associated with Stroke

**DOI:** 10.1101/2023.10.05.23296511

**Authors:** Juan L. Rodriguez-Flores, Shareef Khalid, Neelroop Parikshak, Asif Rasheed, Bin Ye, Manav Kapoor, Joshua Backman, Farshid Sepehrband, Silvio Alessandro DiGioia, Sahar Gelfman, Tanima De, Nilanjana Banerjee, Deepika Sharma, Hector Martinez, Sofia Castaneda, David D’Ambrosio, Xingmin A. Zhang, Pengcheng Xun, Ellen Tsai, I-Chun Tsai, Regeneron Genetics Center, Maleeha Zaman Khan, Muhammad Jahanzaib, Muhammad Rehan Mian, Muhammad Bilal Liaqat, Khalid Mahmood, Tanvir Us Salam, Muhammad Hussain, Javed Iqbal, Faizan Aslam, Michael N. Cantor, Gannie Tzoneva, John Overton, Jonathan Marchini, Jeff Reid, Aris Baras, Niek Verweij, Luca A. Lotta, Giovanni Coppola, Katia Karalis, Aris Economides, Sergio Fazio, Wolfgang Liedtke, John Danesh, Ayeesha Kamal, Philippe Frossard, Thomas Coleman, Alan R. Shuldiner, Danish Saleheen

**Author notes:** **Correspondence**, Danish Saleheen, Alan Shuldiner.

## Abstract

The genetic factors of stroke in South Asians are largely unexplored. Exome-wide sequencing and association analysis (ExWAS) in 75 K Pakistanis identified NM_000435.3(NOTCH3):c.3691C>T, encoding the missense amino acid substitution p.Arg1231Cys, enriched in South Asians (alternate allele frequency = 0.58% compared to 0.019% in Western Europeans), and associated with subcortical hemorrhagic stroke [odds ratio (OR) = 3.39, 95% confidence interval (CI) = [2.26, 5.10], p value = 3.87×10^-9^), and all strokes (OR [CI] = 2.30 [1.77, 3.01], p value = 7.79×10^-10^). *NOTCH3* p.Arg231Cys was strongly associated with white matter hyperintensity on MRI in United Kingdom Biobank (UKB) participants (effect [95% CI] in SD units = 1.1 [0.61, 1.5], p value = 3.0×10^-6^). The variant is attributable for approximately 2.0% of hemorrhagic strokes and 1.1% of all strokes in South Asians. These findings highlight the value of diversity in genetic studies and have major implications for genomic medicine and therapeutic development in South Asian populations.

## Introduction

Pakistan, a country in South Asia, comprises over 231 million inhabitants. It is the fifth most populous country in the world with diverse ancestral backgrounds from South and Central Asia, West Asia, and Africa. Pakistan, and in general South Asia, represents an understudied region in large-scale genetic studies [1], thus providing an opportunity for novel discoveries of the genetic basis of diseases.

Stroke is a leading cause of death globally [2], and epidemiological studies suggest an elevated incidence and prevalence of stroke in Pakistan [2, 3] relative to Europe. The disparities in incidence and prevalence between Pakistan and Europe could be due to many factors, including difference in access to healthcare facilities with high-quality diagnostic capabilities and public health awareness and education. These disparities also may reflect differences in prevalence of risk factors such as hypertension and diabetes, lifestyle factors such as diet, physical activity and smoking, and genetic predispositions [4–7] . Studies of the genetic underpinnings of stroke in Pakistani populations have been limited, making this understudied population an opportune venue for stroke exome-wide sequencing and association studies (ExWAS).

At least 9 rare monogenic disorders are characterized by increased stroke risk, such as cerebral autosomal dominant arteriopathy with subcortical infarcts and leukoencephalopathy (CADASIL) due to mutations in *NOTCH3* [8]. CADASIL is distinct from other hereditary stroke diseases because it is characterized by vascular smooth muscle cell (VSMC) degeneration in small arteries and accumulation of protein aggregates known as granular osmophilic material (GOM) that contain aggregates of misfolded *NOTCH3* extracellular domain (ECD). The more common forms of stroke are likely polygenic with substantial contributions from behavioral and environmental factors as well as age. Major risk factors include high systolic blood pressure, high body mass index, hyperlipidemia, elevated glucose, and smoking [9]. Recent genome-wide association studies (GWAS) identified single nucleotide variants (SNVs) in more than 28 loci associated with stroke [10]. These variants are common non-coding variants with small effect sizes and were identified in predominantly European ancestry populations.

The aim of this study was to identify protein coding deleterious missense or loss-of-function (LoF) variants associated with stroke phenotypes in the Pakistani population. We performed exome sequencing in a 31 K discovery cohort consisting of 5,135 stroke cases and 26,602 controls of Pakistani origin. ExWAS identified NM_000435.3(NOTCH3):c.3691C>T, encoding the missense amino acid substitution p.Arg1231Cys, with an approximately three-fold increased risk of hemorrhagic stroke in heterozygotes. Follow-up meta-analysis of 61 K Pakistani (including an additional 160 cases and 30,239 controls) provided further support for association of *NOTCH3* p.Arg1231Cys with stroke (combined ischemic and hemorrhagic). This variant was present in approximately 1% of Pakistani and was markedly enriched with respect to Europeans in multiple South Asian (SAS) and West Asian (WAS) (also referred to as Greater Middle Eastern [11]) populations ranging from Turkey to India. The variant was estimated to explain up to 1.1% of strokes and 2.0% of hemorrhagic strokes in South Asia, a region having a population of > 2 billion people, thus having significant medical implications in these very large yet understudied populations and their global diaspora.

## Results

### ExWAS in the Pakistan Genomics Resource (PGR) discovery cohort identifies a markedly enriched missense variant in *NOTCH3* associated with stroke

Characteristics of the n = 5,135 stroke cases and n = 26,602 controls in the discovery cohort are summarized in Table 1. Compared to controls, stroke cases were modestly older and had a higher prevalence of known risk factors for vascular disease including hypertension, diabetes, myocardial infarction, and tobacco use (all p < 0.01). As expected, in this cohort ascertained for stroke, most cases were ischemic strokes and most hemorrhagic strokes were subcortical (Supplementary Figures 1 and 2, Supplementary Table 1).

**Table 1.** Baseline Characteristics of PGR Stroke Case-control Discovery Cohort^1^.

|  | Case<br>(n = 5,135) |  | Control<br>(n = 26,602) |  | P value | Case<br>p.Arg1231Cys<br>Carrier<br>(n=103)<sup>3</sup> |  | Case Non-Carrier<br>(n=4,998) |  | P value |
| --- | --- | --- | --- | --- | --- | --- | --- | --- | --- | --- |
|  | mean /<br>n | SD /<br>% | mean /<br>n | SD /<br>% |  | mean /<br>n | SD /<br>% | mean /<br>n | SD /<br>% |  |
| Female, n (%) | 2,277 | 44.3 | 9,330 | 35.1 | < 0.01 | 50 | 48.5 | 2,211 | 44.2 | 0.44 |
| Age at enrolment, years | 58.9 | 13.1 | 52.8 | 11.4 | < 0.01 | 58.7 | 12.1 | 58.9 | 13.2 | 0.87 |
| BMI, kg/m<sup>2</sup> | 25 | 3.9 | 27.5 | 4.4 | < 0.01 | 24.6 | 3.4 | 24.9 | 3.9 | 0.35 |
| Cholesterol mg/dL | 175.3 | 55.5 | 172.1 | 48.2 | < 0.01 | 174.8 | 49.1 | 175.3 | 55.7 | 0.94 |
| LDL-C mg/dL | 111.1 | 45.8 | 101.2 | 38.2 | < 0.01 | 110.8 | 41.1 | 111.1 | 45.9 | 0.94 |
| HDL-C mg/dL | 37.8 | 12.7 | 35 | 10.8 | < 0.01 | 38.8 | 12.1 | 37.8 | 12.7 | 0.43 |
| Triglyceride mg/dL | 140.3 | 81.8 | 185.7 | 120.1 | < 0.01 | 126.3 | 62.7 | 140.5 | 82.3 | 0.04 |
| Glucose, mg/dL | 148.7 | 75.5 | 143.4 | 84.9 | < 0.01 | 145.2 | 74.7 | 148.6 | 75.3 | 0.68 |
| HbA1c % | 6.9 | 1.8 | 6.6 | 2 | < 0.01 | 7.3 | 1.9 | 6.9 | 1.9 | 0.15 |
| Creatinine mg/dl | 1.2 | 0.8 | 0.9 | 0.5 | < 0.01 | 1.1 | 0.7 | 1.2 | 0.8 | 0.39 |
| Tobacco or other<br>stimulant user<sup>2</sup> | 1,850 | 36 | 9,242 | 34.7 | < 0.01 | 32 | 31.1 | 0 | 0 | 0.34 |
| **Comorbidities** |  |  |  |  |  |  |  |  |  |  |
| Hypertension , n (%) | 2,953 | 57.5 | 9,385 | 35.3 | < 0.01 | 57 | 55.3 | 2,879 | 57.6 | 1 |
| Diabetes, n (%) | 1,195 | 23.3 | 5,766 | 21.7 | < 0.01 | 29 | 28.2 | 1,157 | 23.1 | 0.29 |
| Myocardial infarction,<br>n (%) | 371 | 6.2 | 622 | 2.3 | < 0.01 | 5 | 4.9 | 215 | 4.3 | 0.98 |
| **Family history of** |  |  |  |  |  |  |  |  |  |  |
| Stroke, n (%) | 324 | 6.3 | 0 | 0 | < 0.01 | 14 | 13.6 | 306 | 6.1 | < 0.01 |
| Hypertension, n (%) | 573 | 11.2 | 4,299 | 16.2 | < 0.01 | 15 | 14.6 | 556 | 11.1 | 0.35 |
| Diabetes, n (%) | 349 | 6.8 | 5,199 | 19.5 | < 0.01 | 11 | 10.7 | 334 | 6.7 | 0.16 |
| Sudden death, n (%) | 150 | 2.9 | 569 | 2.1 | < 0.01 | 8 | 7.8 | 142 | 2.8 | < 0.01 |

- Shown are mean or total n and standard deviation or percentage for n = 5,135 cases versus n = 26,602 controls on the left and n = 103 case p.Arg1231Cys *NOTCH3* carriers versus n = 4,998 case non-carriers on the right. Comparison was conducted using R and p values are shown from chi-square test for categorical variables (Fisher's exact test if cell size was <5) and from T-test for continuous variables. - Tobacco or other stimulants include cigarettes, paan (chewed betel leaf and areca nut), naswar (snuff), gutka (chewing tobacco), huqqa (water pipe), chillum (hasish pipe). - Genotypes for the p.Arg1231Cys variant were not available for n=34.

Case:control ExWAS for all stroke cases and 4 stroke subtypes with sufficient case counts to provide statistical power (Supplementary Table 1) identified a genome-wide significant (p value < 5.0×10^-8^) association for NM_000435.3(NOTCH3):c.3691C>T (rs201680145), encoding the missense amino acid substitution p.Arg1231Cys, with subcortical hemorrhagic stroke (OR [95% CI] = 3.39 [2.26, 5.1], p value = 3.87×10^-9^; AAF = 0.58%) (Figure 1B, Supplementary Table 1 and Supplementary Figures 3 and, 4). The p.Arg1231Cys variant also showed evidence for association with all strokes combined (OR [95% CI] = 2.18 [1.65, 2.89], p value = 4.44×10^-8^) and other sub-categories of stroke (Supplementary Table 2). No other variants in the locus were associated with stroke (Figure 1C). We did not observe an association between p.Arg1231Cys and history of hypertension, elevated systolic or diastolic blood pressure, or smoking, known major risk factors for stroke (Supplementary Tables 3 and 4), and inclusion of these risk factors in regression analysis did not appreciably alter the effect of p.Arg1231Cys on stroke risk (Supplementary Table 5).

**Figure 1.**
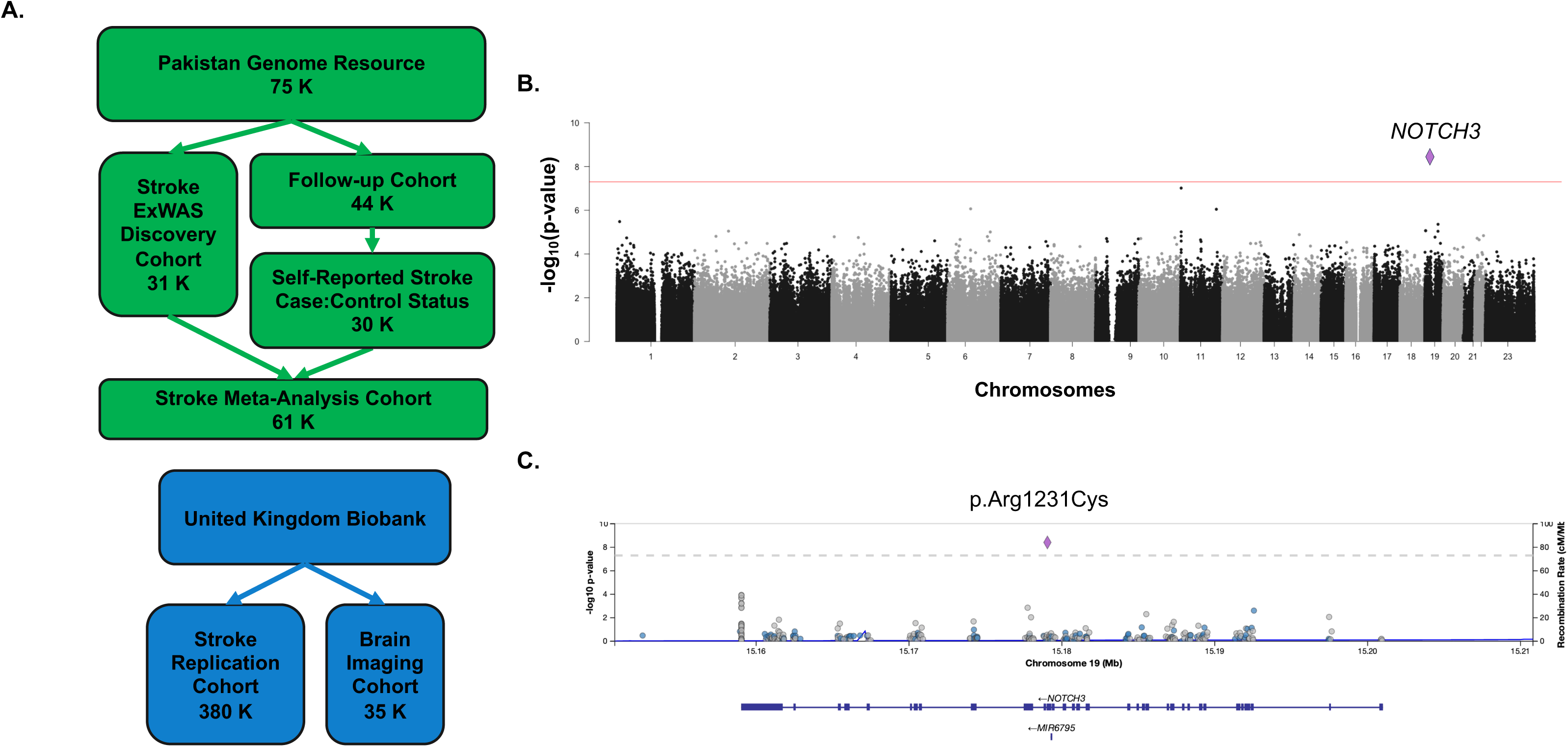
ExWAS Identifies *NOTCH3* p.Arg1231Cys Associated with Subcortical Hemorrhagic Stroke in Pakistan Genome Resource 31 K Discovery Cohort. A. Flow chart of the study described in this report. The discovery cohort consisted of a 31 K stroke case-control cohort (n = 31,737, including n = 5,135 stroke cases and n = 26,602 controls) from the Pakistan Genome Resource (PGR) (green boxes). A second PGR follow-up cohort of 44 K (n = 44,082) included 30 K participants with self-reported stroke case:control status for replication (n = 30,399, including n = 160 cases and n = 30,239 controls). UK Biobank data from 450 K sequenced participants was used for further analysis in a predominantly European ancestry population (blue boxes), 380 K of whom had stroke case:control status known (n = 9,143 cases and n = 371,403 controls), and 35 K of whom had brain MRI data (n = 35,344). **B.** Manhattan plot of subcortical hemorrhagic stroke ExWAS in 31 K PGR discovery cohort participants (n = 1,388 cases and n = 26,602 controls) with likelihood ratio test -log_10_ p-values of calculated using REGENIE (y-axis) across chromosomes (alternating grey and black dots) and variants (x-axis). A single variant (NC_000019.10:g.15179052G>A) on chromosome 19 predicting a missense variant p.Arg1231Cys in *NOTCH3* (pink diamond) exceeded the genome-wide significance threshold of 5×10^-8^ (red line). C. *NOTCH3* locus zoom plot of subcortical stroke ExWAS. The likelihood ratio test -log_10_ p values for variants tested are shown on the y-axis. The p.Arg1231Cys variant is labeled as a diamond. Other variants (circles) are colored based on linkage disequilibrium with the reference variant in 1000 Genomes [38]. Gene exon (thick line) and intron (thin line) model shown below the graph.

*NOTCH3* encodes Notch Receptor 3, a transmembrane signaling protein and part of an evolutionarily conserved family that plays a pleiotropic role in cell-cell interaction and neural development [12]. The extra-cellular domain (ECD) of *NOTCH3* consists of 34 Epidermal Growth Factor-like repeat (EGFr) domains [13], each containing 6 Cysteine (Cys) residues that form three disulfide bonds (Figure 2). Adding or removing Cys residues in the first 6 EGFr domains cause classical CADASIL, a highly penetrant rare autosomal dominant disease clinically characterized by migraine with aura, early-onset recurrent strokes, dementia, and behavioral changes [8]. The Cys-altering variant associated with stroke in this study, p.Arg1231Cys, occurs in the 31^st^ EGFr [14] and is predicted deleterious (Supplementary Table 6). Stroke cases heterozygous for p.Arg1231Cys and stroke cases without the variant were similar with respect to age, age of stroke onset, type of stroke, and stroke risk factors, suggesting a milder form of CADASIL not obviously clinically distinguishable from common forms of stroke in this population, although a detailed history for migraine or other manifestations of CADASIL were not available (Table 1).

**Figure 2.**
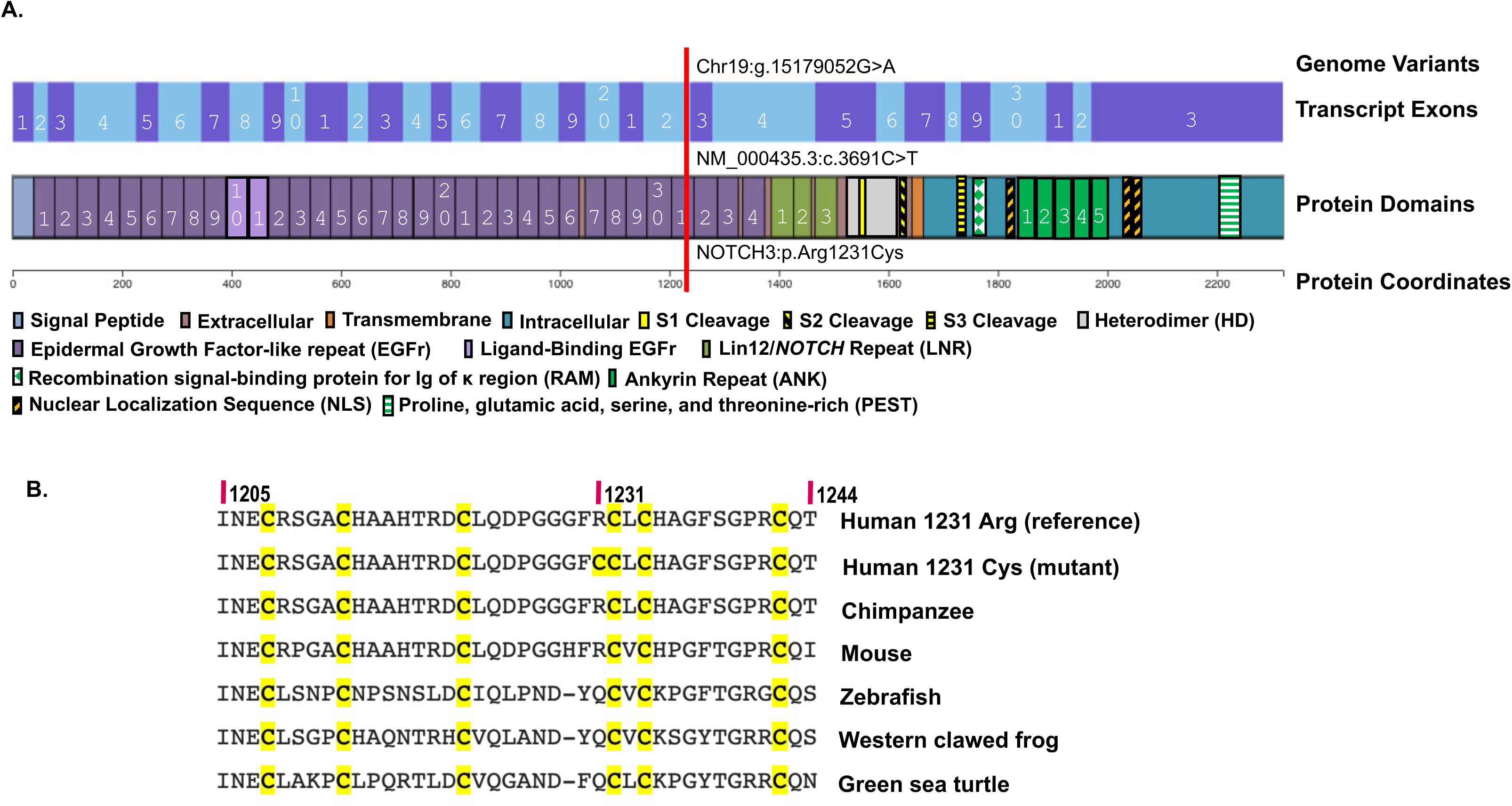
*NOTCH3* EGFr Domain Disruption by p.Arg1231Cys. Shown is *NOTCH3* p.Arg1231 in context of human *NOTCH3* protein domains and cross-species alignment of *NOTCH3* amino acid sequences. **A.** Human *NOTCH3* Protein Domains. Shown is the position of the associated variant in context of transcript exons (top, alternating blue and purple with numbering) and protein domains (bottom, color coded). *NOTCH3* can be divided into four major regions, from left-to-right the signal peptide (light blue), the extra-cellular domain (ECD, brown), the transmembrane domain (orange), and the intra-cellular domain (ICD, blue). The majority of the ECD is composed of 34 Epidermal Growth Factor-like repeat (EGFr) domains (in purple with white numbers). Domains involved in signaling are highlighted, including EGFr domains 10 to 11 involved in ligand binding (light purple, numbered), three cleavage domains (S1 in yellow, S2 in yellow with diagonal black stripes, S3 in yellow with black horizontal stripes), and three *Lin12*/*NOTCH* repeats (light green, numbered). The ICD contains the Recombination signal-binding protein for Ig of κ region (RAM) domain for transcription factor interaction (green and white checkers), the Nuclear Localization Sequences (NLS, orange with black stripes), five Ankyrin repeats involved in signal transduction (green with white numbers), and the Proline, glutamic acid, serine, and threonine-rich (PEST) domain essential for degradation (green with white stripes). The p.Arg1231Cys variant (red line top to bottom) removes a disulfide-bridge-forming cysteine in the 31^st^ EGFr domain of the ECD, coded by the 22^nd^ exon. **B.** Cross-species Alignment of *NOTCH3* (EGFr) Domain # 31 Amino Acid Sequences calculated using BLAST. Shown is an amino acid alignment of 31^st^ EGFr domain of *NOTCH3* (human sequence amino acids 1205 to 1244), including (top-to-bottom) human reference, human p.Arg1231Cys mutant, chimpanzee (*Pan troglodytes*), mouse (*Mus musculus*), zebrafish (*Danio rerio*), western clawed frog (*Xenopus tropicalis*), and green sea turtle (*Chelonia mvdas*), indicating conservation of the arginine (R) at position 1231 in mammals. Highly-conserved cysteine (C) residues (normally 6 per EGFr) are highlighted in yellow.

### Replication and Meta Analysis in PGR

An additional 30 K of whom self-reported stroke case:control status was obtained (160 cases and 30,239 controls) were sequenced by CNCD. Replication of the association was observed in this independent cohort (OR [95% CI] = 3.49 [1.56, 7.83],p value = 5.00×10^-3^. Meta-analysis for all strokes in the combined 61 K cohort achieved a genome-wide significant p value (OR [95% CI]) = 2.30 [1.76, 2.99], p value = 7.08×10^-10^) (Figure 3).

**Figure 3.**
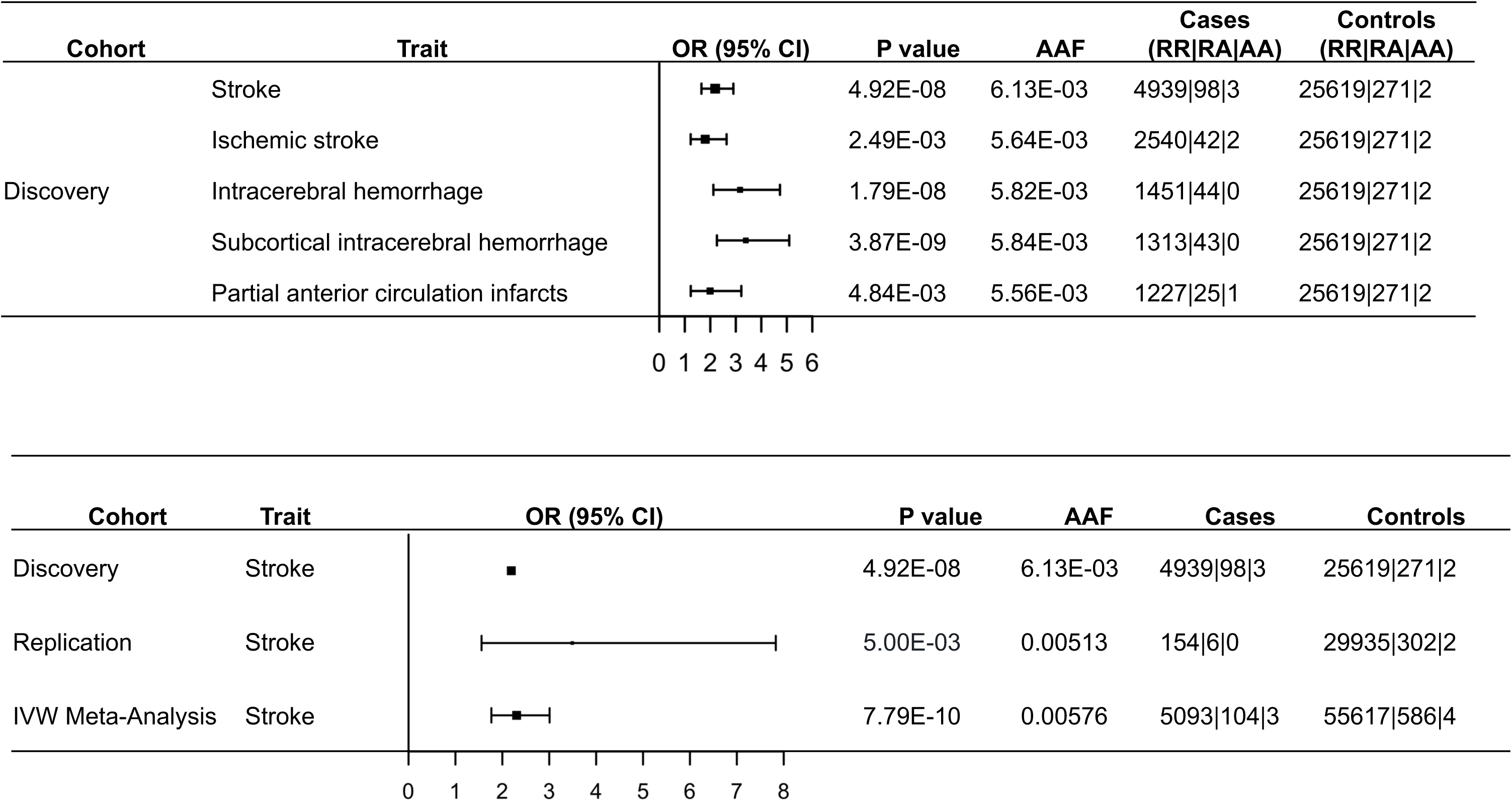
Forest Plot Showing Replication of *NOTCH3* p.Arg1231Cys Association with Stroke Across 61 K Pakistan Genome Resource Meta-Analysis. Shown is the cohort name, trait, odds ratio with 95% confidence interval, likelihood ratio test p value calculated using REGENIE\, alternate allele frequency, case count, and control count for five stroke phenotypes in the PGR 31 K discovery cohort (n = 5,135 stroke cases and n = 26,602 controls) (top), and Inverse Variance Weighted (IVW) Meta-Analysis using METAL of stroke in 61 K PGR Cohort, including 31 K PGR discovery cohort and 30 K PGR replication cohort (n = 160 cases and n = 30,239 controls) subset of the 44 K PGR follow-up cohort (bottom).

### Recall by Genotype

A total of 12 p.Arg1231Cys homozygotes from 9 nuclear families were identified in the PGR, including 9 discovery cohort probands and 3 follow-up cohort relatives identified through a callback of 128 call-back participants. Baseline characteristics of the 12 homozygotes are shown in Supplementary Table 7. Three of twelve (25%) homozygotes had a history of stroke; of note, all with stroke were >65 years of age while all without a history of stroke were <55 years of age.

Eight out of twelve (66%) p.Arg1231Cys homozygotes had a history of hypertension. Among the 128 callback participants, both systolic and diastolic blood pressure were trending higher (Supplementary Table 8). While there was no association with hypertension in the discovery cohort, p.Arg1231Cys homozygotes in the PGR had nominally higher diastolic blood pressure than heterozygotes or homozygous reference individuals (median = 95 mmHg, interquartile range 86 to 100; heterozygotes (median = 80 mmHg, interquartile range = 80 to 90), p value = 0.016 (Supplementary Table 9).

### Allele Frequency and Population Attributable Risk

The allele frequency of p.Arg1231Cys was 1.1% across the PGR 75 K, equivalent to a population prevalence of 1 in 46. After removing cases recruited for cardiovascular diseases (individuals enrolled at time of acute stroke, MI, and heart failure), the allele frequency of the variant was 0.51%, equivalent to a population prevalence of 1 in 98. This frequency was orders of magnitude higher relative to exomes of European ancestry from UK Biobank (AAF = 0.019%), corresponding to a population prevalence of 1 in 2,614). The variant was enriched (AAF > 0.1%) in other South Asian and West Asian populations [15] both within and outside of Pakistan (Supplementary Data, Supplementary Tables 10 and 11, Supplementary Figure 5).

We estimate that 2.0% [bootstrap 95% CI based on 10,000 resamples: 1.0% to 2.9%] of hemorrhagic strokes and 1.1% [bootstrap 95% CI based on 10,000 resamples: 0.6% to 1.6%] of all strokes in the Pakistani population are attributable to p.Arg1231Cys. Thus, this variant is a common cause of strokes in SAS and WAS populations, a finding that has implications for medical care as well as global health in these populations.

### Suggestive Associations at Other Loci

Although *NOTCH3* p.Arg1231Cys was the only variant associated with stroke at a genome-wide significant p value below 5.0×10^-8^, there were a total of 9 associations (5 loci) with p values below 1.0×10^-6^ and at least 10 variant carriers (Supplementary Table 12). In addition to *NOTCH3*, these included one known locus previously associated with stroke in a recent GWAS, lymphocyte specific protein *LSP1* [10].

The *LSP1* locus variant with suggestive association with intracerebral hemorrhagic stroke in this study was a common intronic variant (rs661348, OR [95% CI] = 1.3 [1.2, 1.4], p value = 8.0×10^-8^, AAF = 0.27). While rs661348 was not previously associated with stroke, two common non-coding variants in the *LSP1* locus were previously reported to be associated with stroke (rs569550 and rs1973765) [10]. Both variants were in linkage disequilibrium with rs661348 (r^2^ > 0.4 in 10 K unrelated PGR participants). A test of association for rs661348 conditional on these variants reduced the strength of the association for rs661348 to nominal (8.83×10^-4^) (Supplementary Table 13), suggesting that rs661348 represents the same known stroke risk locus. The *LSP1* locus was previously reported to be associated with hypertension [16]. In PGR there was a nominal association with hypertension (rs661348, OR [95% CI] = 1.0 [1.0, 1.1], p value = 2.61×10^-2^) (Supplementary Tables 14 and 15) also observed in UKB (OR [95% CI] = 1.0 [1.0, 1.1], p value = 8.05×10^-25^ (Supplementary Tables 15 and 16).

### *NOTCH3* p.Arg1231Cys is associated with stroke and CADASIL-like phenotypes in UK Biobank

To investigate stroke and CADASIL-related phenotypes in an independent cohort, UK Biobank data was reviewed for associations with *NOTCH3* p.Arg1231Cys. A total of 255 heterozygotes for *NOTCH3* p.Arg1231Cys were observed in 450 K exome-sequenced individuals from this predominantly European cohort, with a markedly lower allele frequency (AAF = 0.019%) (Supplementary Table 18). Phenome-wide association (PheWAS) of 10,168 phenotypes revealed nominally significant association of p.Arg1231Cys with ischemic stroke (OR [95%CI] = 4.0 [1.9, 8.6]), p value = 4.1×10^-4^), all strokes combined (OR [95% CI] = 1.9 [1.1, 3.5], p value = 0.031), hypertension (ICD 10 code I10) (OR [95% CI] = 1.5 [1.1, 2.2], p value = 0.019), and recurrent major depression (OR [95% CI] = 3.2 [1.5, 6.8], p value = 0.0031) (Supplementary Table 19). No association was observed for hemorrhagic stroke, migraine, dementia, mood changes, Alzheimer’s disease, or urinary incontinence. The lack of association (p value = 0.066) with hemorrhagic stroke in UKB Europeans was likely due to low statistical power, given the lower variant allele frequency and lower hemorrhagic stroke prevalence in UKB compared to PGR. Nonetheless, the odds ratio (OR [95% CI] = 5.8 [0.88, 39.1]) was high.

In addition to recurrent strokes, brain white matter loss is a major and early phenotype characteristic of CADASIL that is focused on particular brain regions [8, 14]. *NOTCH3* p.Arg1231Cys was strongly associated with a cluster of brain MRI quantitative phenotypes, e.g., total volume of white matter hyperintensities (WMH) from T1 and T2 FLAIR images (effect [95% CI] in SD units = 1.1 [0.61, 1.5], p value = 3.0×10^-6^) with carriers having 7.4 cm^3^ more WMH volume than controls (Supplementary Figure 6 and Supplementary Table 20). The most prominent alterations in WMH in p.Arg1231Cys carriers were observed in the centrum semiovale and periventricular white matter (Supplementary Figure 7). Taken together, these results demonstrate *NOTCH3* p.Arg1231Cys carriers have increased risk of established markers of small vessel disease and clinical phenotypes observed in CADASIL [8].

### Pathogenic burden of all Cys-altering variants within *NOTCH3* EGFr domains specifically associated with CADASIL phenotypes in UK Biobank

Burden test analysis allows for increased statistical power to detect association by combining signal across multiple rare variants. Prior studies have shown that pathogenic variants in CADASIL are limited to variants that add or remove a Cysteine (Cys-altering) in *NOTCH3* EGFr domains normally containing 6 Cysteines. Furthermore, patients with Cys-altering variants in the first 6 EGFr domains have more severe symptoms than in EGFr domains 7 to 34 [14, 17, 18], including larger regions of brain white matter loss [19], more granular osmophilic material (GOM) aggregates in blood vessels [19], and worse prognosis [14].

In order to test these hypotheses, a set of custom gene burden tests were designed and compared to single variant test results for *NOTCH3* p.Arg1231Cys. In UKB, 758 individuals carried one of 98 unique Cys-altering variants across the 34 EGFr domains in *NOTCH3* (Supplementary Tables 21 and 22). A burden test aggregating all UKB EGFr domain Cys-altering variants into a single statistical test was strongly associated with stroke (OR [95% CI] = 2.86 [2.14, 3.82], p value = 6.29×10^-10^; AAF = 0.01%) (Table 2). In contrast to Cys-altering variants within EGFr domains, Cys-altering variants outside of EGFr domains were not associated with stroke (OR [95% CI] = 0.97 [0.46, 2.03], p value = 9.3×10^-1^; AAF =0.039%) (Table 2). In order to rule out the possibility that any missense variants in EGFr domains are associated with stroke, a test limited to the most commonly altered (added or removed) amino acid in *NOTCH3*, serine (Ser), was tested and did not show any evidence of association with stroke (OR [95% CI] = 0.98 [(0.8, 1.2], p value = 0.084; AAF = 0.54%] (Table 2, Supplementary Table 23). Interestingly, a burden test limited only to predicted loss of function (pLoF) variants (frameshift, splice variant, stop gain) did not show significant evidence for association with stroke (OR [95% CI] = 1.38 [0.50, 3.85], p value = 0.54; AAF = 0.019%) (Table 2, Supplementary Table 24). These results provide evidence to support the hypothesis that EGFr domain Cys-altering variants within *NOTCH3* are associated with stroke, in contrast to other protein-altering variants.

**Table 2.** UKB Ischemic Stroke Association Across *NOTCH3* Variant Classes and Domains^1^.

| *NOTCH3*<br>Variant Class | Effect<br>[OR (95%CI)] | P value | AAF<br>(%) | Cases<br>(RR\|RA\|AA) | Controls<br>(RR\|RA\|AA) |
| --- | --- | --- | --- | --- | --- |
| p.Arg1231Cys | 3.38 (1.65,6.94) | 8.8x10<sup>-4</sup> | 0.02 | 9124\|11\|0 | 370986\|139\|0 |
| EGFr 1-34 Cys-altering | 2.86 (2.14,3.82) | 6.3x10<sup>-10</sup> | 0.01 | 9094\|49\|0 | 370693\|709\|1 |
| EGFr 1-6 Cys-altering | 29.51 (10.39,83.82) | 1.4x10<sup>-7</sup> | 0.002 | 9137\|6\|0 | 371394\|9\|0 |
| EGFr 7-34 Cys-altering | 2.55 (1.87,3.46) | 1.6x10<sup>-7</sup> | 0.098 | 9100\|43\|0 | 370702\|700\|1 |
| Non-EGFr Cys-altering | 0.97 (0.46,2.03) | 9.3x10<sup>-1</sup> | 0.039 | 9136\|7\|0 | 371111\|292\|0 |
| EGFr 1-34 Ser-altering | 0.98 (0.8,1.2) | 8.4x10<sup>-1</sup> | 0.54 | 9046\|97\|0 | 367355\|4042\|6 |
| EGFr 1-6 Ser-altering | 0.76 (0.23,2.56) | 6.6x10<sup>-1</sup> | 0.018 | 9141\|2\|0 | 371271\|132\|0 |
| EGFr 7-34 Ser-altering | 0.99 (0.8,1.21) | 8.9x10<sup>-1</sup> | 0.53 | 9048\|95\|0 | 367486\|3911\|6 |
| Non-EGFr Ser-altering | 0.84 (0.52,1.34) | 4.5x10<sup>-1</sup> | 0.10 | 9128\|15\|0 | 370656\|744\|3 |
| LoF variants | 1.38 (0.50,3.85) | 5.4x10<sup>-1</sup> | 0.019 | 9138\|5\|0 | 371261\|142\|0 |
| LoF + any missense | 1.09 (1.01,1.19) | 3.3x10<sup>-2</sup> | 3.30 | 8490\|652\|1 | 346648\|24710\|45 |

- <sup>1</sup>. Burden test with age, age<sup>2</sup>, gender, 10 PCs as covariates was conducted using REGENIE to compare the association signal and effect size across variant classes and domains for ischemic stroke in n = 9,143 cases and n = 371,403 controls from UK Biobank. *NOTCH3* variant classes include the single variant association (p.Arg1231Cys) for reference at the top and burden tests limited to: Cys-altering variants in EGFr domains 1 to 34; Cys-altering variants in EGFr domains 1 to 6; Cys-altering variants in EGFr domains 7 to 34; Cys-altering variants outside of EGFr domains; Ser-altering variants in EGFr domains 1 to 6; Ser-altering variants in EGFr domains 7 to 34; Ser-altering variants outside of EGFr domains; Loss of function (LoF) variants (defined as stop-gain, stop-loss, frameshift and splice-site variants with AAF <1%); and LoF and any missense variants (AAF < 1%). Abbreviations: EGFr, epidermal growth factor-like repeat domain; AAF, alternate allele frequency; RR, reference allele homozygote; RA, reference/alternate allele heterozygote; AA, alternate allele homozygote; 95% CI, 95% confidence interval

While hemorrhagic stroke represents a small proportion of the strokes reported in the UKB, the set of Cys-altering variants were also tested for association with hemorrhagic stroke. A nominal association with hemorrhagic stroke (OR [95% CI] = 3.61 [1.39, 9.34], p value = 8.31×10^-3^; AAF 0.025%) was observed, despite low statistical power.

Consistent with stroke risk, in MRI data of 35,344 UKB individuals, Cys-altering variants in *NOTCH3* EGFr domains were strongly associated with WMH volume (p value = 3.7×10^-13^; with carriers having 5.4 cm^3^ greater WMH volume than controls). These WMH differences were strongest in the centrum semiovale and periventricular white matter (Supplementary Figure 7). Additionally, we found strong WMH signal in the external capsule, which is known to be involved in CADASIL. We found weaker evidence for association of *NOTCH3* LoF variants with WMH (effect size [95% CI] = 6.8 cm^3^ [4.2 cm^3^, 10.9 cm^3^], p-value = 1.68×10^-4^).

Prior studies have binned *NOTCH3* EGFr domain Cys-altering variants in up to three distinct severity or risk groups based on EGFr domain number [10, 14, 17]. Indeed, we observed a much larger effect size for Cys-altering variants in high-risk EGFr domains 1-6 (OR [95% CI] = 29.5 [10.4, 83.8], p value = 1.37×10^-7^; AAF = 0.002%] compared to Cys-altering variants in EGFr domains 7-34 (OR [95% CI] = 2.55 [1.87, 3.46], p value = 1.59×10^-7^; AAF = 0.098%) (Table 2,

Supplementary Table 23). These results are consistent with prior reports of differences in stroke risk between EGFr domain risk groups not correlated with differences in signaling activity between EGFr risk groups [17].

## Discussion

This report describes the largest ExWAS of stroke conducted thus far in a South Asian population and highlights a Cys-altering missense variant in the 31^st^ EGFr domain of *NOTCH3* associated with stroke at a genome-wide level of statistical significance. This is the first study to report a genome-wide-significant association between *NOTCH3* and stroke, a discovery enabled because *NOTCH3* p.Arg1231Cys is markedly enriched in Pakistanis compared to Western European and non-Eurasian populations. Harbored in ∼1 percent of Pakistani, p.Arg1231Cys is associated with a ∼3-fold increased risk of hemorrhagic stroke. While some regional variability in the allele frequency is observed, p.Arg1231Cys is enriched in populations ranging from Turkey in West Asia to India in South Asia, suggesting a substantial contribution to stroke risk in millions of individuals across South Asia and West Asia as well as their global diaspora.

*NOTCH3* was not previously associated with stroke in the largest GWAS predominantly consisting of European-derived participants [10]. In contrast to prior studies, both the discovery and replication cohorts in this study were South Asian, hence avoiding the bias encountered in studies with a European discovery cohort. Given the much lower allele frequency of p.Arg1231Cys in European populations, we observed a nominal association between p.Arg1231Cys and stroke in the UK Biobank study, showing a similar effect size as in South Asians. Nominal associations were also observed for phenotypes related to CADASIL, such as hypertension and depression. While brain images were not available for the Pakistani cohort, a strong association was observed between p.Arg1231Cys and quantitative brain MRI phenotypes in UKB data, such as white matter hyperintensity.

Cys-altering mutations in proximal EGFr domains of *NOTCH3* are known to cause autosomal dominant CADASIL, a rare highly penetrant distinct syndrome that includes early onset recurrent subcortical strokes. In contrast to classical CADASIL pathogenic variants, p.Arg1231Cys is in the 31^st^ of 34 EGFr domains, appears to have more moderate penetrance, and is not obviously clinically distinguishable from more common multi-factorial forms of stroke in South Asians. The p.Arg1231Cys *NOTCH3* variant is currently classified in ClinVar and recent reviews [14] as a variant of uncertain significance [20] or “low risk” [17]; however, based on our current findings, there is strong genetic, computational, and imaging evidence of pathogenicity for this variant despite reduced penetrance and severity compared to “classic” Cys-altering CADASIL pathogenic variants in EGFr domains 1 to 6 [14, 19].

Prior studies have debated if the mechanism whereby Cys-altering variants contribute to CADASIL-related pathology is through toxic aggregate gain of function (GoF) or a loss of normal signaling function (LoF). One study demonstrated excess risk of CADASIL-related phenotypes for Cys-altering variants in EGFr domains 1 to 34 [18], while other studies showed greater risk in EGFr domains 1 to 6 relative to EGFr domains 7 to 34 [14, 19]. A recent study showed evidence for expanding the high-risk tier of EGFr domains to include domains 8, 11, and 26 [17]. The current study provides three additional refinements. First, this study is the first to assess risk for LoF variants, and did not observe significant association signal (**Table 2**), although the number of LoF carriers was small and thus power is limited to detect such associations. These findings suggest that pathological mechanisms driven by dysfunctional disulfide bridge formation and subsequent protein misfolding and aggregation, as is commonly observed in CADASIL, may be more pathologic than simple LoF (haploinsufficiency) [19].

Second, we demonstrate association between p.Arg1231Cys with stroke, thus demonstrating that CADASIL-related stroke is not uncommon as was previously thought. While prior studies have shown enrichment of p.Arg1231Cys in South Asians [21], and have used this information as evidence to classify p.Arg1231Cys variant as “low-risk” [17], the current study provides evidence contrary to that verdict. Furthermore, we have demonstrated a broader enrichment of the variant across the region, including multiple West Asian and South Asian populations. Third, the prior studies demonstrated a brain-wide association with WMH, while the current study identifies pathology focused in the external capsule and other brain regions known for CADASIL pathology.

CADASIL is characterized by both ischemic and hemorrhagic strokes, although the factors that contribute to the manifestation of one versus the other stroke type awaits further clarification [8]. Hemorrhagic stroke appears to represent a larger proportion of strokes in South Asia than in Europe [2]. In this study, the p.Arg1231Cys association signal was stronger in PGR for hemorrhagic strokes than for ischemic strokes, despite nearly two-fold larger ischemic stroke case counts. In contrast, the UKB association signal appeared stronger for ischemic stroke, possibly due to low hemorrhagic stroke case count and thus statistical power in this cohort.

Statistical power issues aside, differences in manifestation of p.Arg1231Cys in South Asians compared to Europeans may be attributable to differences in risk factors such as age, blood pressure, diabetes, air pollution, smoking, medications such as anti-platelet and anti-coagulants used to manage atherosclerotic disease, genetic background, or study-specific differences in criteria to categorize stroke sub-types. Further research is needed to better ascertain the mechanism behind cerebral arterial wall pathobiology and clinical presentation of ischemic versus hemorrhagic stroke in p.Arg1231Cys carriers.

Currently there are no known effective preventive or therapeutic interventions for CADASIL or less penetrant forms of *NOTCH3* related stroke. However, our analyses provide clues toward their development. First, in contrast to our analyses of EGFr domain Cys-altering missense variants in *NOTCH3* that were significantly associated with stroke, predicted loss of function variants that would be expected to not produce a functional protein were not significantly associated with stroke. These observations suggest that targeting therapeutic interventions that decrease expression of mutant protein (such as siRNA, antisense oligonucleotides, and CRISPR), induce exon skipping of altered EGFr domains [22, 23], or accelerate removal of GOM may prove beneficial for prevention and/or treatment [24].

Our analyses also suggest that p.Arg1231Cys is modestly associated with hypertension, although p.Arg1231Cys association with stroke risk appears independent of hypertension or other stroke risk factors such as smoking, age and sex. Animal models of CADASIL show decreased vascular tone and contractility, most likely driven by loss of physiologic function and subsequent degeneration of vascular smooth muscle cells (VSMCs) [25]. These observations suggest that while management of hypertension and smoking cessation are effective modalities for primary and secondary prevention of stroke, those with *NOTCH3* mutation related strokes will need additional therapeutic interventions, as existing hypertensive medications cannot restore VSMC function.

A limitation of this study is the lack of brain imaging analysis for the Pakistani carriers, such that specific brain regions affected by the ischemic and hemorrhagic strokes could be ascertained and compared. In addition, we lacked more detailed clinical data such as presence of migraines and longitudinal data of disease course including stroke recurrence, dementia, and depression.

Further characterization of p.Arg1231Cys carriers will be necessary to obtain better estimates of penetrance as well as to identify distinguishing clinical or biomarker characteristics that may have utility in early diagnosis, prevention and treatment, and for recommendations for cascade screening in family members. Migraine symptoms typically precede stroke by 10+ years in CADASIL patients [8], thus the combination of migraine with aura, depression and family history of stroke could be sufficient evidence to prescribe *NOTCH3* genetic testing. Finally, the effect of LoFs on stroke risk will require larger sample sizes for more definitive comparison to stroke risk of Cys-altering variants.

In conclusion, we identified a highly enriched Cys-altering variant in *NOTCH3* in South Asians that expands the phenotypic spectrum of CADASIL from rare and highly penetrant to common and moderately penetrant. Based on our estimates, this single variant may be responsible for ∼1.1% of all strokes combined and ∼2.0% of hemorrhagic strokes in South Asians. Among 1.9 billion South Asians there could be over 26 million carriers for the variant. Thus, this work has important implications for genetic screening and early identification of at-risk individuals, and the future opportunity for rationally targeted therapeutic interventions.

## Methods

### Summary

Details of methods below. Briefly, 75 K individuals were recruited and consented in Pakistan for whole exome sequencing, including a stroke case:control discovery cohort of 31 K (including n = 5,135 cases and n = 26,602 controls) sequenced by the Regeneron Genetics Center and a follow-up cohort of 44 K (n=44,082), including 30 K with self-report stroke case:control status used for replication and meta-analysis (n = 160 cases and n = 30,239 controls). ExWAS was conducted for stroke and 4 overlapping stroke subtypes (intracerebral hemorrhage, subcortical intracerebral hemorrhage, ischemic stroke, and partial anterior circulating infarct) in the discovery cohort and combined in a meta-analysis of stroke with the replication cohort using both single-variant and gene burden test models [26]. Population attributable fraction of stroke for associated variants was calculated as based on the prevalence of mutation among cases and odds ratio (OR) for risk of stroke in the discovery cohort using the standard definition [27].

Consented callbacks were conducted in n = 128 individuals within families of homozygotes for associated variants. For comparison and validation, analyses were conducted in UK Biobank data using publicly available datasets and methodologies [28–31], including association analysis of p.Arg1231Cys *NOTCH3* with stroke phenotypes in 380 K participants, and brain imaging phenotypes in 35 K participants (Figure 1A).

### Study Populations

This study focused on two distinct cohorts, including 75 thousand (K) individuals from the Pakistan Genomic Resource (PGR) and 370 K individuals from the United Kingdom Biobank (UKB) The PGR 75 K individuals were recruited and consented in Pakistan for whole exome sequencing (WES) (n = 75,819), including a stroke case:control discovery cohort of 31 K (n = 31,737, including n = 5,135 cases and n = 26,602 controls) sequenced by the Regeneron Genetics Center and a follow-up cohort of 44 K (n = 44,082), including 30 K with self-report stroke case:control status used for replication and meta-analysis (n = 30,399, including n = 160 cases and n = 30,239 controls). The remaining n = 13,684 in the follow-up cohort had sequence data but not stroke case:control status known, including n = 6,067 produced by WES and n = 7,616 produced by whole genome sequencing (WGS). (Figure 1A).

### Pakistan Genomic Resource (PGR)

PGR is a growing biobank that aims to enroll 1 million participants across Pakistan and as of September 2023, ∼250,000 participants across 48 clinical sites in 17 cities from all over Pakistan have been enrolled. Following the success of a case-control study design in genetic studies adopted by several international (e.g., Wellcome Trust case-control consortium) and regional studies (e.g., PROMIS), PGR is a national consortium of several case-control studies focused on 50 distinct phenotypes, including: stroke, myocardial infarction, angiographically confirmed coronary artery disease, heart failure, age-related macular degeneration, keratoconus, diabetic retinopathy, glaucoma, asthma, chronic obstructive pulmonary disease (COPD), non-alcoholic fatty liver disease (NAFLD), type-2 diabetes, chronic kidney disease, Alzheimer’s disease, Parkinson’s disease, dementia, progressive multiple sclerosis, autism, Huntington’s disease, hematological cancers, breast cancer, ovarian cancer, cancers of head and neck, esophageal cancer, lung cancer, gastric cancer, colorectal cancer, melanoma, cancers of the urinary tract, cervical cancer, prostate cancer, rheumatoid arthritis, systemic lupus erythematosus (SLE), psoriatic arthritis, ankylosing spondylitis, osteoarthritis, scleroderma, juvenile arthritis, systemic sclerosis, inflammatory myositis, alopecia areata, acne rosacea, primary Sjogren’s syndrome, sarcoidosis, idiopathic pulmonary cholangitis, idiopathic pulmonary fibrosis, vitiligo, longevity / healthy aging, and previously uncharacterized Mendelian disorder. For each of these phenotypes, screening is carried out at specialized clinical sites across Pakistan by trained research medical officers who review inclusion and exclusion criteria and approach eligible participants for recruitment into PGR. In a similar manner, for each of the phenotypes, controls are frequency-matched to cases on sex and age (in 5-year bands). Controls are being recruited in the following order of priority: (1) visitors of patients attending the out-patient department; (2) patients attending the out-patient department for routine non-phenotype related complaints, or (3) non-blood related visitors. Following informed consent, both cases and controls are enrolled.

Research medical officers administer pre-piloted epidemiological questionnaires to participants that seek a total of >200 items of information in relation to: ethnicity (e.g, personal and paternal ethnicity, spoken language, place of birth and any known consanguinity); demographic characteristics; lifestyle factors (e.g., tobacco and alcohol consumption, dietary intake and physical activity); and personal and family history of disease; and medication usage. The Center for Non-Communicable Diseases (CNCD), Pakistan serves as the sponsor and the coordinating center of PGR.

Using standardized procedures and equipment, research officers obtain measurements of height, weight, waist and hip circumference, systolic and diastolic blood pressure, and heart rate. Waist circumference is assessed over the abdomen at the widest diameter between the costal margin and the iliac crest, and hip circumference is assessed at the level of the greater trochanters.

Information extracted from questionnaires and physical measurements is entered by two different operators into the central database, which is securely held at CNCD, Karachi, Pakistan. Non-fasting blood samples (with the time since last meal recorded) are drawn by phlebotomists from each participant and centrifuged within 45 min of venipuncture. A total of 29 ml of blood is drawn from each participant in 2 × 6 ml serum tubes and 3 × 5 ml EDTA tubes. Hence, a total of five blood tubes are collected per participant, including serum, EDTA plasma and whole blood which are all stored in cryogenic vials. All samples are stored temporarily at each recruitment center at −20°C. Serum, plasma and whole blood samples are transported daily to the central laboratory at CNCD where they are stored at −80°C. The long-term −80°C sample repository.

Measurements of total cholesterol, high-density lipoprotein-cholesterol, triglycerides, AST, ALT, glucose, creatinine, and HbA1c (in a subset) are performed centrally using (Roche Diagnostics GmbH, USA) in all study participants.

Research technicians trained in accordance with standard operating procedures in laboratories at CNCD extracted DNA from leukocytes using a reference phenol-chloroform protocol. DNA concentrations are determined. The yield of DNA per participant is typically between 600 and 800 ng/μl in a total volume of about 500 μl. To minimize any systematic biases arising from plate- or batch-specific genotyping error and/or nonrandom missingness, stock plates are used to generate genotyping plates which contain a mixture of cases and controls along with negative and positive controls designed to address genotyping quality control (QC), plate identification and orientation.

PGR has received approval by the relevant research ethics committee of each of the institutions involved in participant recruitment, as well as centrally by the IRB board of the Center for Non-Communicable Diseases which is registered with the National Institutes of Health, USA. PGR has also been approved by the National Bioethics Committee, Islamabad Health Research Institute, National Institutes of Health of Pakistan.

Eligibility criteria was defined as described below. Ischemic stroke sub-types in the PGR cohort were defined using TOAST [32] and Oxfordshire [33] clinical criteria.

### UK Biobank

The UK Biobank (UKB) cohort had detailed medical records and lifestyle data as described online in the UKB Showcase (https://biobank.ndph.ox.ac.uk/showcase/) [28]. Stroke case:control status was available in 380 K UK Biobank participants, of which 280 K also had smoking and hypertension status (referred to as UKB replication cohort). A sub-cohort of n = 35,977 UKB participants had brain MRI data which was produced and analyzed as described online (https://biobank.ctsu.ox.ac.uk/crystal/crystal/docs/brain_mri.pdf) [29–31]. MRI images for n = 19 p.Arg1231Cys carriers were re-analyzed in order to identify brain regions affected (referred to as UKB 35 K brain imaging cohort). The description of phenotypes and methods for normalizing the data, including rank-inverse normal transformation (RINT) are described online [28].

### Exome Sample Preparation, Sequencing, and QC

Genomic DNA samples were transferred to the Regeneron Genetics Center from the CNCD and stored in an automated sample biobank at –80 °C before sample preparation. DNA libraries were created by enzymatically shearing DNA to a mean fragment size of 200 bp, and a common Y-shaped adapter was ligated to all DNA libraries. Unique, asymmetric 10 bp barcodes were added to the DNA fragment during library amplification to facilitate multiplexed exome capture and sequencing. Equal amounts of sample were pooled before overnight exome capture, with a slightly modified version of IDT’s xGenv1 probe library; all samples were captured on the same lot of oligonucleotides. The captured DNA was PCR amplified and quantified by quantitative PCR. The multiplexed samples were pooled and then sequenced using 75 bp paired-end reads with two 10 bp index reads on an Illumina NovaSeq 6000 platform on S4 flow cells. A total of n = 42,695 samples were made available for processing. We were unable to process n = 1,948 samples, most of which failed QC during processing owing to low or no DNA being present.

A total of n = 40,747 samples were sequenced, of which n = 2,943 (7%) did not pass one or more of our QC metrics and were subsequently excluded. Criteria for exclusion were as follows: disagreement between genetically determined and reported sex (n = 900); high rates of heterozygosity or contamination (VBID > 5%) (n = 709); low sequence coverage (less than 80% of targeted bases achieving 20× coverage) (n = 115); genetically identified sample duplicates (n = 1,662 total samples); WES variants discordant with the genotyping chip (n = 43). The remaining n = 37,804 (37 K) samples were then used to compile a project-level VCF (PVCF) for downstream analysis using the GLnexus joint genotyping tool. This final dataset contained n = 7,655,430 variants. Within this dataset of 37 K exomes, stroke case:control status was known for n = 31,737, referred to as the 31 K discovery cohort. The remaining n = 6,067 were part of the 41 K follow-up cohort.

Exome sequencing in the replication 30 K cohort (n = 39,399) was conducted by the CNCD using a publicly available protocol [34]. Briefly, blood derived DNA samples, with 10 to100 ng concentration of initial genomic DNA, underwent hybridization and capture using Illumina Rapid Cature Exome Kit or Agilents SureSelect Human Exon v2. Samples were denatured and amplified HiSeq v3 cluster chemistry and HiSeq 2000 or 2500 flowcells based on the manufacturers protocol. Reads were aligned to the GRCh38 genome reference and variants were called using GATK v.30 folllowed by variant recaliberation to remove false positive variants.

The remaining n = 7,616 exome samples of the 75 K PGR consisted of whole genome sequence (WGS) data that produced a VCF subsequently filtered to include only variants in protein coding sequence. WGS samples were sequenced and process as described in a publicly available protocol (https://www.nature.com/articles/s41586-021-03205-y). Briefly, 30x whole genome sequencing was performed using Illumina HiSeqX instruments. Reads were aligned to the GRCh38 reference using BWA-align and variants were called using the publicly available GotCloud pipeline (https://genome.sph.umich.edu/wiki/GotCloud), which includes QCing variants based on a support vector machine trained on specific site quality metrics.

### Variant calling

The PGR discovery cohort WES data was reference-aligned using the OQFE protocol [35], which uses BWA MEM to map all reads to the GRCh38 reference in an alt-aware manner, marks read duplicates and adds additional per-read tags. The OQFE protocol retains all reads and original quality scores such that the original FASTQ is completely recoverable from the resulting CRAM file. Single-sample variants were called using DeepVariant with custom exome parameters [35], generating a gVCF for each input OQFE CRAM file. These gVCFs were aggregated and joint-genotyped using GLnexus (v.1.3.1). All constituent steps of this protocol were executed using open-source software. The PGR replication and follow-up cohort were analyzed using the publicly available GotCloud workflow (https://genome.sph.umich.edu/wiki/GotCloud).

### Identification of low-quality variants from sequencing using machine learning

Similar to other recent large-scale sequencing efforts, we implemented a supervised machine-learning algorithm to discriminate between probable low-quality and high-quality variants [36, 37]. In brief, we defined a set of positive control and negative control variants based on the following criteria: (1) concordance in genotype calls between array and exome-sequencing data; (2) transmitted singletons; (3) an external set of likely ‘high quality’ sites; and (4) an external set of likely ‘low quality’ sites. To define the external high-quality set, we first generated the intersection of variants that passed QC in both TOPMed Freeze 8 and GnomAD v.3.1 genomes. This set was additionally restricted to 1000 Genomes Phase 1 high-confidence SNPs from the 1000 Genomes Project [38] and gold-standard insertions and deletions from the 1000 Genomes Project and a previous study [39], both available through the GATK resource bundle (https://gatk.broadinstitute.org/hc/en-us/articles/360035890811-Resource-bundle). To define the external low-quality set, we intersected GnomAD v3.1 fail variants with TOPMed Freeze 8 Mendelian or duplicate discordant variants. Before model training, the control set of variants were binned by allele frequency and then randomly sampled such that an equal number of variants were retained in the positive and negative labels across each frequency bin. A support vector machine using a radial basis function kernel was then trained on up to 33 available site quality metrics, including, for example, the median value for allele balance in heterozygote calls and whether a variant was split from a multi-allelic site. We split the data into training (80%) and test (20%) sets. We performed a grid search with fivefold cross-validation on the training set to identify the hyperparameters that returned the highest accuracy during cross-validation, which were then applied to the test set to confirm accuracy. This approach identified a total of *n* = 931,823 WES variants as low-quality, resulting in a dataset of n = 6,723,607 variants.

### Variant annotation

Variants were annotated as described in a publicly available pipeline <u>38</u>. In brief, variants were annotated using Ensembl variant effect predictor, with the most severe consequence for each variant chosen across all protein-coding transcripts. In addition, we derived canonical transcript annotations based on a combination of MANE, APPRIS and Ensembl canonical tags. MANE annotation was given the highest priority followed by APPRIS. When neither MANE nor APPRIS annotation tags were available for a gene, the canonical transcript definition of Ensembl was used. Gene regions were defined using Ensembl release 100. Variants annotated as stop gained, start lost, splice donor, splice acceptor, stop lost or frameshift, for which the allele of interest was not the ancestral allele, were considered predicted loss-of-function variants. Five annotation resources were utilized to assign deleteriousness to missense variants: SIFT, Polyphen2_HDIV, Polyphen2_HVAR, LRT, MutationTaster [40–43], and LRT, obtained using dbNSFP [44]. Missense variants were considered ‘likely deleterious’ if predicted deleterious by all five algorithms, ‘possibly deleterious’ if predicted deleterious by at least one algorithm and ‘likely benign’ if not predicted deleterious by any algorithm.

### Pakistan Genome Resource Statistical Analysis

ExWAS of SNPs with minor allele count > 5 in the 31 K PGR discovery cohort was conducted with 5 binary stroke phenotypes using REGENIE (v 3.1.1) [26] with age, age^2^, sex, age*sex, exome batch, 10 genotyping array principal components (PCs), 10 common variant exome PCs and 10 rare variant exome PCs as covariates. The minimum of 1,000 cases was selected based on a power calculation [45] (1,000 cases; 25,000 controls; significance threshold 0.005; prevalence 0.0012; disease allele frequency 0.005; genotype relative risk 3.0; > 80% power). Follow-up analyses were conducted with the added covariates of hypertension and tobacco use, or as environmental factors in a gene-by-environment interaction test using REGENIE [26]. Gene burden analysis was conducted using REGENIE with separate masks for pLoF, pLoF + missense, pLoF + deleterious missense (as predicted by at least 1 of 5 algorithms), and pLoF + deleterious missense (as predicted by 5 of 5 algorithms). Analysis in the 61 K PGR meta-analysis cohort, including 31 K discovery and 30 K replication cohorts, was conducted using REGENIE.

### PGR Population Genetic Analysis

Using principal components (PCs) [46] and Uniform Manifold Approximation and Projection (UMAP) [47] based analyses PGR and UKB South Asian sub-populations were mapped to distinct groups or clusters. Specifically, we used the imputed genotypes to merge the PGR dataset with UKB and 1000 genome datasets. Imputed data was used to maximize the number of common variants between all three datasets. The Plink [48] “--bmerge” option was used to merge datasets. A minimal QC was applied to the merged genotypes to exclude variants with MAF less than 5%, missing genotype rate greater than 10%, and Hardy Weinberg equilibrium P value less than 5 x 10^-5^. Variants mapping within the HLA region were excluded. Merged datasets were pruned for linkage disequilibrium (r^2^ > 0.25). A total of 20 PCAs were calculated in the merged data using the Plink –pca option. Calculated PCAs were imported to R and merged with reported ethnicities or country of birth information. The first 6 PCs calculated on the merged data were reduced to two dimensions using the UMAP package in R. The two eigenvectors of UMAP were calculated using an alpha value of 1.1 and beta value of 0.8. Two eigenvectors were plotted along with ethnicity and country of birth labels using the Plotly package in R. UKB self-reported ethnicities or country of birth was confirmed to be highly correlated with data obtained from UMAP.

### Population Attributable Fraction

Estimation of the proportion of all strokes combined or hemorrhagic stroke in Pakistan population attributable to p.Arg1231Cys was calculated using the formula [27],

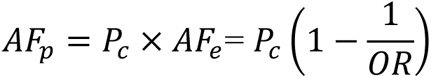

where *P_c_* is the prevalence of mutation among cases, *AF_e_* is the attributable fraction in the exposure, and *OR* was for risk of stroke (i.e., all stroke combined or hemorrhagic stroke) comparing mutant vs wild type of p.Arg1231Cys in the discovery cohort.

OR was obtained directly from Supplementary Table 2. The related 95% confidence intervals were constructed using bootstrap method with 10,000 resamples [49], which was implemented with the command “Bootstrap” using Stata (College Station, Texas 77845 USA).

### Recall by Genotype

A subset of carriers of *NOTCH3* p.Arg1231Cys were contacted by the Center of Non-Communicable Diseases in Karachi Pakistan under protocols approved by the IRB committee of the Center for Non-Communicable Diseases (NIH registered IRB 00007048). After obtaining consent from the proband and from the family members, questionnaires regarding past medical and family history were administered by trained research staff, in the local language. Physical measurements such as height, weight, hip and waist circumference were obtained in the standing position by using height and weight scales. Blood pressure and heart rate were recorded sitting by using OMRON healthcare M2 blood pressure monitors. Non-fasting blood samples were collected from each participant in EDTA and Gel Tubes. Serum and plasma were separated within 45 minutes of venipuncture. A random urine sample was also collected from each participant. The samples were stored temporarily in dry ice in the field and transported to the central laboratory based at CNCD and stored at -80 degrees Celsius. Measurements for total-cholesterol, HDL cholesterol, LDL cholesterol, triglycerides, VLDL, AST, ALT and creatinine were obtained from serum samples using enzymatic assays. HbA1c was measured using a turbidemetric assay in whole-blood samples (Roche Diagnostics). All measurements were done at a central laboratory at CNCD. Statistical analysis comparing across genotypes was conducted using the numpy library in Python 3.11.4.

### PGR Stroke Case Control Definitions

• Controls

• Inclusion

• No medical history of stroke, myocardial infarction (MI), coronary artery disease, heart failure (HF), valvular disease, or pacemaker
Cases

• Inclusion

• General Criteria

◦ Stroke: Diagnosis of ‘Stroke’
◦ Ischemic: Diagnosis of ’Ischemic stroke’
◦ Hemorrhagic: Diagnosis of ’Hemorrhagic stroke’

∙ Subcortical: Type of intracerebral hemorrhage = ’Subcortical’
∙ Parenchymal: Type of intracerebral hemorrhage = ‘Parenchymal’
• Oxfordshire Criteria

◦ Partial anterior circulation infarct (PACI): Partial anterior circulation infarcts (PACI) stroke sub-type
◦ Posterior circulation infarction (POCI): Posterior circulation infarcts POCI stroke sub-type
◦ Total anterior circulation infarct (TACI): Total anterior circulation infarcts TACI’ stroke sub-type
◦ Lacunar infarct (LACI): Lacunar infarcts stroke sub-type
• TOAST Criteria

◦ Cardioembolism (CE): Cardioembolism ischemic stroke subtype
◦ CE probable: CE criteria, in addition diagnosis is made if the clinical findings, neuroimaging data, and results of diagnostic studies are consistent with one subtype and other etiologies have been excluded
◦ Large artery atherosclerosis (LAA): Large artery atherosclerosis ischemic stroke subtype based on TOAST classification
◦ LAA probable: LAA criteria, in addition in addition diagnosis is made if the clinical findings, neuroimaging data, and results of diagnostic studies are consistent with one subtype and other etiologies have been excluded
◦ Small artery atherosclerosis (SAA): Small artery atherosclerosis ischemic stroke subtype
◦ SAA probable: SAA criteria, in addition in addition diagnosis is made if the clinical findings, neuroimaging data, and results of diagnostic studies are consistent with one subtype and other etiologies have been excluded

### UK Biobank Stroke Case Control Definition

UK Biobank stroke case control definitions were based on ICD10 codes as follows.

- Cases

◦ Inclusion

∙ Phe10_I63, ICD10 3D: Cerebral infarction
∙ Phe10_I630, ICD10 4D: Cerebral infarction due to thrombosis of precerebral arteries
∙ Phe10_I631, ICD10 4D: Cerebral infarction due to embolism of precerebral arteries
∙ Phe10_I632, ICD10 4D: Cerebral infarction due to unspecified occlusion or stenosis of precerebral arteries
∙ Phe10_I633, ICD10 4D: Cerebral infarction due to thrombosis of cerebral arteries
∙ Phe10_I634, ICD10 4D: Cerebral infarction due to embolism of cerebral arteries
∙ Phe10_I635, ICD10 4D: Cerebral infarction due to unspecified occlusion or stenosis of cerebral arteries
∙ Phe10_I638, ICD10 4D: Other cerebral infarction
∙ Phe10_I639, ICD10 4D: Cerebral infarction, unspecified
∙ Self-reported: SR_1583_ischaemic_stroke
∙ Primary and secondary cause of death using above ICD codes.
◦ Exclusion

∙ Phe10_I636, ICD10 4D: Cerebral infarction due to cerebral venous thrombosis, nonpyogenic
- Controls

◦ Inclusion

• Negative for the above codes
• Negative for Phe10_Z823, ICD10 4D: Family history of stroke
2. Exclusion:

• Phe10_G45, ICD10 3D: Transient cerebral ischemic attacks and related syndromes
• Phe10_G458, ICD10 4D: Other transient cerebral ischemic attacks and related syndromes
• Phe10_G459, ICD10 4D: Transient cerebral ischemic attack, unspecified

### Custom Burden Tests in UKB 450 K

Burden tests aim to boost statistical power by aggregating association signal across multiple rare variants. Prior studies in human and animal models have debated the role of various variant classes on *NOTCH3* function, CADASIL pathology and patient prognosis, including experiments designed to determine if the pathogenicity of CADASIL variants follows a loss of function (LoF) mechanism [8, 25, 50]. Using data from hundreds of missense and LoF variants in *NOTCH3* observed in 450 K UKB participant exomes, burden tests were conducted to assess the impact of LoF and missense variants.

Ten distinct gene burden tests of association with stroke were conducted using REGENIE [26], divided into three distinct groups. The Group I tests assessed the impact of Cys-altering variants, Group II tests assessed the impact of Ser-altering variants, and Group III tests assessed LoF and all missense variants. Group I and II consisted of four distinct tests, including (1) a test of all group variants in EGFr domains 1 to 34, (2) a test of all group variants in EGFr domains 1 to 6, (3) a test of all group variants in EGFr domains 7 to 34, and (4) a test of all group variants outside of EGFr domains. The difference between Group I and Group II was Group I variants are missense variants that either add or remove a Cysteine (Cys-altering), while Group II variants are missense variants that either add or remove a Serine (Ser-altering). While the role of Cys-altering variants in CADASIL is well known [15, 19, 20], Ser-altering variants were chosen based on being the most common class of variants among *NOTCH3* variants in UKB 450 K exomes. Group III consisted of two tests, including (1) a test limited to LoF variants and (2) a test limited to LoF and missense variants.

## Data Availability

The primary data in this study not already presented in the manuscript and supplement consists of ExWAS summary statistics for 5 stroke phenotypes analyzed in the PGR cohort. This data is publicly available in the GWAS Catalog under accessions GCST90432122, GCST90432123, GCST90432124, GCST90432125, and GCST90432126.

## Code Availability

Data collection and analysis software, tools, algorithms, and packages used in this manuscript are publicly available in software repositories. Below is a list of the softwares used, their versions, and contemporary links to public repositories.

### Data Production

Read alignment was conducted using BWA MEM v.0.7.17. Variant calling was conducted using Deep Variant v.0.10.0. Joint genotyping was conducted using GLnexus v1.3.1. Variant deleteriousness was calculated using SIFT v.2011, Polyphen 2 v.2011, LRT v.2013, MutationTaster v.4.3, and dbNSFP v.3.2. Single variant ExWAS and gene burden test of PGR and UKB data was conducted using REGENIE v.3.1.3.

### Data Analysis

Figures were plotted using R v.4.4.1. Data management was conducted using Python v.3.11.4. Statistical tests in Table 1 were conducted using R stats library v.4.3.0. Supplementary Figure 1 and 2 were produced using R UpSetR library v.1.4.0. Supplementary Figures 3 and 4 were produced using R QQMAN library v.0.1.8. Supplementary Figure 5 was produced using R Plotly library v.4.10.1. Supplementary Figure 6 was produced using R v.4.4.1. Supplementary Figure 7 was produced using ITK-SNAP v.3.8.0. Principal Components Analysis was conducted using PLINK v.1.9. Uniform Manifold Approximation and Projection was conducted using UMAP v.0.2.10.0. Power calculation was conducted using GAS Power Calculator v.2017. Meta Analysis in Figure 3 was conducted using METAL v.2011-03-25. MRI Image Analysis was conducted using FMRIB Software Library (FSL) v.5.0.10 and FreeSurfer v.6.0. Alignment in Figure 2 was conducted using BLAST v.2.14.

## Acknowledgements

Supported by Regeneron Pharmaceuticals.

This research has been conducted using the UK Biobank Resource (project 26041). The authors thank everyone who made this work possible, particularly the UK Biobank team, their funders, the professionals from the member institutions who contributed to and supported this work, and most especially the UK Biobank participants, without whom this research would not be possible. The exome sequencing was funded by the UK Biobank Exome Sequencing Consortium (Bristol Myers Squibb, Regeneron, Biogen, Takeda, Abbvie, Alnylam, AstraZeneca and Pfizer). Ethical approval for the UK Biobank was previously obtained from the North West Centre for Research Ethics Committee (11/NW/0382).

Disclosure forms provided by the authors are available with the full text of this article. Drs. Rodriguez-Flores, Khalid, Shuldiner and Saleheen contributed equally to this article.

## Inclusion and Ethics

The Institutional Review Board (IRB) at the Center for Non-Communicable Diseases (IRB: 00007048, IORG0005843, FWAS00014490) approved the study. All participants gave written informed consent.

## Author Contributions Statement

Conceptualization by Juan L. Rodriguez-Flores (JLRF), Shareef Khalid (SK), Alan R. Shuldiner (ARS), and Danish Saleheen (DS).

Data curation by Nilanjana Banerjee (NB), Deepika Sharma (DeS), Michael Cantor (MC), John Overton (JO), and Jeff Reid (JR).

Formal analysis by Bin Ye (BY), Manav Kapoor (MK), Joshua Backman (JB), Gannie Tzoneva (GT), Ellen Tsai (ET), Sahar Gelfman (SG), Tanima De (TD), Niek Verweij (NV), Luca A. Lotta (LAL), Aaron Zhang (AZ), Neelroop Parikshak (NP), Farshid Sepehrband (FS), Jonathan Marchini (JM), Giovanni Coppola (GC), Sofia Castaneda (SC), Pengcheng Xun (PX), Ellen Tsai (ET), and Regeneron Genetics Center (RGC).

Funding acquisition by DS, ARS, Aris Baras (AB), and RGC.

Investigation by Silvio Alessandro DiGioia (SADG), Hector Martinez (HM), I-Chun Tsai (IT), Katia Karalis (KK), Aris Economides (AE), David D’Ambrosio (DDA), and Asif Rasheed (AR). Methodology by JLRF, SK, and RGC.

Project administration by Thomas Coleman (TC), RGC, and AR.

Resources by Muhammad Jahanzaib (MJ), Maleeha Zaman (MZ), Muhammad Rehan Mian (MRM), Muhammad Bilal Liaqat (MBL), Khalid Mahmood (KM), Tanvir-us-Salam (TUS), Muhammad Hussain (MH), Ayeesha Kamal (AK), Javed Iqbal (JI), and Faizan Aslam (FA).

Software by RGC, JLRF, and SK. Supervision by DS and ARS. Validation by SK and JLRF. Visualization by JLRF, NP, FS, JM, and RGC.

Writing of original draft by JLRF, SK, ARS, DS, PX, Sergio Fazio (SF), Wolfgang Liedtke (WL), John Danesh (JD), Ayeesha Kamal (AK), Philippe Frossard (PF), and RGC.

Writing review and editing by JLRF, PX, SK, ARS, DS, and RGC.

## Competing Interests Statement

### Employment

JLRF, ARS, NB, DeS, MC, JO, JR, BY, MK, JB, GT, SG, TD, NV, LAL, AZ, NP, FS, JM, GC, PX, AB, SADG, HM, IT, KK, AE, DDA, SF, WL, and TC are or were employees of Regeneron Genetics Center LLC or Regeneron Pharmaceuticals Inc. and contributed to this manuscript as part of their regular duties as salaried employees.

ET and SC are or were student interns of Regeneron Genetics Center LLC or Regeneron Pharmaceuticals Inc. and contributed to this manuscript as part of their internship activites.

AR, MJ, MZK, MRM, MBL, PF, and DS and SK are or were employees of the Center for Non-Communicable Disease and received salaried compensation for their contribution to this manuscript.

### Personal Financial Interests

JLRF, ARS, NB, DeS, MC, JO, JR, BY, MK, JB, GT, SG, TD, NV, LAL, AZ, NP, FS, JM, GC, PX, AB, SADG, HM, IT, KK, AE, DDA, SF, WL, TC are or were employees of Regeneron Genetics Center LLC or Regeneron Pharmaceuticals Inc. and received stock and stock options as part of their compensation as employees.

JLRF, ARS, DS, AB, and SK are named inventors on patent pending US 20230000897A1 that discloses methods of treating subjects having a cerebrovascular disease by administering Neurogenic Locus Notch Homolog Protein 3 (*NOTCH3*) agents, and methods of identifying subjects having an increased risk of developing a cerebrovascular disease.

