## Supplement for "*NOTCH3* p.Arg1231Cys is Markedly Enriched in South Asians and Associated with Stroke"

**for**

**Correspondence**

**Table of Contents**

Supplementary Data
Supplementary Tables
Supplementary Figures
Supplementary References
Supplementary Figures

### Supplementary Data

**Population Genetic Analysis**

In order to better understand the population frequencies of p.Arg1231Cys variant and its implications for stroke risk across South Asia (SAS) and neighboring West Asia / North Africa (also referred to as the Greater Middle East, abbreviated GME), a regional survey of p.Arg1231Cys was conducted. For these analyses, public datasets such as 1000 Genomes Phase 3 [1], GnomAD [2], Greater Middle East (GME) Variome [3], Iranome [4], and QChip Knowledgebase [5] were used. p.Arg1231Cys was enriched (AAF > 0.001) in most GME and SAS populations sampled.

- QChip Knowledgebase, Qatar (AAF = 0.0075)

- GME Variome (AAF = 0.004)
  - Allele Counts (RR:RA:AA)
    - Northwest Africa (98:01:00), AAF = 0.0051
    - Northeast Africa (367:01:00); AAF = 0.0014
    - Arabian Peninsula (168:03:00); AAF = 0.0088
    - Israel (10:00:00); AAF = 0.0
    - Syrian Desert (58:00:00); AAF = 0
    - Turkish Peninsula (163:01:00); AAF = 0.003
    - Central Asia (130:02:00); AAF = 0.0076

- Iranome
  - Lur (AAF = 0.01)
  - Baloch (AAF = 0.005)
  - Kurd (AAF = 0.005)
  - Persian (AF = 0.005)
  - Turkmen (AF = 0.005)

- GnomAD, Pooled SAS (AAF=0.00536)

- 1000 Genomes Phase 3
  - Aggregate SAS (AAF=0.0041)
  - GIH, Gujarati Indians in Houston, USA (AAF = 0.0049)
  - ITU, Indian Telugu in the UK (AAF=0.0049)
  - PJL, Punjabi in Lahore, Pakistan (AAF= 0.0052)
  - STU, Sri Lankan Tamil in the UK (AAF= 0.0049)
  - BEB, Bengali in Bangladesh (not detected)

- UKB Participants born in SAS or GME (n > 100)
  - SAS (aggregate AAF = 0.0042),
    - Afghanistan (AAF = 0.005)
    - Pakistan (AAF = 0.003)
    - India (AAF = 0.0041)
  - GME (aggregate AAF = 0.0031)
    - Egypt (not detected)
    - Turkey (AAF = 0.0029)
    - Iran (AAF = 0.0049)

**Variant Prevalence in Pakistani Population Clusters**

Combined analysis of 37 K exomes from the PGR, 1000G and UKB identified three major clusters of Pakistanis, and ethnic/caste data was used to label them as Punjab/Rajput which clustered with 1000G Punjab; Ansari, which formed a distinct cluster closer to populations from Eastern India; and Baloch/Pathan which clustered with Afghans (**Supplementary Figure 5, Supplementary Table 9)**. The variant was enriched (AAF > 0.005) in all three clusters.

- Baloch/Pathan (AAF = 0.0074)
- Punjab/Rajput (AAF = 0.006)
- Ansari (AAF = 0.0056)

**Supplementary Tables**

**Supplementary Table 1. ExWAS in PGR Discovery Cohort across 5 Stroke Phenotypes [1].**

| # | Phenotype | N Cases[1] | N Controls | Total |
|---|---|---|---|---|
| 1 | Stroke | 5,135 | 26,602 | 31,737 |
| 2 | Ischemic | 2,619 | 26,602 | 29,221 |
| 3 | Intracerebral hemorrhage | 1,533 | 26,602 | 28,135 |
| 4 | Subcortical intracerebral hemorrhage | 1,388 | 26,602 | 27,990 |
| 5 | Partial anterior circulation infarcts (PACI) | 1,274 | 26,602 | 27,876 |

1. Shown are 5 stroke phenotypes that were ascertained for cases using TOAST [6] and Oxfordshire [7] criteria. From left-to-right is the phenotype name, detailed description, number of cases, number of controls, and total sample size among the PGR discovery cohort participants.
2. While other ischemic stroke subtypes were identified using TOAST and Oxfordshire criteria, this study was limited to subtypes with at least 500 cases. Other ischemic stroke subtypes (including posterior circulation infarction, total anterior circulation infarct, lacunar infarct, cardioembolism, large artery artherosclerosis, and small artery artherosclerosis) had fewer than 500 cases. Using the University of Michigan Genetic Association Study Power Calculator [8], we estimated power over a range of alternate allele frequency (AAF) and genotype relative risk (GRR) values. Looking across a range of AAF's, we estimate that a variant would need to have 0.11 AAF to achieve 80% power with 500 cases and 26,602 controls, significance level of $5.0x10^{-8}$, disease prevalence of 0.0011 and a genotype relative risk of 2.0. Looking across a range of GRRs, we estimate that a variant with a AAF of 0.01 would need to have a GRR of 6.0 to achieve 80% power with 500 cases and 26,602 controls, significance level of $5.0x10^{-8}$, disease prevalence of 0.0011. With less than 500 cases, power to detect association at the genome-wide level of significance ($5.0x10^{-8}$) would require a very large effect size (e.g. 6.0).

**Supplementary Table 2. Association Summary Statistics for p.Arg1231Cys *NOTCH3* and 5 Stroke Phenotypes in PGR Discovery Cohort[1].**

| Phenotype (ordered by increasing odds ratio) | Odds Ratio (95% Confidence Interval) | P value | AAF | Num Cases | Num Controls |
|---|---|---|---|---|---|
| Ischemic | 1.80 (1.23, 2.64) | 2.49E-03 | 5.64E-03 | 2540\|42\|2 | 25619\|271\|2 |
| Partial anterior circulation infarcts (PACI) | 1.99 (1.23, 3.22) | 4.84E-03 | 5.56E-03 | 1227\|25\|1 | 25619\|271\|2 |
| Stroke | 2.2 (1.7, 2.9) | 4.93E-08 | 6.10E-03 | 4939\|98\|3 | 25619\|271\|2 |
| Intracerebral hemorrhage | 3.17 (2.12, 4.73) | 1.79E-08 | 5.82E-03 | 1451\|44\|0 | 25619\|271\|2 |
| Subcortical intracerebral hemorrhage | 3.39 (2.26, 5.1) | 3.87E-09 | 5.84E-03 | 1313\|43\|0 | 25619\|271\|2 |

1. Shown are ExWAS summary statistics for 5 stroke phenotypes and the *NOTCH3* p.Arg1231Cys variant. From left-to-right is the phenotype, effect size, effect size 95% confidence interval (lower, upper), p value, alternate allele frequency (AAF), cases count subdivided by genotype (homozygous reference | heterozygous | homozygous alternate) and controls count subdivided by genotype (homozygous reference | heterozygous | homozygous alternate).

**Supplementary Table 3. Phenome-wide Association Summary Statistics for p.Arg1231Cys *NOTCH3* and Comorbidities and Related Binary Phenotypes in PGR Discovery Cohort[1].**

| Phenotype (Binary Traits) | Odds Ratio (95% Confidence Interval) | P value | AAF | Cases | Controls |
|---|---|---|---|---|---|
| Subcortical intracerebral hemorrhage | 3.4 (2.3, 5.1) | 3.87E-09 | 5.80E-03 | 1313\|43\|0 | 25619\|271\|2 |
| Intracerebral hemorrhage diagnosis | 3.2 (2.1, 4.7) | 1.80E-08 | 5.80E-03 | 1451\|44\|0 | 25619\|271\|2 |
| Stroke | 2.2 (1.7, 2.9) | 4.93E-08 | 6.10E-03 | 4939\|98\|3 | 25619\|271\|2 |
| Partial anterior circulation infarcts (PACI) | 2 (1.2, 3.2) | 4.85E-03 | 5.60E-03 | 1227\|25\|1 | 25619\|271\|2 |
| Lacunar cerebral infarct (LACI) | 2.9 (1.3, 6.3) | 7.05E-03 | 5.40E-03 | 364\|8\|1 | 25619\|271\|2 |
| Stroke, undetermined cause | 1.9 (1.2, 3) | 7.81E-03 | 5.50E-03 | 1395\|26\|1 | 25619\|271\|2 |
| Pain in lower extremities while walking | 2.4 (1.2, 4.7) | 1.26E-02 | 6.80E-03 | 454\|11\|0 | 20286\|265\|4 |
| Right or left ventricular hypertrophy diagnosis, S in V3 + R in avl > 20 mm in women | 2.7 (1.2, 6) | 1.27E-02 | 6.10E-03 | 286\|9\|0 | 7908\|92\|0 |
| Stroke diagnosis self-reported | 1.9 (1.1, 3.1) | 1.30E-02 | 6.90E-03 | 856\|21\|0 | 20540\|271\|4 |
| Self-reported myocardial infarction (MI) | 0.69 (0.51, 0.94) | 1.85E-02 | 5.90E-03 | 5319\|45\|0 | 30641\|376\|5 |
| Small artery atherosclerosis, TOAST classification | 2.6 (1.1, 5.8) | 2.30E-02 | 5.40E-03 | 309\|7\|1 | 25619\|271\|2 |
| Lower extremities pain gets better on resting | 2.7 (1.1, 6.5) | 2.90E-02 | 8.30E-03 | 277\|10\|0 | 7192\|110\|3 |
| Right or left ventricular hypertrophy diagnosis, S in lead V1 + R in V5/V6 > 35mm | 1.7 (1, 2.9) | 3.49E-02 | 6.20E-03 | 839\|18\|0 | 16918\|198\|3 |
| Stroke family history | 1.6 (1, 2.4) | 4.07E-02 | 5.90E-03 | 1406\|25\|0 | 34993\|398\|5 |
| Self-reported heart failure (HF), first occurrence | 0.63 (0.41, 0.98) | 4.17E-02 | 4.40E-03 | 4173\|25\|0 | 8439\|86\|1 |
| Clopidogrel medication for heart disease | 0.63 (0.4, 0.99) | 4.36E-02 | 5.90E-03 | 2248\|17\|0 | 31074\|373\|5 |
| Hypertension family history, mother | 0.7 (0.48, 1) | 5.69E-02 | 5.80E-03 | 2683\|22\|0 | 31500\|376\|3 |
| Hypertension family history, father | 0.68 (0.46, 1) | 5.75E-02 | 5.80E-03 | 2348\|18\|0 | 31841\|380\|3 |
| Type of myocardial infarction (MI), anterior | 1.5 (0.95, 2.3) | 8.67E-02 | 5.90E-03 | 1819\|29\|0 | 34580\|394\|5 |
| Total anterior circulation infarcts (TACI) | 0.41 (0.15, 1.1) | 8.81E-02 | 5.20E-03 | 441\|1\|0 | 25619\|271\|2 |
| Diabetes family history, mother | 0.75 (0.54, 1.1) | 1.02E-01 | 5.80E-03 | 3262\|29\|0 | 30927\|369\|3 |
| Self-reported history of diabetes | 0.84 (0.67, 1.1) | 1.47E-01 | 5.90E-03 | 8634\|86\|1 | 27270\|335\|3 |
| Posterior circulation infarcts (POCI) | 2 (0.78, 5.1) | 1.50E-01 | 5.40E-03 | 469\|8\|0 | 25619\|271\|2 |
| Stroke family history, father | 1.7 (0.81, 3.6) | 1.59E-01 | 5.80E-03 | 615\|11\|0 | 33575\|387\|3 |
| Intracranial large artery atherosclerosis (LAA) | 2.8 (0.66, 11) | 1.64E-01 | 5.30E-03 | 270\|4\|0 | 25619\|271\|2 |
| Large artery atherosclerosis (LAA) | 1.9 (0.64, 5.8) | 2.47E-01 | 5.30E-03 | 366\|6\|0 | 25619\|271\|2 |
| Type of myocardial infarction (MI), inferior | 0.68 (0.35, 1.3) | 2.48E-01 | 5.90E-03 | 972\|6\|0 | 35427\|417\|5 |
| Self-reported depression | 0.36 (0.059, 2.2) | 2.68E-01 | 4.50E-03 | 128\|0\|0 | 12490\|112\|1 |
| Stroke family history, mother | 1.6 (0.7, 3.5) | 2.82E-01 | 5.80E-03 | 530\|9\|0 | 33659\|389\|3 |
| Heart failure family history | 1.8 (0.59, 5.5) | 3.03E-01 | 5.90E-03 | 284\|5\|0 | 36115\|418\|5 |
| Myocardial infarction family history | 1.1 (0.86, 1.5) | 3.76E-01 | 5.90E-03 | 5669\|69\|1 | 30730\|354\|4 |
| Myocardial infarction | 0.91 (0.72, 1.1) | 4.27E-01 | 5.70E-03 | 11386\|118\|1 | 21370\|254\|3 |
| Diabetes controlled or not | 1.4 (0.62, 3) | 4.47E-01 | 1.00E-02 | 815\|16\|1 | 610\|12\|0 |
| Smoked more than 100 cigarette or beedies | 0.74 (0.34, 1.6) | 4.51E-01 | 5.80E-03 | 6406\|72\|1 | 824\|9\|1 |
| Heart failure (HF) case vs control | 0.86 (0.57, 1.3) | 4.54E-01 | 4.50E-03 | 7702\|63\|0 | 4921\|49\|1 |
| Myocardial infarction family history, mother | 1.2 (0.75, 1.8) | 5.14E-01 | 5.80E-03 | 1981\|26\|0 | 32208\|372\|3 |
| TOAST classification, cardio embolism | 0.69 (0.21, 2.2) | 5.29E-01 | 5.30E-03 | 412\|2\|0 | 25619\|271\|2 |
| Self-reported father and mother are first degree cousin relative | 0.94 (0.75, 1.2) | 6.00E-01 | 5.90E-03 | 8630\|94\|2 | 26401\|315\|3 |
| Diabetes family history, father | 0.9 (0.6, 1.4) | 6.06E-01 | 5.80E-03 | 2217\|23\|0 | 31972\|375\|3 |
| Self-reported high cholesterol | 0.73 (0.22, 2.5) | 6.10E-01 | 4.50E-03 | 310\|2\|0 | 12308\|110\|1 |
| Myocardial infarction family history, father | 1.1 (0.74, 1.6) | 6.69E-01 | 5.80E-03 | 2595\|32\|0 | 31590\|366\|3 |
| Tobacco use | 0.96 (0.77, 1.2) | 7.03E-01 | 5.90E-03 | 13161\|143\|1 | 22679\|276\|3 |
| Type of myocardial infarction (MI), anterior and anteroseptal | 1.3 (0.27, 6.6) | 7.18E-01 | 5.90E-03 | 179\|2\|0 | 36220\|421\|5 |
| Self-reported hypertension | 0.97 (0.79, 1.2) | 7.46E-01 | 5.80E-03 | 14958\|170\|1 | 20756\|244\|3 |
| Previous transient ischemic attack (TIA) | 1.2 (0.26, 5.1) | 8.40E-01 | 8.30E-03 | 125\|2\|0 | 7205\|116\|3 |
| Heart failure family history, mother | 1.1 (0.13, 8.3) | 9.62E-01 | 4.50E-03 | 105\|1\|0 | 12515\|111\|1 |

1. Shown are ExWAS summary statistics for stroke comorbidities and related binary phenotypes and the *NOTCH3* p.Arg1231Cys variant in the PGR discovery cohort. From left-to-right is the phenotype name, odds ratio (OR), OR effect size 95% confidence interval (lower, upper), p value, alternate allele frequency (AAF), cases count subdivided by genotype (homozygous reference | heterozygous | homozygous alternate) and controls count subdivided by genotype (homozygous reference | heterozygous | homozygous alternate).

 

**Supplementary Table 4. Phenome-wide Association Summary Statistics for p.Arg1231Cys *NOTCH3* and Comorbidities and Related Quantitative Phenotypes in PGR Discovery Cohort[1].**

| Phenotype (Quantitative Trait) | Beta (95% Confidence Interval) | P value | AAF | Sample size | Units |
|---|---|---|---|---|---|
| Sudden death age, father | -1 (-1.8, -0.3) | 6.18E-03 | 6.50E-03 | 382\|5\|0 | Years |
| Hemoglobin A1C | 0.24 (0.04, 0.44) | 1.86E-02 | 7.90E-03 | 5634\|88\|1 | % |
| Waist hip ratio | -0.087 (-0.18, 0.01) | 7.90E-02 | 6.00E-03 | 31972\|380\|3 | Decimal |
| Age at diagnosis of myocardial infarction (MI) | -0.29 (-0.63, 0.04) | 8.44E-02 | 3.50E-03 | 4690\|33\|0 | Years |
| Stroke diagnosis age, mother | -0.47 (-1, 0.067) | 8.59E-02 | 9.30E-03 | 476\|9\|0 | Years |
| Earliest sudden death age in family | -0.45 (-1, 0.13) | 1.28E-01 | 5.50E-03 | 804\|9\|0 | Years |
| Hypertension diagnosis age, brother | 0.33 (-0.11, 0.77) | 1.37E-01 | 5.70E-03 | 1301\|13\|1 | Years |
| Age quit paan | 0.59 (-0.19, 1.4) | 1.37E-01 | 4.40E-03 | 569\|5\|0 | Years |
| Systolic blood pressure | 0.079 (-0.026, 0.18) | 1.40E-01 | 5.90E-03 | 27322\|321\|2 | mmHg |
| Age started smoking | 0.15 (-0.056, 0.35) | 1.57E-01 | 5.80E-03 | 7177\|82\|1 | Years |
| Diastolic blood pressure | 0.073 (-0.028, 0.17) | 1.57E-01 | 5.90E-03 | 27679\|328\|2 | mmHg |
| Myocardial infarction diagnosis age, brother | -0.2 (-0.55, 0.15) | 2.56E-01 | 7.40E-03 | 1657\|23\|1 | Years |
| Triglycerides | -0.047 (-0.14, 0.046) | 3.20E-01 | 5.90E-03 | 34425\|402\|4 | Mg/dl |
| Diabetes diagnosis age, brother | -0.2 (-0.63, 0.24) | 3.73E-01 | 4.80E-03 | 1740\|17\|0 | Years |
| Age quit naswar | 0.21 (-0.29, 0.71) | 4.09E-01 | 1.70E-02 | 140\|5\|0 | Years |
| Heart failure (HF) diagnosis age | -0.17 (-0.6, 0.25) | 4.32E-01 | 2.70E-03 | 3743\|20\|0 | Years |
| Stroke diagnosis age, father | -0.19 (-0.73, 0.34) | 4.78E-01 | 7.90E-03 | 558\|9\|0 | Years |
| Diabetes diagnosis age, mother | -0.13 (-0.5, 0.24) | 4.85E-01 | 4.10E-03 | 2994\|25\|0 | Years |
| Earliest heart failure diagnosis age in family | -0.24 (-0.95, 0.46) | 5.03E-01 | 9.30E-03 | 264\|5\|0 | Years |
| Hypertension diagnosis age, sister | 0.18 (-0.36, 0.72) | 5.07E-01 | 5.70E-03 | 951\|11\|0 | Years |
| Age quit smoking | 0.16 (-0.31, 0.63) | 5.08E-01 | 5.10E-03 | 1443\|15\|0 | Years |
| Earliest hypertension diagnosis age in family | 0.087 (-0.19, 0.36) | 5.33E-01 | 4.50E-03 | 5254\|46\|1 | Years |
| High-density lipoprotein | -0.029 (-0.12, 0.062) | 5.33E-01 | 5.90E-03 | 34400\|402\|4 | Mg/dl |
| Earliest stroke diagnosis age in family | -0.12 (-0.52, 0.28) | 5.57E-01 | 8.10E-03 | 1275\|21\|0 | Years |
| Creatinine level in blood | -0.022 (-0.11, 0.07) | 6.37E-01 | 5.60E-03 | 32559\|360\|3 | Mg/dl |
| Stroke diagnosis age | -0.1 (-0.54, 0.33) | 6.40E-01 | 1.40E-02 | 539\|15\|0 | Years |
| Low-density lipoprotein (LDL) calculated | -0.022 (-0.12, 0.073) | 6.49E-01 | 5.90E-03 | 33742\|397\|4 | mg/dl |
| Myocardial infarction diagnosis age, sister | -0.14 (-0.75, 0.47) | 6.55E-01 | 6.60E-03 | 601\|8\|0 | Years |
| Number of Cigarettes / Beedies per day | 0.043 (-0.15, 0.24) | 6.70E-01 | 5.90E-03 | 6757\|79\|1 | Integer |
| Age started huqqa chillum | 0.098 (-0.38, 0.58) | 6.86E-01 | 1.20E-02 | 294\|5\|1 | Years |
| Diabetes diagnosis age, father | 0.08 (-0.32, 0.48) | 6.95E-01 | 4.90E-03 | 2015\|20\|0 | Years |
| Cholesterol | -0.019 (-0.11, 0.075) | 6.97E-01 | 5.90E-03 | 34552\|404\|4 | mg/dl |
| Hypertension diagnosis age, mother | 0.081 (-0.33, 0.49) | 6.98E-01 | 4.30E-03 | 2416\|21\|0 | Years |
| Earliest myocardial infarction diagnosis age in family | -0.044 (-0.27, 0.18) | 6.99E-01 | 6.30E-03 | 5420\|67\|1 | Years |
| Age at diagnosis of diabetes | -0.042 (-0.27, 0.18) | 7.12E-01 | 5.00E-03 | 7272\|74\|0 | Years |
| Hypertension diagnosis age, father | -0.077 (-0.54, 0.39) | 7.48E-01 | 3.80E-03 | 2114\|16\|0 | Years |
| Earliest diabetes diagnosis age in family | -0.032 (-0.27, 0.21) | 7.92E-01 | 5.00E-03 | 6118\|62\|0 | Years |
| Myocardial infarction diagnosis age, mother | -0.047 (-0.42, 0.32) | 8.03E-01 | 6.20E-03 | 1898\|24\|0 | Years |
| Diabetes diagnosis age, sister | 0.052 (-0.46, 0.56) | 8.44E-01 | 4.20E-03 | 1545\|13\|0 | Years |
| Age started gutka | -0.037 (-0.56, 0.49) | 8.90E-01 | 8.70E-03 | 623\|11\|0 | Years |
| Myocardial infarction diagnosis age, father | 0.015 (-0.33, 0.36) | 9.34E-01 | 5.60E-03 | 2477\|28\|0 | Years |
| Age at diagnosis of hypertension | -0.0067 (-0.17, 0.15) | 9.35E-01 | 5.80E-03 | 12072\|139\|1 | Years |
| Age started naswar | 0.019 (-0.49, 0.53) | 9.44E-01 | 5.50E-03 | 997\|11\|0 | Years |
| Glucose | -0.0027 (-0.098, 0.092) | 9.55E-01 | 5.90E-03 | 33746\|394\|3 | mg/dl |
| Age started paan | -0.0078 (-0.38, 0.36) | 9.67E-01 | 5.60E-03 | 2109\|24\|0 | Years |

1. Shown are ExWAS summary statistics for stroke comorbidities and the *NOTCH3* p.Arg1231Cys variant in the PGR discovery cohort. From left-to-right is the phenotype name, effect size (regression beta), effect size (95% confidence interval), p value, alternate allele frequency (AAF), and sample size count subdivided by genotype (homozygous reference | heterozygous | homozygous alternate).

**Supplementary Table 5. Inclusion of risk factors in regression model[1].**

| Phenotype (in order of increasing odds ratio) | Odds Ratio (95% Confidence Interval) | P value | AAF | Num Cases | Num Controls |
|---|---|---|---|---|---|
| Ischemic | 1.60 (1.08, 2.38) | 2.04E-02 | 5.52E-03 | 2471\|37\|2 | 24151\|256\|0 |
| Partial anterior circulation infarcts (PACI) | 1.81 (1.11, 2.97) | 1.80E-02 | 5.48E-03 | 1196\|23\|1 | 24151\|256\|0 |
| Stroke | 2.01 (1.49, 2.69) | 3.18E-06 | 6.02E-03 | 4696\|90\|3 | 24151\|256\|0 |
| Intracerebral hemorrhage | 2.96 (1.95, 4.5) | 3.64E-07 | 5.76E-03 | 1408\|42\|0 | 24151\|256\|0 |
| Subcortical intracerebral hemorrhage | 3.19 (2.08, 4.88) | 9.87E-08 | 5.77E-03 | 1273\|41\|0 | 24151\|256\|0 |

1. To assess the impact of other risk factors for stroke, statistical analysis was repeated in PGR discovery cohort cases and controls with smoking and hypertension as covariates in the model. Shown are results for association test between *NOTCH3* p.Arg1231Cys and five stroke phenotypes including standard covariates as well as smoking and hypertension. From left to right is the cohort, model, odds ratio (95% confidence interval), p value, alternate allele frequency, and genotype counts among cases and controls (reference homozygotes | heterozygotes | alternate allele homozygotes).

**Supplementary Table 6. Computational Prediction of Variant Deleteriousness[1].**

| Algorithm | Score | Interpretation |
|---|---|---|
| SIFT 4G converted rank score | 0.639 | Deleterious |
| PolyPhen2 Hum Div | 0.843 | Possibly Damaging |
| PolyPhen2 Hum Var | 0.462 | Possibly Damaging |
| Mutation Taster Rank Score | 0.588 | Deleterious |
| CADD | 25.1 | Pathogenic |

[1.] To computationally assess pathogenicity for *NOTCH3* p.Arg1231Cys, the variant was submitted to online portals for scoring using 5 algorithms. Details of the variant submitted included the GRCh38 coordinates Chr19:15179052, the reference and alternate allele G>A, and the ENSEMBL transcript ID ENST00000263388. Shown is a summary of the output, including the algorithm, the quantitative score reported and the interpretation of the score.

**Supplementary Table 7. *NOTCH3* p.Arg1231Cys Homozygotes in PGR[1].**

| Cohort[2] | ID[3] | Age (decade)[4] | Sex[5] | BMI[6] | Stroke | Type of Stroke | Migraine | Dementia | Depression | Hypertension | Diabetes | Other | Family History of CVD | Status at follow up |
|---|---|---|---|---|---|---|---|---|---|---|---|---|---|---|
| Follow-up | 1 | 50-59 | F | 26.6 | No | | No | No | No | Yes | No | MI, Angina | HTN | Angina |
| Discovery | 2 | 40-49 | M | NA | No | | No | No | No | No | No | MI | | Deceased |
| Discovery | 3* | 40-49 | M | 26.9 | No | | No | No | No | Yes | No | | | HTN |
| Replication | 4 | 40-49 | M | 31.2 | No | | No | No | No | No | No | | | |
| Follow-up | 5 | 50-59 | M | 27.2 | No | | No | No | No | Yes | No | | Stroke, HTN | |
| Replication | 6 | 50-59 | M | NA | No | | No | No | No | No | Yes | MI | | Sudden death |
| Discovery | 7 | 60-69 | M | NA | Yes | Ischemic stroke, Recurrent lacunar infarct | No | No | No | Yes | Yes | MI, Angina | | HTN |
| Discovery | 8 | 70-79 | M | 22.5 | Yes | Ischemic Stroke (PACI) | No | No | No | No | No | | HTN MI | |
| Discovery | 9 | 80-89 | M | 22.1 | Yes | Ischemic Stroke (PACI) | No | No | No | Yes | No | MI | | |
| Callback | 1 | 30-39 | F | 34.6 | No | | | No | No | Yes | No | Stomach issues | MI | |
| Callback | 2 | 40-49 | M | 26.7 | No | | | No | No | Yes | Yes | SVT | MI | |
| Discovery & Callback | 3* | 50-59 | M | 24.2 | No | | | No | No | Yes | No | | | |

1. Shown are relevant observations in the medical history of *NOTCH3* p.Arg1231Cys homozygotes identified in exome sequencing of the discovery cohort (top, numbered 1 to 9 by age and gender) or callback within families of exome-sequenced homozygotes (bottom, numbered 1 to 3 by age and gender), including , age, sex, BMI, stroke, type of stroke (if known), migraine with or without aura, dementia, depression, hypertension, diabetes, other family history of cardiovascular diseases, and status at follow up. Probands at the top were identified through exome sequencing of PGR, while homozygotes identified through callback are at the bottom. CVD = cardiovascular disease. HTN = hypertension. MI = myocardial infarction, SVT = supraventricular tachycardia
2. Cohorts include 31 K discovery 30 K replication, 14 K follow-up and n = 128 callback cohorts.
3. Individual #3 in top and bottom table are the same individual, aged in the 40s during recruitment and in the 50s during callback. Both individual IDs marked with an "*".
4. Age in decades
5. Sex: M=male, F=female
6. Body Mass Index (BMI) measured in $kg/m^2$

**Supplementary Table 8. Comparison of *NOTCH3* p.Arg1231Cys Genotypes in Callback Participants[1]**

| Phenotype | Counts (RR\|RA\|AA) | Homozygote RR | Heterozygote RA | Homozygote AA |
|---|---|---|---|---|
| Female | 75\|50\|3 | 43 (57.3%) | 25 (50%) | 1 (33.3%) |
| Age | 75\|50\|3 | 46 (36 - 57) | 50 (39 - 62) | 49 (40 - 52) |
| BMI (kg/m$^2$) | 74\|44\|3 | 27.5 (23 - 32) | 29 (26 - 35) | 27 (25 - 31) |
| Height (m) | 74\|45\|3 | 158 (153 - 166) | 161 (150 - 167) | 161 (157 - 165) |
| Weight (kg) | 74\|44\|3 | 71 (58 - 79) | 75.5 (64 - 90) | 75 (69 - 78) |
| WHR (Waist to Hip Ratio) | 73\|46\|3 | 0.91 (0.86 - 0.95) | 0.91 (0.88 - 0.97) | 0.96 (0.88 - 0.97) |
| Systolic BP | 68\|45\|3 | 127 (120 - 141.5) | 128 (116 - 132) | 145 (133 - 146.5) |
| Diastolic BP | 68\|45\|3 | 80.5 (78 - 90) | 80 (80 - 87) | 89 (83 - 91) |
| Cholesterol (mg/dl) | 73\|49\|3 | 162.9 (133 - 185.9) | 161.6 (129 - 190) | 168.4 (160 - 179) |
| TG (mg/dl) | 73\|49\|3 | 156 (119 - 189) | 194 (133 - 253) | 168 (128 - 235) |
| HDL (mg/dl) | 73\|49\|3 | 37 (30 - 44) | 31 (28 - 38) | 32 (27.5 - 36.5) |
| LDL (mg/dl) | 73\|49\|3 | 90 (71 - 112) | 84 (62 - 111) | 103 (85 - 117) |
| Fat Composition | 63\|36\|3 | 28.7 (24.5 - 38.15) | 35.35 (24.1 - 41.8) | 23.7 (20.8 - 36.05) |
| Impedance | 59\|33\|1 | 544 (477.5 - 589) | 519 (464 - 549) | 443 (443 - 443) |
| Fat Mass (kg) | 62\|35\|3 | 9.3 (6.76 – 12.11) | 11.02 (7.98-16.06) | 8.03 (6.53 – 12.79) |
| BMR (kJ) | 61\|35\|3 | 6086 (5356 - 6759) | 6185 (5751 - 7047) | 5676 (5656 - 6298) |
| FFM (kg) | 61\|35\|3 | 21.2 (18.55 – 24.95) | 22.1 (19.32 -24.61) | 23.1 (20.91 – 24.49) |
| Total Body Water (kg) | 61\|35\|3 | 15.5 (13.2 – 18.14) | 16.2 (14.36 – 17.87) | 16.9 (14.36 – 17.19) |
| Creatinine (mg/dl) | 73\|49\|3 | 0.78 (0.61 - 0.94) | 0.8 (0.65 - 1.03) | 0.92 (0.68 - 0.92) |
| eGFR (mL/min/1.73m$^2$) | 73\|49\|3 | 98 (79 - 111) | 89 (76 - 107) | 87 (86 - 128) |
| HbA1c (%) | 33\|19\|2 | 5.8 (5.4 - 6.64) | 5.89 (5.68 - 8.13) | 5.95 (5.88 - 6.02) |
| Gluscose (mg/dl) | 73\|49\|3 | 104.4 (81.5 - 123) | 104.2 (88.5 - 129.6) | 107.7 (91.8 - 147.9) |
| AST (U/L) | 73\|49\|3 | 21.8 (17.4 - 28) | 21.6 (17.7 - 25.8) | 24 (20.35 - 25.8) |
| ALT (U/L) | 73\|49\|3 | 19.1 (12.4 - 26.9) | 18.2 (14.9 - 25.4) | 27.7 (20.7 - 30.3) |
| WBC ($10^3$/μl) | 33\|23\|3 | 5.66 (4.62 - 7.08) | 6.88 (3.8 - 8.25) | 3.8 (3.37 - 4.23) |
| RBC ($10^6$/μl) | 33\|23\|3 | 4.87 (4.51 - 5.11) | 4.8 (4.66 - 5.15) | 4.76 (4.62 - 5.17) |
| Hgb (g/dl) | 33\|23\|3 | 13.1 (12.3 - 14.3) | 13.3 (12.8 - 13.55) | 13.2 (13.15 - 14.1) |
| Hct (%) | 33\|23\|3 | 41.8 (39.9 - 45.6) | 42.2 (41.15 - 44) | 43.9 (41.9 - 46.65) |
| MCV (fL) | 33\|23\|3 | 87.1 (82.5 – 92.7) | 89.7 (83.95 – 91.4) | 88.5 (86.2 – 93.3) |
| MCH (pg) | 33\|23\|3 | 27 (25.3 – 29) | 27.1 (26.1 – 28.8) | 27.5 (27.2 – 28.5) |
| MCHC (g/dl) | 33\|23\|3 | 30.8 (29.8 - 32) | 31.3 (29.85 - 32) | 30.4 (30.25 - 31.6) |
| Plt ($10^3$/μl) | 33\|23\|3 | 275 (226 - 307) | 217 (146 - 288) | 248 (218 - 286) |
| Neutrophils (%) | 33\|23\|3 | 52 (41.4 - 57.1) | 59.3 (25 - 65) | 46.7 (42.7 - 53.3) |
| Lymphocytes (%) | 33\|23\|3 | 31.5 (28 - 42) | 25.5 (21 - 37) | 31.7 (27.8 - 37.8) |
| Monocytes (%) | 33\|23\|3 | 8.2 (6.1 - 9.9) | 7 (5.15 - 9.75) | 13.3 (9.35 - 14.15) |
| Eosinophils (%) | 33\|22\|3 | 3.6 (2.2 - 6.7) | 2.05 (1.5 - 4.8) | 2.4 (1.95 - 7.1) |
| Basophils (%) | 33\|21\|3 | 1.8 (1.3 - 3.9) | 1.7 (1.2 - 6.3) | 1.7 (1.6 - 2.15) |
| | | **Binary Traits** | | |
| Angina | Cases: 2\|5\|0<br>Controls: 73\|45\|3 | 2 (2.7%) | 5 (10%) | 0 |
| Diabetes | Cases: 19\|16\|1<br>Controls: 56\|36\|2 | 19 (25.3%) | 16 (32%) | 1 (33.3%) |
| Hypertension | Cases: 20\|28\|3<br>Controls: 55\|22\|0 | 20 (26.7%) | 28 (56%) | 3 (100%) |
| MI | Cases: 3\|16\|3<br>Controls: 72\|34\|0 | 3 (4%) | 16 (32%) | 0 (0%) |
| MI age of Onset | Cases: 3\|16\|0 | 63 (56 - 69) | 52 (48 - 63) | NA |
| Stroke | Cases: 1\|1\|0<br>Controls: 74\|49\|3 | 1 (1.3%) | 1 (2%) | 0 |

1. Shown is a summary by genotype of the 128 callback participants, including 27 families of heterozygotes and 1 homozygotes family successfully contacted for a recall study and genotyped for Arg1231Cys. In total 2 additional homozygotes were identified, 50 heterozygous carriers and 75 non-carriers among the families. Baseline characteristics and self-reported medical history for all individuals is provided in the table. From left-to-right is the phenotype, the total genotype counts (RR =reference homozygote | RA = heterozygote | AA = alternate homozygote), and the median with interquartile range in parenthesis for RR, RA and AA genotypes. Quantitative traits on top and binary traits on the bottom. eGFR: estimated Glomerular Filtration Rate (Calculate from creatinine using MDRD equation, BMR: Base Metabolic Rate, FFM: Fat Free Mass, Hgb: Hemoglobin, Hct: Hematocrit, MCV: Mean Corpuscular Volume, MCH: , Mean Corpuscular Hemoglobin, MCHC: Mean Corpuscular Hemoglobin Concentration, Plt: Platelets, MI: Myocardial Infarction

**Supplementary Table 9. Comparison of *NOTCH3* p.Arg1231Cys Genotypes in PGR[1]**

| Phenotype | Genotype counts (RR\|RA\|AA) | Homozygous RR | Heterozygous RA | Homozygous AA | P val additive | P val Het RA vs Hom AA |
|---|---|---|---|---|---|---|
| Age | 74997\|813\|9 | 54 (47, 60) | 53 (47, 60) | 50 (45, 66) | | |
| Stroke All | Cases: 4939\|98\|3 Controls: 25619\|271\|2 | 16% | 27% | 60% | 4.92E-08 | 0.28 |
| Stroke Age | 4939\|98\|3 | 60 (50, 67) | 57 (49, 63) | 70 (68, 75) | | |
| Myocardial infarction | Cases: 26665\|272\|3 Controls: 33563\|361\|3 | 44% | 43% | 50% | 0.89 | 0.86 |
| Myocardial Infarction Age | 26665\|272\|3 | 55 (46, 60) | 52 (45, 60) | 45 (42.5, 47.5) | | |
| Hypertension (Self-Reported) | Cases: 27304\|294\|3 Controls: 46954\|508\|5 | 37% | 37% | 38% | 0.58 | 0.46 |
| Type 2 Diabetes | Cases: 21322\|221\|2 Controls: 37824\|410\|6 | 36% | 35% | 25% | 0.67 | 0.68 |
| Height (m) | 44680\|503\|5 | 165 (159, 170) | 165 (158, 170) | 167 (160, 170) | 0.75 | 0.82 |
| Weight (kg) | 45066\|507\|5 | 71 (65, 80) | 70 (64, 80) | 70 (69, 76) | 0.055 | 0.82 |
| BMI (kg/m$^2$) | 44125\|497\|5 | 26.45 (23.88, 29.3) | 26.22 (23.66, 29.38) | 26.95 (22.6, 27.25) | 0.125 | 0.97 |
| Waist to Hip Ratio | 43899\|501\|5 | 0.96 (0.92, 0.99) | 0.96 (0.92, 0.99) | 1.025 (0.96, 1.04) | 0.086 | 0.15 |
| Systolic BP | 63462\|692\|6 | 130 (120, 140) | 130 (120, 140) | 150 (127.5, 150) | 0.78 | 0.068 |
| Diastolic BP | 63992\|703\|6 | 80 (76, 90) | 80 (80, 90) | 95 (86.25, 100) | 0.19 | 0.016 |
| Total Cholesterol (mg/dl) | 70797\|765\|8 | 174 (143.1, 207.9) | 175.5 (142, 208) | 219.5 (162, 227) | 0.85 | 0.087 |
| Triglycerides (mg/dl) | 70617\|759\|8 | 156.75 (109.6, 228.8) | 153.0 (109.95, 235.0) | 190.75 (133.25, 264.65) | 0.72 | 0.74 |
| LDL (mg/dl) | 68503\|734\|8 | 103.5 (78.18, 131) | 104.57 (77.38, 129.35) | 123.14 (104.9, 137.25) | 0.59 | 0.16 |
| HDL (mg/dl) | 70535\|759\|8 | 34 (27.8, 41) | 34 (28, 41) | 31.9 (30.25, 35) | 0.30 | 0.76 |
| Creatinine (mg/dl) | 53244\|567\|7 | 0.9 (0.7, 1.08) | 0.9 (0.7, 1.08) | 0.9 (0.86, 1.05) | 0.38 | 0.56 |
| eGFR | 51084\|541\|7 | 86.75 (69.83, 104.66) | 87.5 (71.45, 105.59) | 93.5 (71.33, 94.51) | 0.68 | 0.89 |

1. Characteristics of non-carriers, heterozygotes and homozygotes of Arg1231Cys across 75,819 sequenced individuals in PGR. P-values were generated using whole genome regression as implemented in REGENIE adjusting for age, age$^2$, age*sex, sex and top 10 genetic PCs. Hypertension was self-reported. Type 2 Diabetes cases were defined as individuals self-reporting to have diabetes, or HbA1c > 6.5, or on oral hypoglycemic medication. eGFR was calculated using MDRD equation.

**Supplementary Table 10**. ***NOTCH3* p.Arg1231Cys Frequency Across 1000 Genomes Phase 3 Populations[1].**

| Primary Ancestral Component | Population code | Population Description | Variant | A1 | A2 | AAF | Allele count | Count A1 | Count A2 | Total People |
|---|---|---|---|---|---|---|---|---|---|---|
| Africa | ACB | African Caribbean in Barbados | 19:15179052:G:A | A | G | 0 | 192 | 0 | 192 | 96 |
| | ASW | African Ancestry in Southwest US | 19:15179052:G:A | A | G | 0 | 122 | 0 | 122 | 61 |
| | ESN | Esan in Nigeria | 19:15179052:G:A | A | G | 0 | 198 | 0 | 198 | 99 |
| | GWD | Gambian in Western Division, The Gambia - Mandinka | 19:15179052:G:A | A | G | 0 | 226 | 0 | 226 | 113 |
| | LWK | Luhya in Webuye, Kenya | 19:15179052:G:A | A | G | 0 | 198 | 0 | 198 | 99 |
| | MSL | Mende in Sierra Leone | 19:15179052:G:A | A | G | 0 | 170 | 0 | 170 | 85 |
| | YRI | Yoruba in Ibadan, Nigeria | 19:15179052:G:A | A | G | 0 | 216 | 0 | 216 | 108 |
| Americas | CLM | Colombian in Medellin, Colombia | 19:15179052:G:A | A | G | 0 | 188 | 0 | 188 | 94 |
| | MXL | Mexican Ancestry in Los Angeles, California | 19:15179052:G:A | A | G | 0 | 128 | 0 | 128 | 64 |
| | PEL | Peruvian in Lima, Peru | 19:15179052:G:A | A | G | 0 | 170 | 0 | 170 | 85 |
| | PUR | Puerto Rican in Puerto Rico | 19:15179052:G:A | A | G | 0 | 208 | 0 | 208 | 104 |
| East Asia | CDX | Chinese Dai in Xishuangbanna, China | 19:15179052:G:A | A | G | 0 | 186 | 0 | 186 | 93 |
| | CHB | Han Chinese in Beijing, China | 19:15179052:G:A | A | G | 0 | 206 | 0 | 206 | 103 |
| | CHS | Han Chinese South | 19:15179052:G:A | A | G | 0 | 210 | 0 | 210 | 105 |
| | JPT | Japanese in Tokyo, Japan | 19:15179052:G:A | A | G | 0 | 208 | 0 | 208 | 104 |
| | KHV | Kinh in Ho Chi Minh City, Vietnam | 19:15179052:G:A | A | G | 0 | 198 | 0 | 198 | 99 |
| Europe | CEU | Utah residents (CEPH) with Northern and Western European ancestry | 19:15179052:G:A | A | G | 0 | 198 | 0 | 198 | 99 |
| | FIN | Finnish in Finland | 19:15179052:G:A | A | G | 0 | 198 | 0 | 198 | 99 |
| | GBR | British in England and Scotland | 19:15179052:G:A | A | G | 0 | 182 | 0 | 182 | 91 |
| | IBS | Iberian populations in Spain | 19:15179052:G:A | A | G | 0 | 214 | 0 | 214 | 107 |
| | TSI | Toscani in Italy | 19:15179052:G:A | A | G | 0 | 214 | 0 | 214 | 107 |
| South Asia | BEB | Bengali in Bangladesh | 19:15179052:G:A | A | G | 0 | 172 | 0 | 172 | 86 |
| | GIH | Gujarati Indians in Houston, TX | 19:15179052:G:A | A | G | 0.0049 | 206 | 1 | 205 | 103 |

| | | | | | | | | | | |
|---|---|---|---|---|---|---|---|---|---|---|
| | ITU | Indian Telugu in the UK | 19:15179052:G:A | A | G | 0.0049 | 204 | 1 | 203 | 102 |
| | PJL | Punjabi in Lahore, Pakistan | 19:15179052:G:A | A | G | 0.0052 | 192 | 1 | 191 | 96 |
| | STU | Sri Lankan Tamil in the UK | 19:15179052:G:A | A | G | 0.0049 | 204 | 1 | 203 | 102 |

1. From left-to-right is the primary ancestral component of the population, the standard population code, description of the population, variant (chr:pos:ref:alt), alternate allele, reference allele, alternative allele frequency, allele count, alternate allele count, reference allele count.

**Supplementary Table 11**. ***NOTCH3* p.Arg1231Cys Frequency Across Pakistani Ethnic Groups[1].**

| Population | Name | A1 | A2 | AAF | Allele count | Count A1 | Count A2 | Total Alleles | Total People | P | Q | $P^2$ | 2PQ | $Q^2$ | n (Het) | n (Hom) | n (Total) | Prevalence (1 in ?) |
|---|---|---|---|---|---|---|---|---|---|---|---|---|---|---|---|---|---|---|
| Ansari | 19:15179052:G:A | A | G | 5.60E-03 | 20,804 | 118 | 20,686 | 20,804 | 10,402 | 9.94E-01 | 5.60E-03 | 9.89E-01 | 1.11E-02 | 3.14E-05 | 116 | 0 | 116 | 90 |
| Rajput/ Punjabi | 19:15179052:G:A | A | G | 6.00E-03 | 41,814 | 252 | 41,562 | 41,814 | 20,907 | 9.94E-01 | 6.00E-03 | 9.88E-01 | 1.19E-02 | 3.60E-05 | 249 | 1 | 250 | 84 |
| Balochi/ Pathan | 19:15179052:G:A | A | G | 7.40E-03 | 6,136 | 46 | 6,090 | 6,136 | 3,068 | 9.93E-01 | 7.40E-03 | 9.85E-01 | 1.47E-02 | 5.48E-05 | 45 | 0 | 45 | 68 |

1. To assess the prevalence of p.Arg1231Cys across ethnic groups, the first seven principal components (PCs) were quantified from Illumina GSA-V2 array data for a sample of PGR where self-reported ethnicity was available, and a UMAP plot [9] was produced from the data. Genetic data for the rest of the PGR cohort was projected into this space. Visual inspection of the plot identified three major ethnic groups, including Ansari (South Asian), Rajput/ Punjabi (South Asian), and Balochi/ Pathan (West Asian). Allele frequency within each ethnic group was calculated from exome sequence data. From left-to-right is the ethnicity, chromosome, variant (chr:pos:ref:alt), alternate allele, reference allele, alternate allele frequency, allele count, alternate allele count, reference allele count.

**Supplementary Table 12. Significant and Suggestive ExWAS Hits in PGR Discovery Cohort Across 5 Stroke Phenotypes[1].**

| Category[2] | Variant | Nearest Gene | DbSNP rsID | Phenotype | Odds ratio (95% CI) | P value | AAF | Cases (RR\|RA\|AA) | Controls (RR\|RA\|AA) |
|---|---|---|---|---|---|---|---|---|---|
| Suggestive | 3:33152905:A:C | *SUSD5* | rs768145924 | Ischemic stroke | 143.28 (20.11, 1020.7) | 7.20E-07 | 2.28E-04 | 2578\|6\|0 | 25885\|7\|0 |
| Suggestive | 6:73615308:C:T | *SLC17A5* | rs146729568 | Intracerebral hemorrhage | 2.29 (1.65, 3.17) | 7.29E-07 | 1.03E-02 | 1442\|50\|3 | 25393\|489\|9 |
| Suggestive | 6:73615308:C:T | *SLC17A5* | rs146729568 | Subcortical intracerebral hemorrhage | 2.35 (1.67, 3.3) | 8.61E-07 | 1.03E-02 | 1306\|48\|2 | 25393\|489\|9 |
| Suggestive | 11:114527797:G:A | *NXPE1* | rs778206967 | Subcortical intracerebral hemorrhage | 23.47 (6.66, 82.65) | 9.00E-07 | 4.61E-04 | 1346\|8\|0 | 25754\|17\|0 |
| Suggestive* | 11:1884062:T:C | *LSP1* | rs661348 | Intracerebral hemorrhage | 1.26 (1.16, 1.37) | 8.01E-08 | 2.74E-01 | 711\|610\|173 | 13951\|9824\|2096 |
| Suggestive* | 11:1884062:T:C | *LSP1* | rs661348 | Subcortical intracerebral hemorrhage | 1.27 (1.16, 1.38) | 9.67E-08 | 2.73E-01 | 637\|560\|158 | 13951\|9824\|2096 |
| Significant | 19:15179052:G:A | *NOTCH3* | rs201680145 | Subcortical intracerebral hemorrhage | 3.39 (2.26, 5.1) | 3.87E-09 | 5.84E-03 | 1313\|43\|0 | 25619\|271\|2 |
| Significant | 19:15179052:G:A | *NOTCH3* | rs201680145 | Intracerebral hemorrhage | 3.17 (2.12, 4.73) | 1.79E-08 | 5.82E-03 | 1451\|44\|0 | 25619\|271\|2 |
| Significant | 19:15179052:G:A | *NOTCH3* | rs201680145 | Stroke | 2.19 (1.65, 2.9) | 4.92E-08 | 6.13E-03 | 4939\|98\|3 | 25619\|271\|2 |

1. Shown are the genome-wide significant (p value < $5.0\times10^{-8}$) and suggestive hits (p value < $1.0\times10^{-6}$ & > $5.0\times10^{-8}$) for 5 stroke phenotype ExWAS in the PGR discovery cohort with at least 10 variant carriers. From left-to-right is the significance category (significant or suggestive), variant name (Chr:pos:ref:alt), nearest gene, dbSNP rsID, phenotype, effect size, odds ratio (95% confidence interval), p value, cases count (total | homozygous reference | heterozygous |homozygous alternate), controls count (total | homozygous reference | heterozygous |homozygous alternate). Hits are organized from top-to-bottom by genomic coordinates and effect size.
2. Known locus from recent major GWAS [10] denoted with "*"
3. Stroke subphenotypes with <500 cases were not analyzed.

**Supplementary Table 13. Conditional Association Test of *LSP1* Locus Variant Associated with Stroke in PGR[1].**

| Analysis[2] | Variant | Nearest Gene | DbSNP rsID | Phenotype | Odds ratio (95% CI) | P value | AAF | Cases (RR\|RA\|AA) | Controls (RR\|RA\|AA) |
|---|---|---|---|---|---|---|---|---|---|
| Initial | 11:1884062:T:C | LSP1 | rs661348 | Intracerebral hemorrhage | 1.26 (1.16, 1.37) | 8.01E-08 | 2.74E-01 | 711\|610\|173 | 13951\|9824\|2096 |
| Initial | 11:1884062:T:C | LSP1 | rs661348 | Subcortical intracerebral hemorrhage | 1.27 (1.16, 1.38) | 9.67E-08 | 2.73E-01 | 711\|610\|173 | 13951\|9824\|2096 |
| Conditional | 11:1884062:T:C | LSP1 | rs661348 | Intracerebral hemorrhage | 1.23 (1.09, 1.39) | 8.83E-04 | 2.74E-01 | 684\|596\|168 | 13139\|9260\|1988 |
| Conditional | 11:1884062:T:C | LSP1 | rs661348 | Subcortical intracerebral hemorrhage | 1.22 (1.08, 1.39) | 2.06E-03 | 2.74E-01 | 613\|546\|153 | 13139\|9260\|1988 |

1. Shown are the top hits ($p < 1.0x10^{-7}$) for 5 stroke Phenotype ExWAS in the PGR discovery cohort in addition to *NOTCH3* p.Arg1231Cys. From left-to-right is the stage in the analysis (initial or conditional), variant name (Chr:pos:ref:alt), nearest gene, dbSNP rsID, phenotype, effect size, odds ratio (95% confidence interval), p value, cases count (total | homozygous reference | heterozygous |homozygous alternate), controls count (total | homozygous reference | heterozygous |homozygous alternate).
2. Initial analysis revealed an association between LSP intronic variant rs661348, a locus where two other non-coding variants (rs1973765 and rs569550) were previously associated with stroke in a large multi-ethnic GWAS [10]. A conditional analysis was conducted, conditional on the two known variants.

**Supplementary Table 14. PheWAS of Binary Traits in PGR Discovery Cohort for *LSP1* rs661348[1].**

| Phenotype | Odds ratio (95% CI) | P value | AAF | Cases (RR\|RA\|AA) | Controls (RR\|RA\|AA) |
|---|---|---|---|---|---|
| Intracerebral hemorrhage diagnosis | 1.3 (1.2, 1.4) | 8.01E-08 | 0.27 | 711\|610\|173 | 13951\|9824\|2096 |
| Subcortical intracerebral hemorrhage | 1.3 (1.2, 1.4) | 9.67E-08 | 0.27 | 637\|560\|158 | 13951\|9824\|2096 |
| Stroke | 1.1 (1, 1.2) | 2.94E-04 | 0.27 | 2579\|1990\|467 | 13951\|9824\|2096 |
| Using combination drug treatment for high blood pressure / hypertension | 1.3 (1.1, 1.5) | 1.50E-03 | 0.28 | 185\|171\|45 | 17763\|12728\|2795 |
| New pathological Q waves | 1.2 (1.1, 1.4) | 1.65E-03 | 0.28 | 295\|254\|64 | 9520\|6743\|1493 |
| Diabetes mellitus case vs control | 0.94 (0.91, 0.98) | 4.10E-03 | 0.28 | 4681\|3409\|648 | 14692\|10487\|2394 |
| Self-reported history of diabetes | 0.95 (0.91, 0.98) | 5.41E-03 | 0.28 | 4665\|3401\|648 | 14703\|10490\|2393 |
| Increase or irregular heartbeat | 0.94 (0.9, 0.99) | 1.01E-02 | 0.27 | 2951\|2050\|436 | 16331\|11766\|2585 |
| Self-reported hypertension | 1 (1, 1.1) | 2.61E-02 | 0.28 | 7924\|5886\|1308 | 11335\|7937\|1713 |
| Physical activity at work, light | 0.94 (0.89, 1) | 3.61E-02 | 0.27 | 2056\|1434\|271 | 17587\|12641\|2809 |
| Digoxin medication for heart failure | 1.2 (1, 1.5) | 3.64E-02 | 0.28 | 93\|108\|17 | 17851\|12789\|2822 |

1. Shown are ExWAS summary statistics for available phenotypes with p values < 0.05 in the PGR discovery cohort. From left to right is the phenotype, odds ratio (95% confidence interval in parenthesis), p value, allele frequency, cases genotype counts (Ref homozygote | heterozygote | Alt homozygote), controls genotype counts (Ref homozygote | heterozygote | Alt homozygote).
2. Three analyses were conducted, with the main difference being how the controls were defined. For more detail see Supplementary Text for details.

**Supplementary Table 15. PheWAS of Quantitative Traits in PGR Discovery Cohort for *LSP1* rs661348[1].**

| Phenotype | Beta (95% CI) | P value | AAF | Cases (RR\|RA\|AA) | Units |
|---|---|---|---|---|---|
| Age at diagnosis of angina | 0.12 (0.021, 0.21) | 1.68E-02 | 0.27 | 479\|336\|71 | Years |
| Oral hypoglycemics I duration | -0.13 (-0.24, -0.021) | 1.96E-02 | 0.25 | 309\|203\|38 | Years |
| Low-density lipoprotein (LDL) calculated | -0.017 (-0.034, -0.0014) | 3.35E-02 | 0.27 | 18241\|13021\|2856 | mmHg |
| Aspirin medication duration | -0.071 (-0.14, -0.0045) | 3.65E-02 | 0.28 | 842\|640\|139 | Years |
| Waist hip ratio | 0.017 (0.00082, 0.034) | 3.96E-02 | 0.27 | 17301\|12358\|2671 | Decimal |

1. Shown are ExWAS summary statistics for available phenotypes with p values < 0.05 in the PGR discovery cohort. From left to right is the phenotype, effect size (95% confidence interval in parenthesis), p value, allele frequency, cases genotype counts (Ref homozygote | heterozygote | Alt homozygote ), and units.

**Supplementary Table 16. Top PheWAS Binary Trait Hits in UKB Replication Cohort for *LSP1* rs661348[1].**

| Phenotype | Odds Ratio (95% CI) | P value | AAF | Cases (RR\|RA\|AA) | Controls (RR\|RA\|AA) |
|---|---|---|---|---|---|
| Hypertension, self-reported - any visit | 1.1 (1, 1.1) | 8.05E-25 | 0.4 | 37969\|56217\|21192 | 74251\|104598\|37317 |
| RGC BP med | 1.1 (1, 1.1) | 1.45E-24 | 0.4 | 32816\|48543\|18397 | 75843\|106627\|38331 |
| I10: Essential (primary) hypertension | 1.1 (1, 1.1) | 2.08E-23 | 0.4 | 34666\|51130\|19254 | 92277\|130085\|46561 |
| Hypertension, ICD10: I10 or self-reported - any visit | 1.1 (1, 1.1) | 7.40E-18 | 0.4 | 33020\|48785\|18340 | 78279\|110745\|39672 |
| Bendroflumethiazide - any visit | 1.1 (1.1, 1.1) | 8.92E-16 | 0.4 | 7761\|12180\|4624 | 100898\|142990\|52104 |
| Amlodipine - any visit | 1.1 (1.1, 1.1) | 2.90E-15 | 0.4 | 6281\|9592\|3809 | 102378\|145578\|52919 |
| Medication for cholesterol, blood pressure or diabetes - any visit | 1.1 (1.1, 1.1) | 3.23E-13 | 0.4 | 17058\|24841\|9501 | 6371\|8741\|2917 |

1. To investigate the potential association between *LSP1* and stroke, a PheWAS analysis was conducted across UKB phenotypes for the associated variant rs661348. Shown are phenotypes with genome-wide significant p values, from left-to-right is the phenotype, odds ratio (95% confidence interval), p value, alternate allele frequency (AAF), case counts by genotype (Ref homozygote | heterozygote | Alt homozygote), and the control counts by genotype (Ref homozygote | heterozygote | Alt homozygote).

**Supplementary Table 17. Top PheWAS Quantitative Trait Hits in UKB Replication Cohort for *LSP1* rs661348[1].**

| Phenotype | Beta (95% CI) | Pval | AAF | Cases (RR\|RA\|AA) | Units |
|---|---|---|---|---|---|
| Systolic blood pressure automated reading - initial visit - mean | 0.021 (0.017, 0.025) | 6.91E-25 | 0.4 | 137851\|196508\|71709 | mmHg |
| Systolic blood pressure automated reading - mean of all visits | 0.021 (0.017, 0.025) | 6.94E-25 | 0.4 | 138486\|197438\|72035 | mmHg |

1. To investigate the potential association between *LSP1* and stroke, a PheWAS analysis was conducted across UKB phenotypes for the associated variant rs661348. Shown are phenotypes with genome-wide significant p values, from left-to-right is the phenotype, effect size (95% confidence interval), p value, alternate allele frequency (AAF), case counts by genotype (Ref homozygote | heterozygote | Alt homozygote), and the units.

**Supplemental Table 18. Comparison of *NOTCH3* p.Arg1231Cys Allele Frequency Across Ancestries in UKB[1].**

| Ancestry | Field | Value |
|---|---|---|
| Total | AAF | 0.00028 |
| | HET RA | 255 |
| | HOM AA | 0 |
| | N | 454,787 |
| African / African American | AAF | 0 |
| | HET RA | 0 |
| | HOM AA | 0 |
| | N | 9,093 |
| Admixed American | AAF | 0.000548 |
| | HET RA | 1 |
| | HOM AA | 0 |
| | N | 912 |
| European / European American | AAF | 0.000191 |
| | HET RA | 165 |
| | HOM AA | 0 |
| | N | 431,384 |
| East Asian | AAF | 0 |
| | HET RA | 0 |
| | HOM AA | 0 |
| | N | 2254 |
| South Asian | AAF | 0.004377 |
| | HET RA | 87 |
| | HOM AA | 0 |
| | N | 9,939 |

1. Shown are the genotype counts and allele frequencies across ancestries in UKB. From left-to-right is the ancestry, field (top-to-bottom: alternate allele frequency, heterozygote genotype count, homozygote genotype count, total genotype count), and the value.

**Supplementary Table 19**. **PheWAS of Binary Trait Associations with p.Arg1231Cys in 450 K UKB European Participants[1]**.

| Phenotype | P value | Odds ratio (CI) | AAF | Cases RR\|RA\|AA | Controls RR\|RA\|AA |
|---|---|---|---|---|---|
| Coffee type - any visit - Instant coffee AnyInst | 6.1E-06 | 2.4 (1.6, 3.5) | 1.9E-04 | 192971\|97\|0 | 148099\|32\|0 |
| Satisfaction with bowel habits - any visit - 10 Very unhappy AnyInst | 3.4E-05 | 6.2 (2.6, 15) | 1.8E-04 | 4977\|9\|0 | 149254\|48\|0 |
| Average total household income before tax - any visit - Less than 18 000 AnyInst | 3.9E-05 | 2.1 (1.5, 3.1) | 2.0E-04 | 85728\|56\|0 | 286961\|91\|0 |
| Qualifications - any visit - College or University degree AnyInst | 7.6E-05 | 0.46 (0.31, 0.67) | 1.9E-04 | 140274\|33\|0 | 213035\|99\|0 |
| Coffee type - any visit - Ground coffee include espresso filter etc AnyInst | 9.4E-05 | 0.39 (0.24, 0.63) | 1.9E-04 | 84032\|16\|0 | 257038\|113\|0 |
| Frequency of drinking alcohol - any visit - Monthly or less AnyInst | 2.9E-04 | 3.3 (1.7, 6.3) | 1.9E-04 | 17953\|16\|0 | 121426\|36\|0 |
| Mixed vegetable intake - any visit - half AnyInst | 3.3E-04 | 6.9 (2.4, 20) | 2.4E-04 | 4751\|8\|0 | 24753\|6\|0 |
| RGC_CVD_Ischemic_Composite_noAfib_CC[2] | 4.1E-04 | 4 (1.9, 8.6) | 2.0E-04 | 6873\|10\|0 | 353143\|134\|0 |
| I67: Other cerebrovascular diseases | 4.2E-04 | 5.3 (2.1, 14) | 2.0E-04 | 3810\|7\|0 | 382830\|148\|0 |
| Illnesses of mother - any visit - 27 None of the above group 2 AnyInst | 4.3E-04 | 0.57 (0.41, 0.78) | 2.0E-04 | 308171\|103\|0 | 115021\|67\|0 |
| O47: False labor | 4.4E-04 | 18 (3.6, 86) | 1.9E-04 | 1128\|3\|0 | 210633\|77\|0 |
| OPCS Z586 | 4.7E-04 | 15 (3.3, 70) | 2.0E-04 | 681\|3\|0 | 350984\|141\|0 |
| Y83.4: Other reconstructive surgery as the cause of abnormal reaction of the patient, or of later complication, without mention of misadventure at the time of the procedure | 5.2E-04 | 9.5 (2.7, 34) | 2.0E-04 | 1102\|4\|0 | 386383\|151\|0 |
| Illnesses of mother - any visit - Parkinsons disease AnyInst | 5.4E-04 | 4.1 (1.8, 9.1) | 2.0E-04 | 6610\|9\|0 | 416582\|161\|0 |
| Illnesses of siblings - any visit - Stroke AnyInst | 6.3E-04 | 3 (1.6, 5.7) | 2.0E-04 | 12230\|14\|0 | 359062\|138\|0 |
| OPCS4 Y39.5: Tattooing of organ NOC | 6.3E-04 | 7.1 (2.3, 22) | 2.0E-04 | 1790\|5\|0 | 349875\|139\|0 |
| Ever taken oral contraceptive pill - any visit - Yes AnyInst | 7.1E-04 | 0.41 (0.25, 0.69) | 1.9E-04 | 191340\|60\|0 | 41534\|27\|0 |
| RGC_CVD_Ischemic_Composite_CC | 8.8E-04 | 3.4 (1.6, 6.9) | 2.0E-04 | 9124\|11\|0 | 370986\|139\|0 |
| Ever taken oral contraceptive pill - any visit - No AnyInst | 1.1E-03 | 2.3 (1.4, 3.9) | 1.9E-04 | 42344\|27\|0 | 190530\|60\|0 |
| Worked with materials containing asbestos - any visit - 131 Sometimes AnyInst | 1.1E-03 | 4.1 (1.8, 9.7) | 2.0E-04 | 9782\|11\|0 | 81098\|25\|0 |
| Never eat eggs dairy wheat sugar - any visit - Sugar or foods drinks containing sugar AnyInst | 1.6E-03 | 0.49 (0.32, 0.77) | 2.0E-04 | 86431\|19\|0 | 342735\|152\|0 |
| Frequency of drinking alcohol - any visit - 2 to 3 times a week AnyInst | 1.8E-03 | 0.32 (0.15, 0.65) | 1.9E-04 | 41933\|6\|0 | 97446\|46\|0 |
| White wine intake - any visit - 6 AnyInst | 2.1E-03 | 19 (2.9, 130) | 1.8E-04 | 517\|2\|0 | 43564\|14\|0 |
| O47.1: False labor at or after 37 completed weeks of gestation | 2.5E-03 | 19 (2.8, 130) | 1.9E-04 | 698\|2\|0 | 228188\|84\|0 |
| Z88.0: Allergy status to penicillin | 2.6E-03 | 0.17 (0.053, 0.54) | 2.0E-04 | 20214\|1\|0 | 365382\|154\|0 |

| Phenotype | P value | Odds ratio (CI) | AAF | Cases RR\|RA\|AA | Controls RR\|RA\|AA |
|---|---|---|---|---|---|
| Red wine intake - any visit - 3 AnyInst | 3.0E-03 | 4.1 (1.6, 10) | 1.9E-04 | 9370\|9\|0 | 45414\|12\|0 |
| Bipolar and major depression status - any visit - Probable Recurrent major depression moderate AnyInst[2] | 3.1E-03 | 3.2 (1.5, 6.8) | 1.8E-04 | 13097\|12\|0 | 90619\|26\|0 |
| Work hours lumped category - any visit - 3040 30 to 40 hours AnyInst | 3.1E-03 | 0.37 (0.19, 0.71) | 2.0E-04 | 61493\|16\|0 | 27105\|20\|0 |
| Bread roll intake - any visit - 2 AnyInst | 3.3E-03 | 3.4 (1.5, 7.5) | 2.5E-04 | 13845\|13\|0 | 35956\|12\|0 |
| E28: Ovarian dysfunction | 3.5E-03 | 15 (2.4, 89) | 1.8E-04 | 497\|2\|0 | 210330\|76\|0 |
| K63.3: Ulcer of intestine | 3.5E-03 | 8.3 (2, 34) | 2.0E-04 | 926\|3\|0 | 386534\|152\|0 |
| Cereal type - any visit - Muesli AnyInst | 3.6E-03 | 0.5 (0.31, 0.8) | 1.9E-04 | 77762\|17\|0 | 281457\|122\|0 |
| Satsuma intake - any visit - 2 AnyInst | 3.7E-03 | 7.1 (1.9, 27) | 1.0E-04 | 14478\|7\|0 | 29319\|2\|0 |
| Grapefruit juice intake - any visit - half AnyInst | 4.2E-03 | 0.04 (0.0044, 0.36) | 3.0E-04 | 3779\|0\|0 | 4549\|5\|0 |
| H02.1: Ectropion of eyelid | 4.3E-03 | 13 (2.2, 76) | 2.0E-04 | 579\|2\|0 | 386906\|153\|0 |
| Illnesses of mother - any visit - Severe depression AnyInst | 4.5E-03 | 2.1 (1.3, 3.5) | 2.0E-04 | 26924\|20\|0 | 396268\|150\|0 |
| L03: Cellulitis and acute lymphangitis | 4.6E-03 | 2.6 (1.3, 5) | 2.0E-04 | 12954\|12\|0 | 360629\|137\|0 |
| Z88: Allergy status to drugs, medicaments and biological substances | 4.8E-03 | 0.35 (0.17, 0.73) | 2.0E-04 | 36306\|5\|0 | 339034\|146\|0 |
| FI3 word interpolation - any visit - Grow AnyInst | 5.0E-03 | 2.4 (1.3, 4.3) | 2.1E-04 | 18366\|17\|0 | 152749\|55\|0 |
| Direct bilirubin aliquot - any visit - 4 AnyInst | 5.3E-03 | 12 (2.1, 70) | 2.0E-04 | 533\|2\|0 | 413906\|164\|0 |
| Month of attending assessment centre - any visit - April AnyInst | 5.4E-03 | 0.41 (0.21, 0.77) | 2.0E-04 | 42560\|7\|0 | 387939\|165\|0 |
| Z34.9: Encounter for supervision of normal pregnancy, unspecified | 5.4E-03 | 13 (2.1, 81) | 1.9E-04 | 808\|2\|0 | 210591\|78\|0 |
| Alcohol intake frequency - any visit - Special occasions only AnyInst | 5.4E-03 | 1.8 (1.2, 2.7) | 2.0E-04 | 49429\|33\|0 | 380488\|139\|0 |
| OPCS4 W15.7: Osteotomy of bone of foot and fixation HFQ | 5.9E-03 | 7.2 (1.8, 29) | 2.0E-04 | 1048\|3\|0 | 350617\|141\|0 |
| Volume level set by participant right - any visit - 100 max AnyInst | 6.0E-03 | 2.8 (1.3, 6) | 2.1E-04 | 8693\|10\|0 | 164367\|63\|0 |
| evening primrose oil product - any visit | 6.1E-03 | 7.1 (1.8, 29) | 2.0E-04 | 988\|3\|0 | 319398\|128\|0 |
| FI3 word interpolation - any visit - Adult AnyInst | 6.1E-03 | 0.46 (0.26, 0.8) | 2.1E-04 | 146971\|52\|0 | 24144\|20\|0 |
| B37.0: Candidal stomatitis | 6.6E-03 | 6.8 (1.7, 27) | 2.0E-04 | 1307\|3\|0 | 386178\|152\|0 |
| Alkaline phosphatase aliquot - any visit - 4 AnyInst | 6.8E-03 | 11 (1.9, 62) | 2.0E-04 | 581\|2\|0 | 414252\|164\|0 |
| OPCS4 M76.4: Endoscopic dilation of urethra | 6.8E-03 | 5.2 (1.6, 17) | 2.0E-04 | 2127\|4\|0 | 349538\|140\|0 |
| OPCS4 W83.8: Other specified therapeutic endoscopic operations on other articular cartilage | 6.9E-03 | 11 (1.9, 61) | 2.0E-04 | 611\|2\|0 | 351054\|142\|0 |
| 3mm asymmetry index irregular astigmatism level R - any visit - doubtful AnyInst | 7.0E-03 | 5.3 (1.6, 18) | 2.1E-04 | 2427\|4\|0 | 99023\|38\|0 |

| Phenotype | P value | Odds ratio (CI) | AAF | Cases RR\|RA\|AA | Controls RR\|RA\|AA |
|---|---|---|---|---|---|
| Non oily fish intake - any visit - Less than once a week AnyInst | 7.2E-03 | 1.5 (1.1, 2.1) | 2.0E-04 | 131016\|69\|0 | 297606\|101\|0 |
| R59.0: Localized enlarged lymph nodes | 7.4E-03 | 5 (1.5, 16) | 2.0E-04 | 2462\|4\|0 | 383593\|151\|0 |
| I63.9: Cerebral infarction, unspecified | 7.6E-03 | 4.2 (1.5, 12) | 2.0E-04 | 3379\|5\|0 | 383680\|150\|0 |
| gabapentin - any visit | 7.7E-03 | 5 (1.5, 16) | 2.0E-04 | 2235\|4\|0 | 318151\|127\|0 |
| UK Biobank assessment centre - any visit - Newcastle AnyInst | 7.8E-03 | 2.2 (1.2, 4.1) | 2.0E-04 | 33212\|27\|0 | 397287\|145\|0 |
| Unspecified place | 7.8E-03 | 4.1 (1.5, 12) | 2.0E-04 | 3884\|5\|0 | 383601\|150\|0 |
| Cholesterol aliquot - any visit - 4 AnyInst | 7.8E-03 | 10 (1.8, 58) | 2.0E-04 | 601\|2\|0 | 414213\|164\|0 |
| Albumin aliquot - any visit - 4 AnyInst | 7.9E-03 | 10 (1.8, 58) | 2.0E-04 | 600\|2\|0 | 414195\|164\|0 |
| Urate aliquot - any visit - 4 AnyInst | 8.0E-03 | 10 (1.8, 57) | 2.0E-04 | 598\|2\|0 | 413822\|164\|0 |
| Vitamin D aliquot - any visit - 4 AnyInst | 8.1E-03 | 10 (1.8, 57) | 2.0E-04 | 599\|2\|0 | 414036\|164\|0 |
| Someone to take to doctor when needed as a child - any visit - Very often true AnyInst | 8.2E-03 | 0.43 (0.23, 0.8) | 1.9E-04 | 116508\|36\|0 | 22120\|16\|0 |
| Rheumatoid factor reportability - any visit - Reportable at assay and after aliquot correction if attempted AnyInst | 8.2E-03 | 1.9 (1.2, 2.9) | 2.0E-04 | 36920\|25\|0 | 374621\|137\|0 |
| Glucose aliquot - any visit - 4 AnyInst | 8.2E-03 | 10 (1.8, 56) | 2.0E-04 | 602\|2\|0 | 413613\|163\|0 |
| IGF 1 aliquot - any visit - 4 AnyInst | 8.2E-03 | 10 (1.8, 57) | 2.0E-04 | 603\|2\|0 | 412008\|162\|0 |
| Alanine aminotransferase aliquot - any visit - 4 AnyInst | 8.2E-03 | 10 (1.8, 56) | 2.0E-04 | 603\|2\|0 | 414285\|164\|0 |
| Cystatin C aliquot - any visit - 4 AnyInst | 8.2E-03 | 10 (1.8, 56) | 2.0E-04 | 603\|2\|0 | 414131\|164\|0 |
| Apolipoprotein B aliquot - any visit - 4 AnyInst | 8.2E-03 | 10 (1.8, 56) | 2.0E-04 | 602\|2\|0 | 414058\|164\|0 |
| Aspartate aminotransferase aliquot - any visit - 4 AnyInst | 8.2E-03 | 10 (1.8, 56) | 2.0E-04 | 603\|2\|0 | 414266\|164\|0 |
| Gamma glutamyltransferase aliquot - any visit - 4 AnyInst | 8.2E-03 | 10 (1.8, 56) | 2.0E-04 | 603\|2\|0 | 414119\|164\|0 |
| HDL cholesterol aliquot - any visit - 4 AnyInst | 8.2E-03 | 10 (1.8, 56) | 2.0E-04 | 603\|2\|0 | 414068\|164\|0 |
| Calcium aliquot - any visit - 4 AnyInst | 8.2E-03 | 10 (1.8, 56) | 2.0E-04 | 603\|2\|0 | 413827\|164\|0 |
| Rubella german measles, ICD10: B06 or Self-reported - any visit | 8.3E-03 | 4.9 (1.5, 16) | 2.1E-04 | 1956\|4\|0 | 326710\|133\|0 |
| C reactive protein aliquot - any visit - 4 AnyInst | 8.3E-03 | 10 (1.8, 56) | 2.0E-04 | 602\|2\|0 | 413866\|164\|0 |
| LDL direct aliquot - any visit - 4 AnyInst | 8.3E-03 | 10 (1.8, 56) | 2.0E-04 | 603\|2\|0 | 413576\|164\|0 |
| Rheumatoid factor aliquot - any visit - 4 AnyInst | 8.3E-03 | 10 (1.8, 56) | 2.0E-04 | 602\|2\|0 | 413525\|162\|0 |
| Phosphate aliquot - any visit - 4 AnyInst | 8.3E-03 | 10 (1.8, 56) | 2.0E-04 | 603\|2\|0 | 413412\|164\|0 |
| Apolipoprotein A aliquot - any visit - 4 AnyInst | 8.3E-03 | 10 (1.8, 56) | 2.0E-04 | 603\|2\|0 | 413613\|164\|0 |

| Phenotype | P value | Odds ratio (CI) | AAF | Cases RR\|RA\|AA | Controls RR\|RA\|AA |
|---|---|---|---|---|---|
| Lipoprotein A aliquot - any visit - 4 AnyInst | 8.3E-03 | 10 (1.8, 56) | 2.0E-04 | 601\|2\|0 | 401676\|159\|0 |
| Triglycerides aliquot - any visit - 4 AnyInst | 8.5E-03 | 10 (1.8, 56) | 2.0E-04 | 593\|2\|0 | 413974\|164\|0 |
| Creatinine aliquot - any visit - 4 AnyInst | 8.6E-03 | 10 (1.8, 55) | 2.0E-04 | 601\|2\|0 | 414001\|164\|0 |
| Total bilirubin aliquot - any visit - 4 AnyInst | 8.6E-03 | 10 (1.8, 55) | 2.0E-04 | 602\|2\|0 | 413955\|164\|0 |
| Urea aliquot - any visit - 4 AnyInst | 8.6E-03 | 10 (1.8, 55) | 2.0E-04 | 602\|2\|0 | 413990\|164\|0 |
| Detention categories - any visit - Informal not formally detained AnyInst | 8.8E-03 | 0.13 (0.029, 0.6) | 1.9E-04 | 34076\|11\|0 | 2285\|3\|0 |
| N90.7: Vulvar cyst | 9.0E-03 | 10 (1.8, 57) | 1.9E-04 | 570\|2\|0 | 228316\|84\|0 |
| Willing to attempt cognitive tests - any visit - Begin games AnyInst | 9.9E-03 | 0.32 (0.13, 0.76) | 2.0E-04 | 424502\|165\|0 | 5738\|7\|0 |
| Someone to take to doctor when needed as a child - any visit - Sometimes true AnyInst | 1.0E-02 | 4.1 (1.4, 12) | 1.9E-04 | 3655\|5\|0 | 134973\|47\|0 |
| Intake of sugar added to coffee - any visit - half AnyInst | 1.0E-02 | 0.17 (0.045, 0.66) | 2.7E-04 | 10906\|1\|0 | 20643\|16\|0 |
| R93.3: Abnormal findings on diagnostic imaging of other parts of digestive tract | 1.1E-02 | 4.6 (1.4, 15) | 2.0E-04 | 2206\|4\|0 | 382708\|150\|0 |
| Death report format - any visit - IC Death Format 2012 onwards AnyInst | 1.1E-02 | 0.12 (0.023, 0.61) | 2.0E-04 | 13517\|2\|0 | 4018\|5\|0 |
| K61.0: Anal abscess | 1.1E-02 | 5.9 (1.5, 23) | 2.0E-04 | 1289\|3\|0 | 386192\|152\|0 |
| RGC_T2D_Nephropathy_Loose_CC | 1.1E-02 | 11 (1.8, 72) | 1.2E-04 | 2064\|2\|0 | 23526\|4\|0 |
| L29.0: Pruritus ani | 1.1E-02 | 5.8 (1.5, 23) | 2.0E-04 | 1389\|3\|0 | 383387\|151\|0 |
| Bipolar and major depression status - any visit - No Bipolar or Depression AnyInst | 1.1E-02 | 0.43 (0.22, 0.82) | 1.8E-04 | 74794\|19\|0 | 28922\|19\|0 |
| M16.1: Unilateral primary osteoarthritis of hip | 1.1E-02 | 3.9 (1.4, 11) | 2.0E-04 | 3950\|5\|0 | 383535\|150\|0 |
| Duration of heavy DIY - any visit - Between 15 and 30 minutes AnyInst | 1.1E-02 | 2 (1.2, 3.4) | 1.9E-04 | 33565\|21\|0 | 145055\|47\|0 |
| M79.2: Neuralgia and neuritis, unspecified | 1.2E-02 | 8.8 (1.6, 48) | 2.0E-04 | 583\|2\|0 | 386529\|153\|0 |
| Sexually molested as a child - any visit - Often AnyInst | 1.2E-02 | 9 (1.6, 50) | 1.9E-04 | 671\|2\|0 | 137322\|50\|0 |
| Z03.4: Observation for suspected myocardial infarction | 1.2E-02 | 4.4 (1.4, 14) | 2.0E-04 | 2719\|4\|0 | 384766\|151\|0 |
| OPCS4 H55.3: Laying open of anal fistula NEC | 1.3E-02 | 8.5 (1.6, 46) | 2.0E-04 | 670\|2\|0 | 350995\|142\|0 |
| Number of vehicles in household - any visit - None AnyInst | 1.3E-02 | 1.8 (1.1, 2.9) | 2.0E-04 | 35285\|24\|0 | 392302\|148\|0 |
| General happiness with own health - any visit - Extremely unhappy AnyInst | 1.3E-02 | 5.7 (1.4, 23) | 1.9E-04 | 1672\|3\|0 | 137526\|49\|0 |
| Plum intake - any visit - 2 AnyInst | 1.3E-02 | 0.088 (0.013, 0.6) | 3.4E-04 | 5311\|0\|0 | 12542\|12\|0 |
| O70.0: First degree perineal laceration during delivery | 1.3E-02 | 6.3 (1.5, 27) | 1.9E-04 | 2360\|3\|0 | 209402\|77\|0 |
| First weekday of wear - any visit - Tuesday AnyInst | 1.3E-02 | 5.8 (1.4, 24) | 1.9E-04 | 1605\|3\|0 | 90054\|31\|0 |

| Phenotype | P value | Odds ratio (CI) | AAF | Cases RR\|RA\|AA | Controls RR\|RA\|AA |
|---|---|---|---|---|---|
| Ever smoked - any visit - No AnyInst | 1.3E-02 | 1.5 (1.1, 2) | 2.0E-04 | 170817\|84\|0 | 257988\|87\|0 |
| Rubella german measles, Self-reported - any visit | 1.3E-02 | 4.3 (1.4, 14) | 2.1E-04 | 2243\|4\|0 | 329123\|134\|0 |
| Pizza intake - any visit - 1 AnyInst | 1.3E-02 | 8.6 (1.6, 48) | 1.9E-04 | 3292\|3\|0 | 9842\|2\|0 |
| OPCS4 S15.1: Biopsy of lesion of skin of head or neck NEC | 1.4E-02 | 8.3 (1.5, 44) | 2.0E-04 | 804\|2\|0 | 350861\|142\|0 |
| FI7 synonym - any visit - Rest AnyInst | 1.4E-02 | 5.7 (1.4, 23) | 2.1E-04 | 1458\|3\|0 | 125363\|51\|0 |
| Which eyes affected astigmatism - any visit - R eye AnyInst | 1.4E-02 | 12 (1.6, 82) | 1.4E-04 | 2587\|3\|0 | 15036\|2\|0 |
| hydrocortisone - any visit | 1.4E-02 | 8.1 (1.5, 43) | 2.0E-04 | 839\|2\|0 | 319547\|129\|0 |
| OPCS4 S36.1: Full thickness autograft of skin to head or neck | 1.4E-02 | 5.4 (1.4, 21) | 2.0E-04 | 1673\|3\|0 | 349992\|141\|0 |
| M47.9: Spondylosis, unspecified | 1.4E-02 | 8 (1.5, 43) | 2.0E-04 | 891\|2\|0 | 386594\|153\|0 |
| I67.9: Cerebrovascular disease, unspecified | 1.5E-02 | 5.4 (1.4, 21) | 2.0E-04 | 1466\|3\|0 | 386017\|152\|0 |
| Willing to attempt cognitive tests - any visit - I am unable to try this AnyInst | 1.5E-02 | 2.9 (1.2, 6.9) | 2.0E-04 | 6204\|7\|0 | 424036\|165\|0 |
| L73.9: Follicular disorder, unspecified | 1.5E-02 | 7.9 (1.5, 42) | 2.0E-04 | 678\|2\|0 | 384535\|151\|0 |
| Intake of sugar added to coffee - any visit - 1 AnyInst | 1.5E-02 | 3.7 (1.3, 10) | 2.7E-04 | 16909\|14\|0 | 14640\|3\|0 |
| Z86.7: Personal history of diseases of the circulatory system | 1.5E-02 | 2 (1.1, 3.6) | 2.0E-04 | 22857\|16\|0 | 360357\|136\|0 |
| I12.0: Hypertensive chronic kidney disease with stage 5 chronic kidney disease or end stage renal disease | 1.5E-02 | 5.3 (1.4, 21) | 2.0E-04 | 1500\|3\|0 | 385985\|152\|0 |
| Dried fruit intake - any visit - 2 AnyInst | 1.5E-02 | 5 (1.4, 18) | 2.5E-04 | 2265\|4\|0 | 29646\|12\|0 |
| 3mm asymmetry index irregular astigmatism level R - any visit - normal AnyInst | 1.6E-02 | 0.18 (0.047, 0.73) | 2.1E-04 | 99437\|39\|0 | 2013\|3\|0 |
| L92: Granulomatous disorders of skin and subcutaneous tissue | 1.6E-02 | 8 (1.5, 43) | 2.0E-04 | 823\|2\|0 | 385945\|153\|0 |
| M80.9: Unspecified osteoporosis with pathological fracture | 1.6E-02 | 7.9 (1.5, 43) | 2.0E-04 | 813\|2\|0 | 386672\|153\|0 |
| Number of days out of 10 with abdominal pain - any visit - 9 days with pain AnyInst | 1.6E-02 | 8.8 (1.5, 51) | 1.9E-04 | 927\|2\|0 | 43716\|15\|0 |
| OPCS4 E85.1: Invasive ventilation | 1.6E-02 | 5.2 (1.4, 20) | 2.0E-04 | 1707\|3\|0 | 349958\|141\|0 |
| Size of white wine glass drunk - any visit - small 125ml AnyInst | 1.6E-02 | 0.24 (0.075, 0.77) | 1.9E-04 | 18239\|2\|0 | 24170\|14\|0 |
| R54: Age-related physical debility | 1.6E-02 | 7.7 (1.5, 41) | 2.0E-04 | 753\|2\|0 | 385988\|153\|0 |
| Index for card A in round - any visit - Fir tree 3 overlapping triangles AnyInst | 1.6E-02 | 0.68 (0.5, 0.93) | 2.0E-04 | 187988\|60\|0 | 241053\|111\|0 |
| F52: Sexual dysfunction not due to a substance or known physiological condition | 1.7E-02 | 7.7 (1.4, 41) | 2.0E-04 | 680\|2\|0 | 385485\|152\|0 |
| I69: Sequelae of cerebrovascular disease | 1.7E-02 | 5.2 (1.3, 20) | 2.0E-04 | 1710\|3\|0 | 385769\|152\|0 |
| OPCS4 W37.1: Primary total prosthetic replacement of hip joint using cement | 1.7E-02 | 3.1 (1.2, 8) | 2.0E-04 | 6319\|6\|0 | 345346\|138\|0 |

| Phenotype | P value | Odds ratio (CI) | AAF | Cases RR\|RA\|AA | Controls RR\|RA\|AA |
|---|---|---|---|---|---|
| R60.9: Edema, unspecified | 1.7E-02 | 4.1 (1.3, 13) | 2.0E-04 | 2766\|4\|0 | 378557\|148\|0 |
| Device used to enter occupational data - any visit - Deskop computer AnyInst | 1.7E-02 | 0.43 (0.22, 0.86) | 2.1E-04 | 40418\|9\|0 | 64408\|34\|0 |
| Liver intake - any visit - 1 AnyInst | 1.7E-02 | 0.13 (0.023, 0.69) | 3.0E-04 | 5113\|1\|0 | 3108\|4\|0 |
| C77.9: Secondary and unspecified malignant neoplasm of lymph node, unspecified | 1.7E-02 | 7.6 (1.4, 41) | 2.0E-04 | 648\|2\|0 | 386836\|153\|0 |
| Day of week questionnaire completed - any visit - Friday AnyInst | 1.8E-02 | 1.8 (1.1, 3) | 1.9E-04 | 51044\|28\|0 | 134385\|42\|0 |
| J18.1: Lobar pneumonia, unspecified organism | 1.8E-02 | 2.6 (1.2, 5.8) | 2.0E-04 | 8248\|8\|0 | 379022\|147\|0 |
| Sex of baby - any visit - Male AnyInst | 1.8E-02 | 0.063 (0.0064, 0.62) | 2.0E-04 | 5216\|0\|0 | 7428\|5\|0 |
| I12: Hypertensive chronic kidney disease | 1.8E-02 | 5.1 (1.3, 19) | 2.0E-04 | 1656\|3\|0 | 385811\|152\|0 |
| Types of spread used on bread crackers - any visit - Polyunsaturated margarine on bread crackers AnyInst | 1.8E-02 | 1.9 (1.1, 3.4) | 1.8E-04 | 50495\|26\|0 | 92518\|25\|0 |
| Feelings of worthlessness during worst period of depression - any visit - Yes AnyInst | 1.8E-02 | 2.6 (1.2, 5.6) | 2.0E-04 | 37577\|21\|0 | 36139\|8\|0 |
| Feelings of worthlessness during worst period of depression - any visit - No AnyInst | 1.8E-02 | 0.39 (0.18, 0.85) | 2.0E-04 | 36139\|8\|0 | 37577\|21\|0 |
| Recent trouble relaxing - any visit - Nearly every day AnyInst | 1.8E-02 | 3.6 (1.2, 10) | 1.9E-04 | 4287\|5\|0 | 134836\|47\|0 |
| Intake of sugar added to cereal - any visit - half AnyInst | 1.8E-02 | 5 (1.3, 19) | 1.2E-04 | 14164\|7\|0 | 22674\|2\|0 |
| OPCS4 A55.9: Unspecified diagnostic spinal puncture | 1.8E-02 | 3.1 (1.2, 7.7) | 2.0E-04 | 5412\|6\|0 | 346253\|138\|0 |
| OPCS4 L88.1: Percutaneous transluminal laser ablation of long saphenous vein | 1.8E-02 | 7.5 (1.4, 40) | 2.0E-04 | 650\|2\|0 | 351015\|142\|0 |
| Z71.2: Person consulting for explanation of examination or test findings | 1.8E-02 | 0.11 (0.019, 0.69) | 2.0E-04 | 11107\|0\|0 | 364211\|150\|0 |
| Ever smoked - any visit - Yes AnyInst | 1.8E-02 | 0.69 (0.51, 0.94) | 2.0E-04 | 262929\|90\|0 | 165876\|81\|0 |
| Liver biliary pancreas problem, ICD10: K87 or Self-reported - any visit | 1.8E-02 | 7.3 (1.4, 38) | 2.1E-04 | 751\|2\|0 | 327915\|135\|0 |
| Handedness chirality laterality - any visit - Left handed AnyInst | 1.8E-02 | 1.7 (1.1, 2.6) | 2.0E-04 | 41001\|26\|0 | 389112\|146\|0 |
| Liver biliary pancreas problem, Self-reported - any visit | 1.8E-02 | 7.3 (1.4, 38) | 2.1E-04 | 794\|2\|0 | 330572\|136\|0 |
| Hypertension, ICD10: I10 or Self-reported - any visit[2] | 1.9E-02 | 1.5 (1.1, 2.2) | 2.1E-04 | 100060\|56\|0 | 228604\|81\|0 |
| Other sweets intake - any visit - 1 AnyInst | 1.9E-02 | 14 (1.5, 130) | 2.8E-04 | 4660\|5\|0 | 4302\|0\|0 |
| L03.0: Cellulitis and acute lymphangitis of finger and toe | 1.9E-02 | 7.3 (1.4, 38) | 2.0E-04 | 690\|2\|0 | 386794\|153\|0 |
| OPCS4 F10.9: Unspecified simple extraction of tooth | 1.9E-02 | 4.9 (1.3, 19) | 2.0E-04 | 1641\|3\|0 | 350024\|141\|0 |
| Frequency of consuming six or more units of alcohol - any visit - Monthly AnyInst | 1.9E-02 | 0.11 (0.018, 0.7) | 1.7E-04 | 11321\|0\|0 | 116681\|44\|0 |
| Fizzy drink intake - any visit - 3 AnyInst | 2.0E-02 | 8.9 (1.4, 56) | 1.2E-04 | 1183\|2\|0 | 28369\|5\|0 |

| Phenotype | P value | Odds ratio (CI) | AAF | Cases RR\|RA\|AA | Controls RR\|RA\|AA |
|---|---|---|---|---|---|
| Illnesses of siblings - any visit - High blood pressure AnyInst | 2.0E-02 | 1.6 (1.1, 2.2) | 2.0E-04 | 75887\|43\|0 | 295405\|109\|0 |
| Berry intake - any visit - half AnyInst | 2.0E-02 | 2.9 (1.2, 7.3) | 1.8E-04 | 16019\|11\|0 | 36496\|8\|0 |
| Hands free device speakerphone use with mobile phone in last 3 month - any visit - More than half the time AnyInst | 2.0E-02 | 0.12 (0.019, 0.71) | 2.1E-04 | 8569\|0\|0 | 356995\|153\|0 |
| K61: Abscess of anal and rectal regions | 2.0E-02 | 4.8 (1.3, 18) | 2.0E-04 | 1730\|3\|0 | 384971\|152\|0 |
| I63: Cerebral infarction | 2.1E-02 | 3.3 (1.2, 9.3) | 2.0E-04 | 4511\|5\|0 | 382181\|149\|0 |
| OPCS4 U19.1: Implantation of electrocardiography loop recorder | 2.1E-02 | 6.9 (1.3, 36) | 2.0E-04 | 936\|2\|0 | 350729\|142\|0 |
| R35: Polyuria | 2.1E-02 | 2.7 (1.2, 6.4) | 1.9E-04 | 7533\|7\|0 | 369584\|138\|0 |
| Number of bread slices with butter margarine - any visit - 6 AnyInst | 2.2E-02 | 4.2 (1.2, 14) | 1.7E-04 | 3328\|4\|0 | 118374\|37\|0 |
| Type milk consumed - any visit - did not have milk AnyInst | 2.2E-02 | 2.2 (1.1, 4.2) | 1.9E-04 | 15496\|13\|0 | 168978\|56\|0 |
| Sliced bread intake - any visit - 6 AnyInst | 2.2E-02 | 3.5 (1.2, 10) | 1.7E-04 | 4694\|5\|0 | 137733\|43\|0 |
| Skin colour - any visit - Fair AnyInst | 2.2E-02 | 1.6 (1.1, 2.3) | 2.0E-04 | 303672\|128\|0 | 121199\|39\|0 |
| OPCS4 S08.2: Curettage and cauterisation of lesion of skin NEC | 2.2E-02 | 6.7 (1.3, 35) | 2.0E-04 | 929\|2\|0 | 350736\|142\|0 |
| R29.8: Other symptoms and signs involving the nervous and musculoskeletal systems | 2.3E-02 | 3.7 (1.2, 12) | 2.0E-04 | 2851\|4\|0 | 380356\|151\|0 |
| R29: Other symptoms and signs involving the nervous and musculoskeletal systems | 2.3E-02 | 2.9 (1.2, 7.3) | 2.0E-04 | 6027\|6\|0 | 375014\|147\|0 |
| OPCS4 M47.3: Removal of urethral catheter from bladder | 2.3E-02 | 2.7 (1.1, 6.5) | 2.0E-04 | 6580\|7\|0 | 345085\|137\|0 |
| J00: Acute nasopharyngitis [common cold] | 2.4E-02 | 3.7 (1.2, 12) | 2.0E-04 | 2554\|4\|0 | 376647\|148\|0 |
| Device used to enter occupational data - any visit - Laptop computer AnyInst | 2.4E-02 | 2 (1.1, 3.6) | 2.1E-04 | 39025\|23\|0 | 65801\|20\|0 |
| Time spent doing vigorous physical activity - any visit - None AnyInst | 2.4E-02 | 1.9 (1.1, 3.4) | 1.9E-04 | 131929\|58\|0 | 53500\|12\|0 |
| N86: Erosion and ectropion of cervix uteri | 2.4E-02 | 6.7 (1.3, 35) | 1.9E-04 | 1105\|2\|0 | 209055\|78\|0 |
| Past tobacco smoking - any visit - I have never smoked AnyInst | 2.4E-02 | 1.4 (1, 2) | 2.0E-04 | 170865\|84\|0 | 225768\|76\|0 |
| Rheumatoid factor reportability - any visit - Not reportable at assay too low AnyInst | 2.4E-02 | 0.59 (0.37, 0.93) | 2.0E-04 | 373485\|138\|0 | 38056\|24\|0 |
| OPCS4 P13.2: Female perineorrhaphy | 2.4E-02 | 6.7 (1.3, 35) | 2.0E-04 | 823\|2\|0 | 350842\|142\|0 |
| Maximum frequency of taking cannabis - any visit - Once a month or more but not every week AnyInst | 2.4E-02 | 5.4 (1.2, 24) | 1.7E-04 | 3575\|3\|0 | 26370\|7\|0 |
| Been in a confiding relationship as an adult - any visit - Rarely true AnyInst | 2.4E-02 | 3 (1.2, 7.7) | 1.8E-04 | 6416\|6\|0 | 129546\|44\|0 |
| UK Biobank assessment centre - any visit - Newcastle imaging AnyInst | 2.5E-02 | 5.2 (1.2, 22) | 2.7E-04 | 9735\|9\|0 | 29018\|12\|0 |
| J69: Pneumonitis due to solids and liquids | 2.5E-02 | 6.5 (1.3, 34) | 2.0E-04 | 1137\|2\|0 | 386307\|153\|0 |
| Detention categories - any visit - Formally detained under Part II Mental Health Act 1983 AnyInst | 2.5E-02 | 5.3 (1.2, 23) | 1.9E-04 | 2669\|3\|0 | 33692\|11\|0 |

| Phenotype | P value | Odds ratio (CI) | AAF | Cases RR\|RA\|AA | Controls RR\|RA\|AA |
|---|---|---|---|---|---|
| Bring up phlegm sputum mucus on most days - any visit - Yes AnyInst | 2.5E-02 | 2.6 (1.1, 6.2) | 2.1E-04 | 9274\|8\|0 | 98828\|37\|0 |
| Bring up phlegm sputum mucus on most days - any visit - No AnyInst | 2.5E-02 | 0.38 (0.16, 0.88) | 2.1E-04 | 98828\|37\|0 | 9274\|8\|0 |
| Z09: Encounter for follow-up examination after completed treatment for conditions other than malignant neoplasm | 2.5E-02 | 0.31 (0.11, 0.86) | 2.0E-04 | 18207\|2\|0 | 364521\|151\|0 |
| IPAQ activity group - any visit - moderate AnyInst | 2.5E-02 | 1.5 (1, 2.1) | 2.0E-04 | 142467\|69\|0 | 206512\|68\|0 |
| Ordering of blows - any visit - third earliest i e latest AnyInst | 2.5E-02 | 1.6 (1.1, 2.3) | 2.0E-04 | 298271\|135\|0 | 96713\|27\|0 |
| Result ranking - any visit - third best i e worst AnyInst | 2.5E-02 | 1.6 (1.1, 2.3) | 2.0E-04 | 298271\|135\|0 | 96713\|27\|0 |
| 3mm regularity index irregular astigmatism level L - any visit - high possibility abnormality AnyInst | 2.6E-02 | 3.4 (1.2, 9.8) | 2.0E-04 | 4385\|5\|0 | 96801\|36\|0 |
| J69.0: Pneumonitis due to inhalation of food and vomit | 2.6E-02 | 6.5 (1.3, 33) | 2.0E-04 | 1098\|2\|0 | 386387\|153\|0 |
| M24: Other specific joint derangements | 2.6E-02 | 3.6 (1.2, 11) | 2.0E-04 | 3196\|4\|0 | 383526\|151\|0 |
| Melon intake - any visit - 2 AnyInst | 2.6E-02 | 7.9 (1.3, 48) | 2.3E-04 | 1428\|2\|0 | 13584\|5\|0 |
| OPCS4 Z94.3: Left sided operation | 2.6E-02 | 1.5 (1, 2) | 2.0E-04 | 132881\|67\|0 | 218784\|77\|0 |
| Ever had cervical smear test - any visit - No AnyInst | 2.6E-02 | 3.2 (1.1, 8.9) | 1.9E-04 | 5066\|5\|0 | 227806\|82\|0 |
| Chickenpox, ICD10: B01 or Self-reported - any visit | 2.6E-02 | 2.6 (1.1, 6.1) | 2.1E-04 | 6567\|7\|0 | 322099\|130\|0 |
| Destinations on discharge from hospital recoded - any visit - Transfer within NHS provider Medical specialty AnyInst | 2.7E-02 | 6.6 (1.2, 35) | 2.0E-04 | 1962\|2\|0 | 341122\|136\|0 |
| Difference in mobile phone use compared to two years previously - any visit - No AnyInst | 2.7E-02 | 0.7 (0.51, 0.96) | 2.1E-04 | 201566\|71\|0 | 164163\|81\|0 |
| N18: Chronic kidney disease | 2.7E-02 | 2.3 (1.1, 5) | 2.0E-04 | 10130\|9\|0 | 372275\|145\|0 |
| R14.0: Abdominal distension | 2.7E-02 | 6.3 (1.2, 32) | 2.0E-04 | 780\|2\|0 | 383609\|153\|0 |
| RGC Any Ischemic Stroke | 2.7E-02 | 2.8 (1.1, 7.2) | 2.0E-04 | 6249\|6\|0 | 270105\|106\|0 |
| Loneliness isolation - any visit - Yes AnyInst | 2.7E-02 | 1.5 (1, 2.1) | 2.0E-04 | 79807\|44\|0 | 344106\|126\|0 |
| R25.1: Tremor, unspecified | 2.7E-02 | 6.2 (1.2, 32) | 2.0E-04 | 1051\|2\|0 | 384860\|153\|0 |
| OPCS4 H33.3: Anterior resection of rectum and anastomosis of colon to rectum using staples | 2.7E-02 | 6.3 (1.2, 32) | 2.0E-04 | 833\|2\|0 | 350832\|142\|0 |
| H53.0: Amblyopia ex anopsia | 2.7E-02 | 6.3 (1.2, 32) | 2.0E-04 | 792\|2\|0 | 386503\|153\|0 |
| RGC Ankylosing Spondylitis, ICD10 M45 only, HLA-B27 positive background | 2.7E-02 | 14 (1.4, 150) | 1.6E-04 | 525\|1\|0 | 30618\|9\|0 |
| N63: Unspecified lump in breast | 2.7E-02 | 2.8 (1.1, 7.2) | 1.9E-04 | 6304\|6\|0 | 195482\|70\|0 |
| Type of special diet followed - any visit - Low calorie AnyInst | 2.7E-02 | 2.7 (1.1, 6.7) | 2.4E-04 | 30105\|19\|0 | 20527\|5\|0 |
| A49: Bacterial infection of unspecified site | 2.7E-02 | 6.2 (1.2, 31) | 2.0E-04 | 1114\|2\|0 | 385399\|153\|0 |
| serevent 25mcg inhaler - any visit | 2.8E-02 | 6.2 (1.2, 31) | 2.0E-04 | 1168\|2\|0 | 319218\|129\|0 |

| Phenotype | P value | Odds ratio (CI) | AAF | Cases RR\|RA\|AA | Controls RR\|RA\|AA |
|---|---|---|---|---|---|
| Hypertension, Self-reported - any visit | 2.8E-02 | 1.5 (1, 2.1) | 2.1E-04 | 115292\|62\|0 | 216074\|76\|0 |
| Ever had bowel cancer screening - any visit - No AnyInst | 2.8E-02 | 1.5 (1, 2.2) | 2.0E-04 | 289067\|126\|0 | 134851\|41\|0 |
| Porridge intake - any visit - 2 AnyInst | 2.8E-02 | 7 (1.2, 39) | 2.1E-04 | 800\|2\|0 | 43850\|17\|0 |
| Carer support indicators - any visit - Yes AnyInst | 2.8E-02 | 3.4 (1.1, 10) | 2.9E-04 | 15143\|15\|0 | 16017\|3\|0 |
| Time spent doing vigorous physical activity - any visit - 10 30 minutes AnyInst | 2.8E-02 | 1.8 (1.1, 3.1) | 1.9E-04 | 36805\|21\|0 | 148624\|49\|0 |
| H01.0: Blepharitis | 2.8E-02 | 4.3 (1.2, 16) | 2.0E-04 | 1876\|3\|0 | 381054\|152\|0 |
| Workplace very noisy - any visit - 141 Often AnyInst | 2.9E-02 | 2.1 (1.1, 4) | 2.1E-04 | 20894\|15\|0 | 85854\|29\|0 |
| oxybutynin - any visit | 2.9E-02 | 6.1 (1.2, 31) | 2.0E-04 | 1069\|2\|0 | 319317\|129\|0 |
| J90: Pleural effusion, not elsewhere classified | 2.9E-02 | 2.6 (1.1, 6) | 2.0E-04 | 7216\|7\|0 | 379974\|148\|0 |
| Type of baguette eaten - any visit - wholemeal AnyInst | 2.9E-02 | 3.2 (1.1, 9.3) | 2.5E-04 | 5285\|6\|0 | 26844\|10\|0 |
| OPCS4 E85.2: Non-invasive ventilation NEC | 2.9E-02 | 3.5 (1.1, 11) | 2.0E-04 | 3020\|4\|0 | 348645\|140\|0 |
| H61: Other disorders of external ear | 2.9E-02 | 2.1 (1.1, 4.1) | 1.9E-04 | 14531\|11\|0 | 355432\|133\|0 |
| OPCS4 A52.2: Therapeutic sacral epidural injection | 2.9E-02 | 3.5 (1.1, 11) | 2.0E-04 | 3126\|4\|0 | 348539\|140\|0 |
| G81.9: Hemiplegia, unspecified | 3.0E-02 | 4.3 (1.2, 16) | 2.0E-04 | 2020\|3\|0 | 385316\|152\|0 |
| Size of white wine glass drunk - any visit - medium 175ml AnyInst | 3.0E-02 | 3.7 (1.1, 12) | 1.9E-04 | 25985\|14\|0 | 16424\|2\|0 |
| N90: Other noninflammatory disorders of vulva and perineum | 3.0E-02 | 3.5 (1.1, 11) | 1.9E-04 | 3230\|4\|0 | 205355\|75\|0 |
| Types of spread used on bread crackers - any visit - Low fat olive spread on bread crackers AnyInst | 3.0E-02 | 2.3 (1.1, 4.8) | 1.8E-04 | 15459\|10\|0 | 127554\|41\|0 |
| FI6 conditional arithmetic - any visit - 72 AnyInst | 3.0E-02 | 6.2 (1.2, 32) | 1.8E-04 | 1057\|2\|0 | 95022\|33\|0 |
| J02: Acute pharyngitis | 3.1E-02 | 2.3 (1.1, 4.7) | 2.0E-04 | 10871\|9\|0 | 350667\|136\|0 |
| RGC Any Stroke[2] | 3.1E-02 | 1.9 (1.1, 3.5) | 2.1E-04 | 18046\|14\|0 | 271922\|108\|0 |
| Illnesses of mother - any visit - Bowel cancer AnyInst | 3.1E-02 | 1.9 (1.1, 3.3) | 2.0E-04 | 20755\|15\|0 | 402437\|155\|0 |
| R26: Abnormalities of gait and mobility | 3.2E-02 | 3.4 (1.1, 11) | 2.0E-04 | 3388\|4\|0 | 382549\|150\|0 |
| C79.3: Secondary malignant neoplasm of brain and cerebral meninges | 3.2E-02 | 5.8 (1.2, 29) | 2.0E-04 | 1143\|2\|0 | 386342\|153\|0 |
| K92.2: Gastrointestinal hemorrhage, unspecified | 3.2E-02 | 2.5 (1.1, 5.8) | 2.0E-04 | 7785\|7\|0 | 379636\|148\|0 |
| Mineral and other dietary supplements - any visit - Fish oil including cod liver oil AnyInst | 3.2E-02 | 1.9 (1.1, 3.5) | 1.7E-04 | 139718\|55\|0 | 50400\|11\|0 |
| W19: Unspecified fall | 3.2E-02 | 2.5 (1.1, 5.8) | 2.0E-04 | 8499\|7\|0 | 372198\|147\|0 |
| G81: Hemiplegia and hemiparesis | 3.2E-02 | 4.2 (1.1, 16) | 2.0E-04 | 2066\|3\|0 | 385110\|152\|0 |

| Phenotype | P value | Odds ratio (CI) | AAF | Cases RR\|RA\|AA | Controls RR\|RA\|AA |
|---|---|---|---|---|---|
| Pure fruit vegetable juice intake - any visit - half AnyInst | 3.2E-02 | 0.22 (0.056, 0.88) | 2.2E-04 | 12960\|1\|0 | 26081\|16\|0 |
| Never eat eggs dairy wheat sugar - any visit - I eat all of the above AnyInst | 3.2E-02 | 1.5 (1, 2.3) | 2.0E-04 | 335176\|145\|0 | 93990\|26\|0 |
| K65: Peritonitis | 3.2E-02 | 5.8 (1.2, 29) | 2.0E-04 | 1098\|2\|0 | 386185\|153\|0 |
| Belittlement by partner or ex partner as an adult - any visit - Very often true AnyInst | 3.3E-02 | 4.3 (1.1, 17) | 1.8E-04 | 2890\|3\|0 | 136263\|47\|0 |
| Pure fruit vegetable juice intake - any visit - 3 AnyInst | 3.3E-02 | 6.3 (1.2, 34) | 2.2E-04 | 1007\|2\|0 | 38034\|15\|0 |
| OPCS O302 | 3.3E-02 | 2.7 (1.1, 6.7) | 2.0E-04 | 5346\|6\|0 | 346319\|138\|0 |
| Low calorie drink intake - any visit - 3 AnyInst | 3.4E-02 | 3.3 (1.1, 9.8) | 2.3E-04 | 3872\|5\|0 | 34423\|13\|0 |
| OPCS4 M65.3: Endoscopic resection of prostate NEC | 3.4E-02 | 3.5 (1.1, 11) | 2.0E-04 | 3114\|4\|0 | 348551\|140\|0 |
| Recent poor appetite or overeating - any visit - Nearly every day AnyInst | 3.4E-02 | 3.5 (1.1, 11) | 1.9E-04 | 3488\|4\|0 | 135873\|48\|0 |
| Month of attending assessment centre - any visit - October AnyInst | 3.5E-02 | 1.6 (1, 2.5) | 2.0E-04 | 41105\|25\|0 | 389394\|147\|0 |
| Felt distant from other people in past month - any visit - Extremely AnyInst | 3.5E-02 | 6.4 (1.1, 36) | 2.1E-04 | 1006\|2\|0 | 61744\|24\|0 |
| OPCS4 S13.1: Punch biopsy of lesion of skin of head or neck | 3.5E-02 | 4 (1.1, 15) | 2.0E-04 | 2598\|3\|0 | 349067\|141\|0 |
| Physically abused by family as a child - any visit - Often AnyInst | 3.5E-02 | 5.7 (1.1, 29) | 1.9E-04 | 1185\|2\|0 | 138071\|50\|0 |
| OPCS4 Y98.3: Radiology of three body areas (or 20-40 minutes) | 3.6E-02 | 2.7 (1.1, 6.6) | 2.0E-04 | 6000\|6\|0 | 345665\|138\|0 |
| RGC_CKD_CC | 3.6E-02 | 2.5 (1.1, 5.8) | 2.0E-04 | 7601\|7\|0 | 353721\|139\|0 |
| R56.8: Other and unspecified convulsions | 3.7E-02 | 4 (1.1, 15) | 2.0E-04 | 2533\|3\|0 | 384952\|152\|0 |
| Degree bothered by feeling heart pound race in the last 3 months - any visit - 602 Bothered a lot AnyInst | 3.7E-02 | 3 (1.1, 8.3) | 1.9E-04 | 4936\|5\|0 | 150007\|53\|0 |
| N99.3: Prolapse of vaginal vault after hysterectomy | 3.7E-02 | 11 (1.2, 100) | 1.9E-04 | 564\|1\|0 | 228322\|85\|0 |
| K92: Other diseases of digestive system | 3.7E-02 | 2.1 (1, 4.2) | 2.0E-04 | 12713\|10\|0 | 372417\|144\|0 |
| Frequency of consuming six or more units of alcohol - any visit - Daily or almost daily AnyInst | 3.7E-02 | 3.5 (1.1, 11) | 1.7E-04 | 4451\|4\|0 | 123551\|40\|0 |
| Mixed vegetable intake - any visit - 1 AnyInst | 3.7E-02 | 0.33 (0.12, 0.94) | 2.4E-04 | 21014\|6\|0 | 8490\|8\|0 |
| Wants to stop smoking - any visit - No probably not AnyInst | 3.7E-02 | 3.6 (1.1, 12) | 1.7E-04 | 8240\|6\|0 | 24733\|5\|0 |
| OPCS4 Q17.8: Other specified therapeutic endoscopic operations on uterus | 3.7E-02 | 5.6 (1.1, 29) | 2.0E-04 | 643\|2\|0 | 351022\|142\|0 |
| Satisfaction with bowel habits - any visit - 2 AnyInst | 3.7E-02 | 0.37 (0.15, 0.94) | 1.8E-04 | 21578\|3\|0 | 132653\|54\|0 |
| Epilepsy, Self-reported - any visit | 3.8E-02 | 3.3 (1.1, 10) | 2.1E-04 | 3587\|4\|0 | 327779\|134\|0 |
| FI5 family relationship calculation - any visit - cousin AnyInst | 3.8E-02 | 0.51 (0.27, 0.96) | 1.8E-04 | 59412\|15\|0 | 47294\|24\|0 |
| O68.0: Labour and delivery complicated by fetal heart rate anomaly | 3.8E-02 | 6.1 (1.1, 34) | 1.9E-04 | 1404\|2\|0 | 227482\|84\|0 |

| Phenotype | P value | Odds ratio (CI) | AAF | Cases RR\|RA\|AA | Controls RR\|RA\|AA |
|---|---|---|---|---|---|
| Chocolate bar intake - any visit - quarter AnyInst | 3.8E-02 | 7.4 (1.1, 49) | 9.9E-05 | 2276\|2\|0 | 22852\|3\|0 |
| C56: Malignant neoplasm of ovary | 3.8E-02 | 5.5 (1.1, 27) | 1.9E-04 | 1184\|2\|0 | 210237\|78\|0 |
| K92.0: Hematemesis | 3.8E-02 | 3.9 (1.1, 14) | 2.0E-04 | 2298\|3\|0 | 385149\|152\|0 |
| Instant coffee intake - any visit - 4 AnyInst | 3.8E-02 | 2.1 (1, 4.2) | 2.2E-04 | 15574\|12\|0 | 88533\|34\|0 |
| R59: Enlarged lymph nodes | 3.8E-02 | 3.2 (1.1, 9.9) | 2.0E-04 | 3661\|4\|0 | 380186\|150\|0 |
| UK Biobank assessment centre - any visit - Middlesborough AnyInst | 3.9E-02 | 0.39 (0.16, 0.95) | 2.0E-04 | 18960\|4\|0 | 411539\|168\|0 |
| M24.1: Other articular cartilage disorders | 3.9E-02 | 5.4 (1.1, 26) | 2.0E-04 | 1155\|2\|0 | 386313\|153\|0 |
| How are people in household related to participant - any visit - Husband wife or partner AnyInst | 3.9E-02 | 0.58 (0.35, 0.97) | 1.9E-04 | 316924\|114\|0 | 32740\|20\|0 |
| H93.29: Other abnormal auditory perceptions | 3.9E-02 | 5.3 (1.1, 26) | 2.0E-04 | 1158\|2\|0 | 382023\|152\|0 |
| H93.299: Other abnormal auditory perceptions, unspecified ear | 3.9E-02 | 5.3 (1.1, 26) | 2.0E-04 | 1158\|2\|0 | 382023\|152\|0 |
| Length of longest manic irritable episode - any visit - Less than a week AnyInst | 3.9E-02 | 2.9 (1.1, 8) | 2.9E-04 | 5602\|7\|0 | 23198\|10\|0 |
| N99: Intraoperative and postprocedural complications and disorders of genitourinary system, not elsewhere classified | 4.0E-02 | 5.3 (1.1, 26) | 2.0E-04 | 1363\|2\|0 | 386023\|153\|0 |
| OPCS4 P23.1: Anterior and posterior colporrhaphy NEC | 4.0E-02 | 4 (1.1, 15) | 2.0E-04 | 2349\|3\|0 | 349316\|141\|0 |
| Vitamin and or mineral supplement use - any visit - Vitamin E AnyInst | 4.0E-02 | 3 (1, 8.5) | 2.1E-04 | 4698\|5\|0 | 86688\|33\|0 |
| H93.2: Other abnormal auditory perceptions | 4.0E-02 | 5.3 (1.1, 26) | 2.0E-04 | 1174\|2\|0 | 382006\|152\|0 |
| I10: Essential (primary) hypertension | 4.1E-02 | 1.5 (1, 2.1) | 2.0E-04 | 104984\|54\|0 | 268794\|94\|0 |
| Friendships satisfaction - any visit - Very unhappy AnyInst | 4.1E-02 | 5.7 (1.1, 30) | 2.1E-04 | 790\|2\|0 | 174766\|73\|0 |
| M13.9: Arthritis, unspecified | 4.1E-02 | 5.2 (1.1, 26) | 2.0E-04 | 1280\|2\|0 | 386205\|153\|0 |
| Workplace full of chemical or other fumes - any visit - 131 Sometimes AnyInst | 4.1E-02 | 1.9 (1, 3.7) | 2.1E-04 | 25527\|17\|0 | 79280\|28\|0 |
| Someone to take to doctor when needed as a child - any visit - Never true AnyInst | 4.1E-02 | 3.9 (1.1, 15) | 1.9E-04 | 2772\|3\|0 | 135856\|49\|0 |
| Grape intake - any visit - 3 AnyInst | 4.1E-02 | 5.6 (1.1, 29) | 2.0E-04 | 1143\|2\|0 | 49439\|18\|0 |
| I69.4: Sequelae of stroke, not specified as haemorrhage or infarction | 4.1E-02 | 5.2 (1.1, 25) | 2.0E-04 | 1219\|2\|0 | 386266\|153\|0 |
| Size of red wine glass drunk - any visit - small 125ml AnyInst | 4.2E-02 | 2.5 (1, 5.8) | 2.0E-04 | 21334\|13\|0 | 32152\|8\|0 |
| H04.1: Other disorders of lacrimal gland | 4.2E-02 | 9.9 (1.1, 90) | 2.0E-04 | 519\|1\|0 | 386960\|154\|0 |
| G45.9: Transient cerebral ischemic attack, unspecified | 4.2E-02 | 3.8 (1, 14) | 2.0E-04 | 2490\|3\|0 | 384983\|152\|0 |
| Length of mobile phone use - any visit - Two to four years AnyInst | 4.2E-02 | 1.5 (1, 2.1) | 2.0E-04 | 76128\|39\|0 | 349079\|130\|0 |
| OPCS4 P05.4: Excision of lesion of vulva NEC | 4.3E-02 | 5.2 (1.1, 26) | 2.0E-04 | 1149\|2\|0 | 350516\|142\|0 |

| Phenotype | P value | Odds ratio (CI) | AAF | Cases RR\|RA\|AA | Controls RR\|RA\|AA |
|---|---|---|---|---|---|
| Avocado intake - any visit - half AnyInst | 4.3E-02 | 0.17 (0.031, 0.94) | 3.0E-04 | 6074\|1\|0 | 5571\|6\|0 |
| Diagnosed with coeliac disease or gluten sensitivity - any visit - Yes AnyInst | 4.3E-02 | 3.9 (1, 15) | 1.9E-04 | 2908\|3\|0 | 150978\|54\|0 |
| Diagnosed with coeliac disease or gluten sensitivity - any visit - No AnyInst | 4.3E-02 | 0.25 (0.067, 0.96) | 1.9E-04 | 150978\|54\|0 | 2908\|3\|0 |
| M80: Osteoporosis with current pathological fracture | 4.3E-02 | 5.1 (1.1, 25) | 2.0E-04 | 1186\|2\|0 | 386109\|153\|0 |
| Recent changes in speed amount of moving or speaking - any visit - More than half the days AnyInst | 4.3E-02 | 5.2 (1.1, 26) | 1.9E-04 | 1122\|2\|0 | 138187\|50\|0 |
| Frequency of tenseness restlessness in last 2 weeks - any visit - Not at all AnyInst | 4.4E-02 | 1.5 (1, 2.1) | 2.0E-04 | 311374\|135\|0 | 103656\|32\|0 |
| OPCS4 H58.2: Drainage of perianal abscess | 4.4E-02 | 5.1 (1, 25) | 2.0E-04 | 1073\|2\|0 | 350592\|142\|0 |
| Leek intake - any visit - 1 AnyInst | 4.4E-02 | 0.21 (0.046, 0.96) | 2.3E-04 | 8364\|1\|0 | 8788\|7\|0 |
| Types of spread used on bread crackers - any visit - Normal fat polyunsaturated margarine on bread crackers AnyInst | 4.4E-02 | 2.3 (1, 5.3) | 1.8E-04 | 10353\|8\|0 | 132660\|43\|0 |
| Triplet correct right - any visit - yes AnyInst | 4.4E-02 | 0.19 (0.039, 0.96) | 2.1E-04 | 171418\|71\|0 | 1285\|2\|0 |
| Handedness chirality laterality - any visit - Right handed AnyInst | 4.5E-02 | 0.65 (0.42, 0.99) | 2.0E-04 | 382077\|144\|0 | 48036\|28\|0 |
| M25: Other joint disorder, not elsewhere classified | 4.5E-02 | 0.56 (0.32, 0.99) | 2.0E-04 | 46004\|11\|0 | 301761\|127\|0 |
| Thickness of butter margarine spread on sliced bread - any visit - medium AnyInst | 4.6E-02 | 0.49 (0.25, 0.99) | 1.6E-04 | 57938\|13\|0 | 45369\|20\|0 |
| White wine intake - any visit - 1 AnyInst | 4.6E-02 | 0.33 (0.11, 0.98) | 1.8E-04 | 20760\|3\|0 | 23321\|13\|0 |
| I67.8: Other specified cerebrovascular diseases | 4.6E-02 | 5 (1, 24) | 2.0E-04 | 1479\|2\|0 | 386006\|153\|0 |
| OPCS Z924 | 4.6E-02 | 2.5 (1, 6.2) | 2.0E-04 | 6888\|6\|0 | 344777\|138\|0 |
| RGC Ankylosing Spondylitis, ICD and self reported, HLA-B27 positive background | 4.7E-02 | 10 (1, 100) | 1.6E-04 | 714\|1\|0 | 30618\|9\|0 |
| N95.2: Postmenopausal atrophic vaginitis | 4.7E-02 | 5 (1, 25) | 1.9E-04 | 1315\|2\|0 | 227571\|84\|0 |
| Abdominal discomfort pain for 6 months or longer - any visit - Yes AnyInst | 4.7E-02 | 0.45 (0.21, 0.99) | 1.6E-04 | 43513\|9\|0 | 39667\|17\|0 |
| Abdominal discomfort pain for 6 months or longer - any visit - No AnyInst | 4.7E-02 | 2.2 (1, 4.8) | 1.6E-04 | 39667\|17\|0 | 43513\|9\|0 |
| OPCS4 H52.3: Injection of sclerosing substance into haemorrhoid | 4.7E-02 | 4.9 (1, 24) | 2.0E-04 | 1234\|2\|0 | 350431\|142\|0 |
| Type of fat oil used in cooking - any visit - Normal fat soft margarine AnyInst | 4.8E-02 | 5 (1, 24) | 1.9E-04 | 1333\|2\|0 | 166606\|61\|0 |
| OPCS Z865 | 4.8E-02 | 4.9 (1, 24) | 2.0E-04 | 1395\|2\|0 | 350270\|142\|0 |
| Milk intake - any visit - 1 AnyInst | 4.8E-02 | 8.8 (1, 76) | 2.7E-04 | 8350\|7\|0 | 4753\|0\|0 |
| Illnesses of siblings - any visit - 17 None of the above group 1 AnyInst | 4.8E-02 | 0.72 (0.51, 1) | 2.0E-04 | 231190\|84\|0 | 140102\|68\|0 |
| Country of birth UK elsewhere - any visit - Elsewhere AnyInst | 4.9E-02 | 2.1 (1, 4.2) | 2.0E-04 | 17974\|19\|0 | 412055\|153\|0 |

1. ExWAS summary statistics were available for 6,396 binary phenotypes out of 10,168 phenotypes tested for association in UKB 450K Europeans. Shown is the phenotype code, p value, odds ratio with 95% confidence interval, case RR|RA|AA counts, controls RR|RA|AA counts. Shown are phenotypes with p values < 0.05.
2. Highlight of phenotypes mentioned in the results section.

**Supplementary Table 20. Quantitative Phenotypes Associated with *NOTCH3* p.Arg1231Cys in UKB 450 K European Participants[1].**

| Phenotype | P value | Beta (95% CI) | AAF | Cases (RR\|RA\|AA) | Units |
|---|---|---|---|---|---|
| Mean MD in external capsule on FA skeleton right - imaging visit - value 0 | 5.40E-10 | 1.4 (0.96, 1.8) | 0.00028 | 33550\|19\|0 | AU |
| Mean MD in superior corona radiata on FA skeleton right - imaging visit - value 0 | 7.60E-10 | 1.4 (0.94, 1.8) | 0.00028 | 33550\|19\|0 | AU |
| Mean L3 in external capsule on FA skeleton right - imaging visit - value 0 | 2.30E-09 | 1.3 (0.9, 1.8) | 0.00028 | 33550\|19\|0 | AU |
| Mean MD in superior corona radiata on FA skeleton left - imaging visit - value 0 | 2.30E-09 | 1.3 (0.89, 1.8) | 0.00028 | 33550\|19\|0 | AU |
| Mean L2 in external capsule on FA skeleton right - imaging visit - value 0 | 3.60E-09 | 1.3 (0.89, 1.8) | 0.00028 | 33550\|19\|0 | AU |
| Mean MD in external capsule on FA skeleton left - imaging visit - value 0 | 4.20E-09 | 1.3 (0.88, 1.8) | 0.00028 | 33550\|19\|0 | AU |
| Mean L2 in external capsule on FA skeleton left - imaging visit - value 0 | 5.40E-09 | 1.3 (0.87, 1.8) | 0.00028 | 33550\|19\|0 | AU |
| Weighted mean MD in tract superior thalamic radiation left - imaging visit - value 0 | 7.20E-09 | 1.3 (0.86, 1.7) | 0.00028 | 33548\|19\|0 | AU |
| Weighted mean L3 in tract superior thalamic radiation left - imaging visit - value 0 | 7.70E-09 | 1.3 (0.86, 1.7) | 0.00028 | 33548\|19\|0 | AU |
| Weighted mean MD in tract superior thalamic radiation right - imaging visit - value 0 | 8.30E-09 | 1.3 (0.85, 1.7) | 0.00028 | 33548\|19\|0 | AU |
| Mean MD in posterior limb of internal capsule on FA skeleton right - imaging visit - value 0 | 1.20E-08 | 1.3 (0.85, 1.7) | 0.00028 | 33550\|19\|0 | AU |
| Mean MD in posterior corona radiata on FA skeleton right - imaging visit - value 0 | 1.30E-08 | 1.3 (0.83, 1.7) | 0.00028 | 33550\|19\|0 | AU |
| Mean L3 in posterior corona radiata on FA skeleton right - imaging visit - value 0 | 2.20E-08 | 1.3 (0.81, 1.7) | 0.00028 | 33550\|19\|0 | AU |
| Mean L3 in external capsule on FA skeleton left - imaging visit - value 0 | 2.40E-08 | 1.3 (0.82, 1.7) | 0.00028 | 33550\|19\|0 | AU |
| Mean ISOVF in superior corona radiata on FA skeleton right - imaging visit - value 0 | 2.40E-08 | 1.3 (0.82, 1.7) | 0.00028 | 33548\|19\|0 | AU |
| Mean L3 in superior corona radiata on FA skeleton right - imaging visit - value 0 | 2.50E-08 | 1.2 (0.81, 1.7) | 0.00028 | 33550\|19\|0 | AU |
| Mean MD in anterior limb of internal capsule on FA skeleton right - imaging visit - value 0 | 3.30E-08 | 1.2 (0.8, 1.7) | 0.00028 | 33550\|19\|0 | AU |
| Weighted mean L3 in tract superior thalamic radiation right - imaging visit - value 0 | 3.30E-08 | 1.2 (0.8, 1.7) | 0.00028 | 33548\|19\|0 | AU |
| Mean L2 in anterior limb of internal capsule on FA skeleton right - imaging visit - value 0 | 3.50E-08 | 1.2 (0.8, 1.7) | 0.00028 | 33550\|19\|0 | AU |
| Mean L1 in superior corona radiata on FA skeleton right - imaging visit - value 0 | 3.80E-08 | 1.2 (0.79, 1.7) | 0.00028 | 33550\|19\|0 | AU |
| Mean L3 in superior corona radiata on FA skeleton left - imaging visit - value 0 | 4.40E-08 | 1.2 (0.79, 1.7) | 0.00028 | 33550\|19\|0 | AU |
| Mean L3 in posterior limb of internal capsule on FA skeleton right - imaging visit - value 0 | 7.60E-08 | 1.2 (0.77, 1.7) | 0.00028 | 33550\|19\|0 | AU |
| Mean L2 in anterior limb of internal capsule on FA skeleton left - imaging visit - value 0 | 9.60E-08 | 1.2 (0.76, 1.6) | 0.00028 | 33550\|19\|0 | AU |
| Volume of WM hypointensities whole brain - imaging visit - value 0 | 1.10E-07 | 1.1 (0.72, 1.6) | 0.00029 | 35921\|21\|0 | $mm^3$ |
| Weighted mean L2 in tract superior thalamic radiation left - imaging visit - value 0 | 1.40E-07 | 1.2 (0.74, 1.6) | 0.00028 | 33548\|19\|0 | AU |

| Phenotype | P value | Beta (95% CI) | AAF | Cases (RR\|RA\|AA) | Units |
|---|---|---|---|---|---|
| Mean L2 in posterior corona radiata on FA skeleton right - imaging visit - value 0 | 1.50E-07 | 1.2 (0.74, 1.6) | 0.00028 | 33550\|19\|0 | AU |
| Weighted mean L2 in tract superior thalamic radiation right - imaging visit - value 0 | 1.60E-07 | 1.2 (0.74, 1.6) | 0.00028 | 33548\|19\|0 | AU |
| Mean L1 in external capsule on FA skeleton right - imaging visit - value 0 | 1.60E-07 | 1.2 (0.74, 1.6) | 0.00028 | 33550\|19\|0 | AU |
| Mean L1 in superior corona radiata on FA skeleton left - imaging visit - value 0 | 1.70E-07 | 1.2 (0.73, 1.6) | 0.00028 | 33550\|19\|0 | AU |
| Weighted mean L2 in tract corticospinal tract right - imaging visit - value 0 | 2.20E-07 | 1.2 (0.73, 1.6) | 0.00028 | 33548\|19\|0 | AU |
| Weighted mean L3 in tract corticospinal tract right - imaging visit - value 0 | 2.80E-07 | 1.2 (0.72, 1.6) | 0.00028 | 33548\|19\|0 | AU |
| Mean ICVF in external capsule on FA skeleton left - imaging visit - value 0 | 3.20E-07 | -1.1 (-1.6, -0.7) | 0.00028 | 33548\|19\|0 | AU |
| Mean FA in anterior limb of internal capsule on FA skeleton left - imaging visit - value 0 | 3.50E-07 | -1.1 (-1.6, -0.7) | 0.00028 | 33550\|19\|0 | AU |
| Weighted mean L3 in tract anterior thalamic radiation right - imaging visit - value 0 | 3.80E-07 | 1.1 (0.7, 1.6) | 0.00028 | 33548\|19\|0 | AU |
| Mean L3 in anterior limb of internal capsule on FA skeleton left - imaging visit - value 0 | 5.40E-07 | 1.1 (0.69, 1.6) | 0.00028 | 33550\|19\|0 | AU |
| Weighted mean MD in tract anterior thalamic radiation right - imaging visit - value 0 | 5.70E-07 | 1.1 (0.69, 1.6) | 0.00028 | 33548\|19\|0 | AU |
| Mean MD in anterior limb of internal capsule on FA skeleton left - imaging visit - value 0 | 6.90E-07 | 1.1 (0.68, 1.6) | 0.00028 | 33550\|19\|0 | AU |
| Mean L3 in anterior limb of internal capsule on FA skeleton right - imaging visit - value 0 | 7.00E-07 | 1.1 (0.68, 1.6) | 0.00028 | 33550\|19\|0 | AU |
| Mean MD in posterior corona radiata on FA skeleton left - imaging visit - value 0 | 7.00E-07 | 1.1 (0.67, 1.5) | 0.00028 | 33550\|19\|0 | AU |
| Mean L2 in posterior corona radiata on FA skeleton left - imaging visit - value 0 | 7.30E-07 | 1.1 (0.67, 1.6) | 0.00028 | 33550\|19\|0 | AU |
| Mean L2 in superior corona radiata on FA skeleton right - imaging visit - value 0 | 7.70E-07 | 1.1 (0.67, 1.5) | 0.00028 | 33550\|19\|0 | AU |
| Mean ISOVF in external capsule on FA skeleton right - imaging visit - value 0 | 8.40E-07 | 1.1 (0.67, 1.6) | 0.00028 | 33548\|19\|0 | AU |
| Mean ICVF in external capsule on FA skeleton right - imaging visit - value 0 | 8.50E-07 | -1.1 (-1.5, -0.66) | 0.00028 | 33548\|19\|0 | AU |
| Mean L1 in posterior corona radiata on FA skeleton right - imaging visit - value 0 | 9.40E-07 | 1.1 (0.66, 1.5) | 0.00028 | 33550\|19\|0 | AU |
| Weighted mean ISOVF in tract superior thalamic radiation right - imaging visit - value 0 | 1.10E-06 | 1.1 (0.66, 1.5) | 0.00028 | 33547\|19\|0 | AU |
| Mean FA in anterior limb of internal capsule on FA skeleton right - imaging visit - value 0 | 1.20E-06 | -1.1 (-1.5, -0.65) | 0.00028 | 33550\|19\|0 | AU |
| Mean FA in external capsule on FA skeleton right - imaging visit - value 0 | 1.20E-06 | -1.1 (-1.5, -0.65) | 0.00028 | 33550\|19\|0 | AU |
| Mean MD in superior fronto occipital fasciculus on FA skeleton left - imaging visit - value 0 | 1.50E-06 | 1.1 (0.64, 1.5) | 0.00028 | 33550\|19\|0 | AU |
| Mean L1 in external capsule on FA skeleton left - imaging visit - value 0 | 1.60E-06 | 1.1 (0.64, 1.5) | 0.00028 | 33550\|19\|0 | AU |
| Weighted mean L1 in tract superior thalamic radiation right - imaging visit - value 0 | 1.60E-06 | 1.1 (0.63, 1.5) | 0.00028 | 33548\|19\|0 | AU |
| Mean ICVF in posterior corona radiata on FA skeleton right - imaging visit - value 0 | 1.70E-06 | -1.1 (-1.5, -0.63) | 0.00028 | 33548\|19\|0 | AU |
| Weighted mean MD in tract corticospinal tract right - imaging visit - value 0 | 1.90E-06 | 1.1 (0.64, 1.5) | 0.00028 | 33548\|19\|0 | AU |

| Phenotype | P value | Beta (95% CI) | AAF | Cases (RR\|RA\|AA) | Units |
|---|---|---|---|---|---|
| Mean ICVF in superior corona radiata on FA skeleton left - imaging visit - value 0 | 2.10E-06 | -1 (-1.5, -0.61) | 0.00028 | 33548\|19\|0 | AU |
| Mean L3 in posterior corona radiata on FA skeleton left - imaging visit - value 0 | 2.20E-06 | 1.1 (0.62, 1.5) | 0.00028 | 33550\|19\|0 | AU |
| Volume of grey matter in Caudate right - imaging visit - value 0 | 2.20E-06 | 1 (0.61, 1.5) | 0.00028 | 35591\|20\|0 | $mm^3$ |
| Mean ISOVF in superior corona radiata on FA skeleton left - imaging visit - value 0 | 2.30E-06 | 1.1 (0.63, 1.5) | 0.00028 | 33548\|19\|0 | AU |
| Mean MD in posterior limb of internal capsule on FA skeleton left - imaging visit - value 0 | 2.40E-06 | 1.1 (0.62, 1.5) | 0.00028 | 33550\|19\|0 | AU |
| Weighted mean L1 in tract anterior thalamic radiation right - imaging visit - value 0 | 2.60E-06 | 1.1 (0.62, 1.5) | 0.00028 | 33548\|19\|0 | AU |
| Mean FA in external capsule on FA skeleton left - imaging visit - value 0 | 2.70E-06 | -1.1 (-1.5, -0.61) | 0.00028 | 33550\|19\|0 | AU |
| Total volume of white matter hyperintensities from T1 and T2 FLAIR images - imaging visit - value 0[2] | 3.00E-06 | 1.1 (0.61, 1.5) | 0.00028 | 34354\|19\|0 | $mm^3$ |
| Mean ISOVF in external capsule on FA skeleton left - imaging visit - value 0 | 3.80E-06 | 1 (0.6, 1.5) | 0.00028 | 33548\|19\|0 | AU |
| Weighted mean L3 in tract corticospinal tract left - imaging visit - value 0 | 5.00E-06 | 1 (0.59, 1.5) | 0.00028 | 33548\|19\|0 | AU |
| Weighted mean L1 in tract superior thalamic radiation left - imaging visit - value 0 | 5.40E-06 | 1 (0.58, 1.5) | 0.00028 | 33548\|19\|0 | AU |
| Mean FA in posterior corona radiata on FA skeleton right - imaging visit - value 0 | 5.40E-06 | -1 (-1.5, -0.58) | 0.00028 | 33550\|19\|0 | AU |
| Weighted mean ISOVF in tract superior thalamic radiation left - imaging visit - value 0 | 5.50E-06 | 1 (0.58, 1.5) | 0.00028 | 33547\|19\|0 | AU |
| Mean ICVF in superior corona radiata on FA skeleton right - imaging visit - value 0 | 5.60E-06 | -1 (-1.4, -0.57) | 0.00028 | 33548\|19\|0 | AU |
| Mean L3 in superior fronto occipital fasciculus on FA skeleton left - imaging visit - value 0 | 8.00E-06 | 1 (0.57, 1.4) | 0.00028 | 33550\|19\|0 | AU |
| Mean L3 in posterior limb of internal capsule on FA skeleton left - imaging visit - value 0 | 8.40E-06 | 1 (0.56, 1.4) | 0.00028 | 33550\|19\|0 | AU |
| Weighted mean L2 in tract anterior thalamic radiation right - imaging visit - value 0 | 9.10E-06 | 1 (0.56, 1.4) | 0.00028 | 33548\|19\|0 | AU |
| Weighted mean MD in tract corticospinal tract left - imaging visit - value 0 | 1.10E-05 | 1 (0.55, 1.4) | 0.00028 | 33548\|19\|0 | AU |
| Mean ISOVF in anterior limb of internal capsule on FA skeleton right - imaging visit - value 0 | 1.20E-05 | 0.99 (0.55, 1.4) | 0.00028 | 33548\|19\|0 | AU |
| Mean L2 in posterior thalamic radiation on FA skeleton left - imaging visit - value 0 | 1.20E-05 | 0.98 (0.54, 1.4) | 0.00028 | 33550\|19\|0 | AU |
| Volume of grey matter in Putamen left - imaging visit - value 0 | 1.50E-05 | 0.94 (0.52, 1.4) | 0.00028 | 35591\|20\|0 | $mm^3$ |
| Mean MD in superior fronto occipital fasciculus on FA skeleton right - imaging visit - value 0 | 1.50E-05 | 0.98 (0.54, 1.4) | 0.00028 | 33550\|19\|0 | AU |
| Weighted mean L3 in tract anterior thalamic radiation left - imaging visit - value 0 | 1.60E-05 | 0.97 (0.53, 1.4) | 0.00028 | 33548\|19\|0 | AU |
| Volume of grey matter in Caudate left - imaging visit - value 0 | 1.60E-05 | 0.94 (0.51, 1.4) | 0.00028 | 35591\|20\|0 | $mm^3$ |
| Mean ISOVF in posterior limb of internal capsule on FA skeleton right - imaging visit - value 0 | 1.60E-05 | 0.98 (0.54, 1.4) | 0.00028 | 33548\|19\|0 | AU |
| Volume of AV right hemisphere - imaging visit - value 0 | 1.70E-05 | 0.92 (0.5, 1.3) | 0.00029 | 35921\|21\|0 | $mm^3$ |
| Mean L2 in superior corona radiata on FA skeleton left - imaging visit - value 0 | 1.80E-05 | 0.96 (0.52, 1.4) | 0.00028 | 33550\|19\|0 | AU |

| Phenotype | P value | Beta (95% CI) | AAF | Cases (RR\|RA\|AA) | Units |
|---|---|---|---|---|---|
| Mean L1 in posterior corona radiata on FA skeleton left - imaging visit - value 0 | 1.80E-05 | 0.95 (0.52, 1.4) | 0.00028 | 33550\|19\|0 | AU |
| Weighted mean FA in tract superior thalamic radiation left - imaging visit - value 0 | 2.00E-05 | -0.96 (-1.4, -0.52) | 0.00028 | 33548\|19\|0 | AU |
| Weighted mean MD in tract anterior thalamic radiation left - imaging visit - value 0 | 2.00E-05 | 0.96 (0.52, 1.4) | 0.00028 | 33548\|19\|0 | AU |
| Mean ISOVF in posterior corona radiata on FA skeleton right - imaging visit - value 0 | 2.00E-05 | 0.97 (0.52, 1.4) | 0.00028 | 33548\|19\|0 | AU |
| Mean L2 in superior fronto occipital fasciculus on FA skeleton right - imaging visit - value 0 | 2.00E-05 | 0.97 (0.52, 1.4) | 0.00028 | 33550\|19\|0 | AU |
| Mean L2 in retrolenticular part of internal capsule on FA skeleton left - imaging visit - value 0 | 2.40E-05 | 0.95 (0.51, 1.4) | 0.00028 | 33550\|19\|0 | AU |
| Mean L3 in superior fronto occipital fasciculus on FA skeleton right - imaging visit - value 0 | 2.50E-05 | 0.95 (0.51, 1.4) | 0.00028 | 33550\|19\|0 | AU |
| Mean L1 in superior fronto occipital fasciculus on FA skeleton left - imaging visit - value 0 | 2.80E-05 | 0.95 (0.51, 1.4) | 0.00028 | 33550\|19\|0 | AU |
| Mean intensity of WM hypointensities whole brain - imaging visit - value 0 | 2.90E-05 | 0.91 (0.48, 1.3) | 0.00029 | 35921\|21\|0 | AU |
| Mean MD in posterior thalamic radiation on FA skeleton left - imaging visit - value 0 | 3.00E-05 | 0.93 (0.49, 1.4) | 0.00028 | 33550\|19\|0 | AU |
| Weighted mean L2 in tract anterior thalamic radiation left - imaging visit - value 0 | 3.30E-05 | 0.93 (0.49, 1.4) | 0.00028 | 33548\|19\|0 | AU |
| Mean L3 in posterior thalamic radiation on FA skeleton left - imaging visit - value 0 | 3.50E-05 | 0.92 (0.49, 1.4) | 0.00028 | 33550\|19\|0 | AU |
| Weighted mean L2 in tract posterior thalamic radiation left - imaging visit - value 0 | 3.60E-05 | 0.93 (0.49, 1.4) | 0.00028 | 33548\|19\|0 | AU |
| Mean L3 in anterior corona radiata on FA skeleton right - imaging visit - value 0 | 4.20E-05 | 0.91 (0.48, 1.3) | 0.00028 | 33550\|19\|0 | AU |
| Weighted mean ICVF in tract superior thalamic radiation left - imaging visit - value 0 | 4.20E-05 | -0.91 (-1.3, -0.47) | 0.00028 | 33547\|19\|0 | AU |
| Weighted mean FA in tract anterior thalamic radiation right - imaging visit - value 0 | 4.20E-05 | -0.92 (-1.4, -0.48) | 0.00028 | 33548\|19\|0 | AU |
| Weighted mean FA in tract anterior thalamic radiation left - imaging visit - value 0 | 4.50E-05 | -0.92 (-1.4, -0.48) | 0.00028 | 33548\|19\|0 | AU |
| Weighted mean FA in tract corticospinal tract right - imaging visit - value 0 | 4.50E-05 | -0.92 (-1.4, -0.48) | 0.00028 | 33548\|19\|0 | AU |
| Mean L2 in posterior thalamic radiation on FA skeleton right - imaging visit - value 0 | 4.80E-05 | 0.91 (0.47, 1.4) | 0.00028 | 33550\|19\|0 | AU |
| Weighted mean MD in tract posterior thalamic radiation left - imaging visit - value 0 | 5.50E-05 | 0.91 (0.47, 1.4) | 0.00028 | 33548\|19\|0 | AU |
| Mean L3 in posterior thalamic radiation on FA skeleton right - imaging visit - value 0 | 5.80E-05 | 0.9 (0.46, 1.3) | 0.00028 | 33550\|19\|0 | AU |
| Mean FA in posterior thalamic radiation on FA skeleton left - imaging visit - value 0 | 6.20E-05 | -0.9 (-1.3, -0.46) | 0.00028 | 33550\|19\|0 | AU |
| Mean MD in posterior thalamic radiation on FA skeleton right - imaging visit - value 0 | 6.40E-05 | 0.89 (0.46, 1.3) | 0.00028 | 33550\|19\|0 | AU |
| Mean ISOVF in posterior corona radiata on FA skeleton left - imaging visit - value 0 | 6.50E-05 | 0.9 (0.46, 1.3) | 0.00028 | 33548\|19\|0 | AU |
| Mean MD in retrolenticular part of internal capsule on FA skeleton left - imaging visit - value 0 | 6.50E-05 | 0.9 (0.46, 1.3) | 0.00028 | 33550\|19\|0 | AU |
| Mean ICVF in superior fronto occipital fasciculus on FA skeleton left - imaging visit - value 0 | 6.50E-05 | -0.89 (-1.3, -0.45) | 0.00028 | 33548\|19\|0 | AU |
| Weighted mean FA in tract superior thalamic radiation right - imaging visit - value 0 | 6.90E-05 | -0.89 (-1.3, -0.45) | 0.00028 | 33548\|19\|0 | AU |

| Phenotype | P value | Beta (95% CI) | AAF | Cases (RR\|RA\|AA) | Units |
|---|---|---|---|---|---|
| Mean MD in anterior corona radiata on FA skeleton right - imaging visit - value 0 | 7.30E-05 | 0.88 (0.45, 1.3) | 0.00028 | 33550\|19\|0 | AU |
| Weighted mean MD in tract inferior fronto occipital fasciculus right - imaging visit - value 0 | 7.50E-05 | 0.88 (0.45, 1.3) | 0.00028 | 33548\|19\|0 | AU |
| Mean FA in posterior corona radiata on FA skeleton left - imaging visit - value 0 | 7.60E-05 | -0.89 (-1.3, -0.45) | 0.00028 | 33550\|19\|0 | AU |
| Mean MD in tapetum on FA skeleton left - imaging visit - value 0 | 7.90E-05 | 0.89 (0.45, 1.3) | 0.00028 | 33550\|19\|0 | AU |
| Weighted mean L3 in tract posterior thalamic radiation left - imaging visit - value 0 | 9.10E-05 | 0.88 (0.44, 1.3) | 0.00028 | 33548\|19\|0 | AU |
| Weighted mean FA in tract posterior thalamic radiation left - imaging visit - value 0 | 9.20E-05 | -0.88 (-1.3, -0.44) | 0.00028 | 33548\|19\|0 | AU |
| Weighted mean L2 in tract superior longitudinal fasciculus right - imaging visit - value 0 | 9.30E-05 | 0.87 (0.43, 1.3) | 0.00028 | 33548\|19\|0 | AU |
| Volume of grey matter in Thalamus right - imaging visit - value 0 | 9.80E-05 | 0.85 (0.42, 1.3) | 0.00028 | 35591\|20\|0 | $mm^3$ |
| Mean FA in posterior thalamic radiation on FA skeleton right - imaging visit - value 0 | 1.00E-04 | -0.87 (-1.3, -0.43) | 0.00028 | 33550\|19\|0 | AU |
| Weighted mean ICVF in tract superior thalamic radiation right - imaging visit - value 0 | 1.00E-04 | -0.86 (-1.3, -0.42) | 0.00028 | 33547\|19\|0 | AU |
| Mean MD in retrolenticular part of internal capsule on FA skeleton right - imaging visit - value 0 | 0.00011 | 0.87 (0.43, 1.3) | 0.00028 | 33550\|19\|0 | AU |
| Weighted mean ICVF in tract corticospinal tract right - imaging visit - value 0 | 0.00011 | -0.86 (-1.3, -0.42) | 0.00028 | 33547\|19\|0 | AU |
| 3mm index of best keratometry results right - initial visit - mean | 0.00012 | -0.47 (-0.7, -0.23) | 0.00022 | 90674\|40\|0 | no_registered_unit |
| Mean L3 in sagittal stratum on FA skeleton right - imaging visit - value 0 | 0.00012 | 0.86 (0.42, 1.3) | 0.00028 | 33550\|19\|0 | AU |
| Mean MD in sagittal stratum on FA skeleton right - imaging visit - value 0 | 0.00013 | 0.86 (0.42, 1.3) | 0.00028 | 33550\|19\|0 | AU |
| Mean MD in body of corpus callosum on FA skeleton - imaging visit - value 0 | 0.00013 | 0.85 (0.42, 1.3) | 0.00028 | 33550\|19\|0 | AU |
| Volume of grey matter in Putamen right - imaging visit - value 0 | 0.00014 | 0.83 (0.4, 1.3) | 0.00028 | 35591\|20\|0 | $mm^3$ |
| Mean L2 in superior fronto occipital fasciculus on FA skeleton left - imaging visit - value 0 | 0.00014 | 0.86 (0.42, 1.3) | 0.00028 | 33550\|19\|0 | AU |
| Mean L3 in tapetum on FA skeleton left - imaging visit - value 0 | 0.00015 | 0.86 (0.41, 1.3) | 0.00028 | 33550\|19\|0 | AU |
| Mean ICVF in posterior corona radiata on FA skeleton left - imaging visit - value 0 | 0.00015 | -0.84 (-1.3, -0.41) | 0.00028 | 33548\|19\|0 | AU |
| Mean L3 in retrolenticular part of internal capsule on FA skeleton right - imaging visit - value 0 | 0.00015 | 0.85 (0.41, 1.3) | 0.00028 | 33550\|19\|0 | AU |
| Weighted mean L1 in tract anterior thalamic radiation left - imaging visit - value 0 | 0.00016 | 0.85 (0.41, 1.3) | 0.00028 | 33548\|19\|0 | AU |
| Weighted mean L3 in tract inferior fronto occipital fasciculus right - imaging visit - value 0 | 0.00016 | 0.84 (0.4, 1.3) | 0.00028 | 33548\|19\|0 | AU |
| Mean L2 in retrolenticular part of internal capsule on FA skeleton right - imaging visit - value 0 | 0.00018 | 0.84 (0.4, 1.3) | 0.00028 | 33550\|19\|0 | AU |
| Mean L3 in retrolenticular part of internal capsule on FA skeleton left - imaging visit - value 0 | 0.00018 | 0.84 (0.4, 1.3) | 0.00028 | 33550\|19\|0 | AU |
| Weighted mean L2 in tract inferior fronto occipital fasciculus right - imaging visit - value 0 | 0.00019 | 0.83 (0.39, 1.3) | 0.00028 | 33548\|19\|0 | AU |
| Mean ICVF in tapetum on FA skeleton right - imaging visit - value 0 | 2.00E-04 | -0.84 (-1.3, -0.39) | 0.00028 | 33548\|19\|0 | AU |

| Phenotype | P value | Beta (95% CI) | AAF | Cases (RR\|RA\|AA) | Units |
|---|---|---|---|---|---|
| Weighted mean L2 in tract corticospinal tract left - imaging visit - value 0 | 0.00021 | 0.84 (0.39, 1.3) | 0.00028 | 33548\|19\|0 | AU |
| Weighted mean ICVF in tract posterior thalamic radiation left - imaging visit - value 0 | 0.00025 | -0.81 (-1.2, -0.38) | 0.00028 | 33547\|19\|0 | AU |
| Mean ISOVF in retrolenticular part of internal capsule on FA skeleton left - imaging visit - value 0 | 0.00026 | 0.83 (0.38, 1.3) | 0.00028 | 33548\|19\|0 | AU |
| Weighted mean L2 in tract inferior fronto occipital fasciculus left - imaging visit - value 0 | 0.00027 | 0.81 (0.38, 1.2) | 0.00028 | 33548\|19\|0 | AU |
| Volume of grey matter in Thalamus left - imaging visit - value 0 | 0.00027 | 0.8 (0.37, 1.2) | 0.00028 | 35591\|20\|0 | $mm^3$ |
| Weighted mean FA in tract corticospinal tract left - imaging visit - value 0 | 0.00029 | -0.82 (-1.3, -0.38) | 0.00028 | 33548\|19\|0 | AU |
| Mean L2 in sagittal stratum on FA skeleton right - imaging visit - value 0 | 3.00E-04 | 0.81 (0.37, 1.3) | 0.00028 | 33550\|19\|0 | AU |
| 3mm index of best keratometry results right - mean of all visits | 3.00E-04 | -0.42 (-0.65, -0.19) | 0.00021 | 103404\|44\|0 | no_registered_unit |
| Mean L2 in anterior corona radiata on FA skeleton right - imaging visit - value 0 | 0.00031 | 0.8 (0.37, 1.2) | 0.00028 | 33550\|19\|0 | AU |
| Mean L2 in superior longitudinal fasciculus on FA skeleton right - imaging visit - value 0 | 0.00032 | 0.81 (0.37, 1.2) | 0.00028 | 33550\|19\|0 | AU |
| Mean ICVF in superior fronto occipital fasciculus on FA skeleton right - imaging visit - value 0 | 0.00036 | -0.8 (-1.2, -0.36) | 0.00028 | 33548\|19\|0 | AU |
| Weighted mean L1 in tract inferior fronto occipital fasciculus right - imaging visit - value 0 | 0.00038 | 0.8 (0.36, 1.2) | 0.00028 | 33548\|19\|0 | AU |
| Mean MD in superior longitudinal fasciculus on FA skeleton left - imaging visit - value 0 | 0.00039 | 0.79 (0.35, 1.2) | 0.00028 | 33550\|19\|0 | AU |
| Weighted mean MD in tract inferior fronto occipital fasciculus left - imaging visit - value 0 | 4.00E-04 | 0.79 (0.35, 1.2) | 0.00028 | 33548\|19\|0 | AU |
| Weighted mean ICVF in tract anterior thalamic radiation right - imaging visit - value 0 | 0.00041 | -0.78 (-1.2, -0.35) | 0.00028 | 33547\|19\|0 | AU |
| Mean FA in sagittal stratum on FA skeleton right - imaging visit - value 0 | 0.00045 | -0.79 (-1.2, -0.35) | 0.00028 | 33550\|19\|0 | AU |
| Mean FA in retrolenticular part of internal capsule on FA skeleton left - imaging visit - value 0 | 0.00046 | -0.79 (-1.2, -0.35) | 0.00028 | 33550\|19\|0 | AU |
| Mean L3 in body of corpus callosum on FA skeleton - imaging visit - value 0 | 0.00047 | 0.78 (0.34, 1.2) | 0.00028 | 33550\|19\|0 | AU |
| Mean ICVF in posterior thalamic radiation on FA skeleton left - imaging visit - value 0 | 0.00047 | -0.77 (-1.2, -0.34) | 0.00028 | 33548\|19\|0 | AU |
| Mean L1 in tapetum on FA skeleton left - imaging visit - value 0 | 0.00053 | 0.78 (0.34, 1.2) | 0.00028 | 33550\|19\|0 | AU |
| Mean ICVF in anterior limb of internal capsule on FA skeleton right - imaging visit - value 0 | 0.00054 | -0.77 (-1.2, -0.33) | 0.00028 | 33548\|19\|0 | AU |
| Weighted mean MD in tract superior longitudinal fasciculus right - imaging visit - value 0 | 6.00E-04 | 0.76 (0.33, 1.2) | 0.00028 | 33548\|19\|0 | AU |
| Mean FA in anterior corona radiata on FA skeleton right - imaging visit - value 0 | 0.00064 | -0.76 (-1.2, -0.32) | 0.00028 | 33550\|19\|0 | AU |
| Mean MD in anterior corona radiata on FA skeleton left - imaging visit - value 0 | 0.00064 | 0.76 (0.32, 1.2) | 0.00028 | 33550\|19\|0 | AU |
| Weighted mean FA in tract medial lemniscus right - imaging visit - value 0 | 7.00E-04 | -0.77 (-1.2, -0.32) | 0.00028 | 33548\|19\|0 | AU |
| Mean L2 in posterior limb of internal capsule on FA skeleton right - imaging visit - value 0 | 0.00072 | 0.76 (0.32, 1.2) | 0.00028 | 33550\|19\|0 | AU |
| Mean ICVF in anterior limb of internal capsule on FA skeleton left - imaging visit - value 0 | 0.00072 | -0.75 (-1.2, -0.32) | 0.00028 | 33548\|19\|0 | AU |

| Phenotype | P value | Beta (95% CI) | AAF | Cases (RR\|RA\|AA) | Units |
|---|---|---|---|---|---|
| Mean FA in retrolenticular part of internal capsule on FA skeleton right - imaging visit - value 0 | 0.00076 | -0.76 (-1.2, -0.32) | 0.00028 | 33550\|19\|0 | AU |
| Mean ISOVF in anterior limb of internal capsule on FA skeleton left - imaging visit - value 0 | 0.00077 | 0.76 (0.32, 1.2) | 0.00028 | 33548\|19\|0 | AU |
| Mean ICVF in tapetum on FA skeleton left - imaging visit - value 0 | 8.00E-04 | -0.76 (-1.2, -0.32) | 0.00028 | 33548\|19\|0 | AU |
| Mean ISOVF in body of corpus callosum on FA skeleton - imaging visit - value 0 | 8.00E-04 | 0.76 (0.31, 1.2) | 0.00028 | 33548\|19\|0 | AU |
| Mean intensity of Thalamus Proper right hemisphere - imaging visit - value 0 | 0.00082 | -0.72 (-1.1, -0.3) | 0.00029 | 35920\|21\|0 | AU |
| Mean L3 in anterior corona radiata on FA skeleton left - imaging visit - value 0 | 0.00082 | 0.74 (0.31, 1.2) | 0.00028 | 33550\|19\|0 | AU |
| Volume of hippocampal fissure right hemisphere - imaging visit - value 0 | 0.00082 | 0.72 (0.3, 1.1) | 0.00029 | 35921\|21\|0 | $mm^3$ |
| Weighted mean ICVF in tract anterior thalamic radiation left - imaging visit - value 0 | 0.00082 | -0.74 (-1.2, -0.31) | 0.00028 | 33547\|19\|0 | AU |
| Weighted mean L2 in tract inferior longitudinal fasciculus left - imaging visit - value 0 | 0.00085 | 0.74 (0.31, 1.2) | 0.00028 | 33548\|19\|0 | AU |
| Weighted mean MD in tract posterior thalamic radiation right - imaging visit - value 0 | 0.00086 | 0.75 (0.31, 1.2) | 0.00028 | 33548\|19\|0 | AU |
| Volume of Putamen left hemisphere - imaging visit - value 0 | 0.00087 | 0.71 (0.29, 1.1) | 0.00029 | 35921\|21\|0 | $mm^3$ |
| Weighted mean MD in tract inferior longitudinal fasciculus left - imaging visit - value 0 | 0.00091 | 0.74 (0.3, 1.2) | 0.00028 | 33548\|19\|0 | AU |
| Mean L2 in tapetum on FA skeleton left - imaging visit - value 0 | 0.00093 | 0.75 (0.31, 1.2) | 0.00028 | 33550\|19\|0 | AU |
| Weighted mean L3 in tract posterior thalamic radiation right - imaging visit - value 0 | 0.00094 | 0.75 (0.3, 1.2) | 0.00028 | 33548\|19\|0 | AU |
| Mean L1 in anterior corona radiata on FA skeleton left - imaging visit - value 0 | 0.00099 | 0.74 (0.3, 1.2) | 0.00028 | 33550\|19\|0 | AU |
| Mean MD in superior longitudinal fasciculus on FA skeleton right - imaging visit - value 0 | 0.001 | 0.73 (0.29, 1.2) | 0.00028 | 33550\|19\|0 | AU |
| Weighted mean L3 in tract superior longitudinal fasciculus right - imaging visit - value 0 | 0.0011 | 0.73 (0.29, 1.2) | 0.00028 | 33548\|19\|0 | AU |
| Mean L1 in anterior limb of internal capsule on FA skeleton right - imaging visit - value 0 | 0.0012 | 0.73 (0.29, 1.2) | 0.00028 | 33550\|19\|0 | AU |
| Mean ICVF in posterior thalamic radiation on FA skeleton right - imaging visit - value 0 | 0.0012 | -0.71 (-1.1, -0.28) | 0.00028 | 33548\|19\|0 | AU |
| Mean ISOVF in superior fronto occipital fasciculus on FA skeleton left - imaging visit - value 0 | 0.0012 | 0.74 (0.29, 1.2) | 0.00028 | 33548\|19\|0 | AU |
| Mean L1 in superior fronto occipital fasciculus on FA skeleton right - imaging visit - value 0 | 0.0013 | 0.73 (0.29, 1.2) | 0.00028 | 33550\|19\|0 | AU |
| Mean OD in middle cerebellar peduncle on FA skeleton - imaging visit - value 0 | 0.0013 | 0.73 (0.29, 1.2) | 0.00028 | 33548\|19\|0 | AU |
| Weighted mean L2 in tract uncinate fasciculus left - imaging visit - value 0 | 0.0013 | 0.72 (0.28, 1.2) | 0.00028 | 33548\|19\|0 | AU |
| Mean FA in superior fronto occipital fasciculus on FA skeleton right - imaging visit - value 0 | 0.0014 | -0.72 (-1.2, -0.28) | 0.00028 | 33550\|19\|0 | AU |
| Mean ICVF in anterior corona radiata on FA skeleton right - imaging visit - value 0 | 0.0014 | -0.7 (-1.1, -0.27) | 0.00028 | 33548\|19\|0 | AU |
| Mean ICVF in uncinate fasciculus on FA skeleton right - imaging visit - value 0 | 0.0014 | -0.71 (-1.2, -0.28) | 0.00028 | 33548\|19\|0 | AU |
| Volume of Putamen right hemisphere - imaging visit - value 0 | 0.0014 | 0.68 (0.26, 1.1) | 0.00029 | 35921\|21\|0 | $mm^3$ |

| Phenotype | P value | Beta (95% CI) | AAF | Cases (RR\|RA\|AA) | Units |
|---|---|---|---|---|---|
| Mean L1 in posterior limb of internal capsule on FA skeleton right - imaging visit - value 0 | 0.0014 | 0.71 (0.28, 1.2) | 0.00028 | 33550\|19\|0 | AU |
| Mean L2 in superior longitudinal fasciculus on FA skeleton left - imaging visit - value 0 | 0.0015 | 0.71 (0.27, 1.2) | 0.00028 | 33550\|19\|0 | AU |
| Mean ISOVF in posterior limb of internal capsule on FA skeleton left - imaging visit - value 0 | 0.0015 | 0.72 (0.28, 1.2) | 0.00028 | 33548\|19\|0 | AU |
| Mean ICVF in sagittal stratum on FA skeleton right - imaging visit - value 0 | 0.0015 | -0.7 (-1.1, -0.27) | 0.00028 | 33548\|19\|0 | AU |
| Weighted mean ICVF in tract corticospinal tract left - imaging visit - value 0 | 0.0015 | -0.7 (-1.1, -0.27) | 0.00028 | 33547\|19\|0 | AU |
| Weighted mean L2 in tract superior longitudinal fasciculus left - imaging visit - value 0 | 0.0015 | 0.71 (0.27, 1.1) | 0.00028 | 33548\|19\|0 | AU |
| Weighted mean ICVF in tract posterior thalamic radiation right - imaging visit - value 0 | 0.0016 | -0.7 (-1.1, -0.27) | 0.00028 | 33547\|19\|0 | AU |
| Weighted mean L3 in tract inferior fronto occipital fasciculus left - imaging visit - value 0 | 0.0016 | 0.7 (0.27, 1.1) | 0.00028 | 33548\|19\|0 | AU |
| Mean L3 in uncinate fasciculus on FA skeleton right - imaging visit - value 0 | 0.0017 | 0.71 (0.27, 1.2) | 0.00028 | 33550\|19\|0 | AU |
| Mean L2 in body of corpus callosum on FA skeleton - imaging visit - value 0 | 0.0018 | 0.7 (0.26, 1.1) | 0.00028 | 33550\|19\|0 | AU |
| Volume of PuA left hemisphere - imaging visit - value 0 | 0.0018 | 0.67 (0.25, 1.1) | 0.00029 | 35921\|21\|0 | $mm^3$ |
| Grey white contrast in lingual left hemisphere - imaging visit - value 0 | 0.0018 | 0.66 (0.24, 1.1) | 0.00029 | 35921\|21\|0 | AU |
| Volume of CL right hemisphere - imaging visit - value 0 | 0.002 | 0.67 (0.24, 1.1) | 0.00029 | 35921\|21\|0 | $mm^3$ |
| Mean ISOVF in retrolenticular part of internal capsule on FA skeleton right - imaging visit - value 0 | 0.002 | 0.7 (0.26, 1.1) | 0.00028 | 33548\|19\|0 | AU |
| Area of S collat transv post left hemisphere - imaging visit - value 0 | 0.002 | 0.67 (0.25, 1.1) | 0.00029 | 35921\|21\|0 | $mm^2$ |
| Weighted mean FA in tract superior longitudinal fasciculus right - imaging visit - value 0 | 0.002 | -0.69 (-1.1, -0.25) | 0.00028 | 33548\|19\|0 | AU |
| Mean MD in fornix cres stria terminalis on FA skeleton left - imaging visit - value 0 | 0.0021 | 0.7 (0.25, 1.1) | 0.00028 | 33550\|19\|0 | AU |
| Weighted mean L1 in tract posterior thalamic radiation left - imaging visit - value 0 | 0.0021 | 0.7 (0.25, 1.1) | 0.00028 | 33548\|19\|0 | AU |
| Weighted mean L2 in tract inferior longitudinal fasciculus right - imaging visit - value 0 | 0.0021 | 0.68 (0.25, 1.1) | 0.00028 | 33548\|19\|0 | AU |
| Weighted mean L3 in tract inferior longitudinal fasciculus left - imaging visit - value 0 | 0.0023 | 0.68 (0.24, 1.1) | 0.00028 | 33548\|19\|0 | AU |
| Mean L1 in anterior corona radiata on FA skeleton right - imaging visit - value 0 | 0.0023 | 0.68 (0.24, 1.1) | 0.00028 | 33550\|19\|0 | AU |
| Weighted mean L2 in tract posterior thalamic radiation right - imaging visit - value 0 | 0.0023 | 0.69 (0.24, 1.1) | 0.00028 | 33548\|19\|0 | AU |
| Weighted mean ICVF in tract inferior fronto occipital fasciculus right - imaging visit - value 0 | 0.0023 | -0.67 (-1.1, -0.24) | 0.00028 | 33547\|19\|0 | AU |
| Mean MO in middle cerebellar peduncle on FA skeleton - imaging visit - value 0 | 0.0024 | -0.68 (-1.1, -0.24) | 0.00028 | 33550\|19\|0 | AU |
| Mean thickness of BA4p left hemisphere - imaging visit - value 0 | 0.0024 | -0.65 (-1.1, -0.23) | 0.00029 | 35921\|21\|0 | mm |
| Mean L3 in superior longitudinal fasciculus on FA skeleton right - imaging visit - value 0 | 0.0025 | 0.67 (0.24, 1.1) | 0.00028 | 33550\|19\|0 | AU |
| Mean corpuscular volume - mean of all visits | 0.0025 | -0.21 (-0.34, -0.073) | 0.0002 | 418705\|167\|0 | femtolitres |

| Phenotype | P value | Beta (95% CI) | AAF | Cases (RR\|RA\|AA) | Units |
|---|---|---|---|---|---|
| Weighted mean FA in tract inferior longitudinal fasciculus right - imaging visit - value 0 | 0.0025 | -0.67 (-1.1, -0.24) | 0.00028 | 33548\|19\|0 | AU |
| Mean FA in middle cerebellar peduncle on FA skeleton - imaging visit - value 0 | 0.0025 | -0.68 (-1.1, -0.24) | 0.00028 | 33550\|19\|0 | AU |
| Weighted mean L3 in tract inferior longitudinal fasciculus right - imaging visit - value 0 | 0.0026 | 0.67 (0.23, 1.1) | 0.00028 | 33548\|19\|0 | AU |
| Mean L2 in genu of corpus callosum on FA skeleton - imaging visit - value 0 | 0.0026 | 0.68 (0.24, 1.1) | 0.00028 | 33550\|19\|0 | AU |
| Mean FA in posterior limb of internal capsule on FA skeleton right - imaging visit - value 0 | 0.0027 | -0.67 (-1.1, -0.23) | 0.00028 | 33550\|19\|0 | AU |
| Volume of CL left hemisphere - imaging visit - value 0 | 0.0028 | 0.64 (0.22, 1.1) | 0.00029 | 35921\|21\|0 | $mm^3$ |
| Area of Pole temporal left hemisphere - imaging visit - value 0 | 0.0028 | 0.64 (0.22, 1.1) | 0.00029 | 35921\|21\|0 | $mm^2$ |
| Weighted mean FA in tract inferior fronto occipital fasciculus right - imaging visit - value 0 | 0.0029 | -0.67 (-1.1, -0.23) | 0.00028 | 33548\|19\|0 | AU |
| Weighted mean MD in tract inferior longitudinal fasciculus right - imaging visit - value 0 | 0.0029 | 0.66 (0.23, 1.1) | 0.00028 | 33548\|19\|0 | AU |
| Mean FA in superior fronto occipital fasciculus on FA skeleton left - imaging visit - value 0 | 0.0029 | -0.67 (-1.1, -0.23) | 0.00028 | 33550\|19\|0 | AU |
| Frequency of travelling from home to job workplace - initial visit - mean | 0.003 | 0.26 (0.087, 0.42) | 0.00021 | 240690\|100\|0 | times |
| Weighted mean L1 in tract inferior longitudinal fasciculus left - imaging visit - value 0 | 0.003 | 0.67 (0.23, 1.1) | 0.00028 | 33548\|19\|0 | AU |
| Mean thickness of insula left hemisphere - imaging visit - value 0 | 0.0031 | -0.64 (-1.1, -0.22) | 0.00029 | 35921\|21\|0 | mm |
| Weighted mean MD in tract superior longitudinal fasciculus left - imaging visit - value 0 | 0.0032 | 0.65 (0.22, 1.1) | 0.00028 | 33548\|19\|0 | AU |
| Volume of Lateral nucleus right hemisphere - imaging visit - value 0 | 0.0032 | 0.63 (0.21, 1.1) | 0.00029 | 35921\|21\|0 | $mm^3$ |
| Mean ISOVF in superior fronto occipital fasciculus on FA skeleton right - imaging visit - value 0 | 0.0032 | 0.67 (0.22, 1.1) | 0.00028 | 33548\|19\|0 | AU |
| Mean ISOVF in tapetum on FA skeleton left - imaging visit - value 0 | 0.0033 | 0.67 (0.22, 1.1) | 0.00028 | 33548\|19\|0 | AU |
| Volume of AV left hemisphere - imaging visit - value 0 | 0.0033 | 0.63 (0.21, 1) | 0.00029 | 35921\|21\|0 | $mm^3$ |
| Mean ICVF in superior longitudinal fasciculus on FA skeleton left - imaging visit - value 0 | 0.0033 | -0.65 (-1.1, -0.21) | 0.00028 | 33548\|19\|0 | AU |
| Volume of SubCortGray whole brain - imaging visit - value 0 | 0.0035 | 0.62 (0.21, 1) | 0.00029 | 35921\|21\|0 | $mm^3$ |
| Mean L2 in anterior corona radiata on FA skeleton left - imaging visit - value 0 | 0.0035 | 0.65 (0.22, 1.1) | 0.00028 | 33550\|19\|0 | AU |
| Volume of MV Re right hemisphere - imaging visit - value 0 | 0.0035 | 0.63 (0.21, 1.1) | 0.00029 | 35921\|21\|0 | $mm^3$ |
| Mean corpuscular volume - initial visit - mean | 0.0036 | -0.2 (-0.33, -0.065) | 0.0002 | 418058\|167\|0 | femtolitres |
| Mean FA in superior corona radiata on FA skeleton left - imaging visit - value 0 | 0.0036 | -0.66 (-1.1, -0.21) | 0.00028 | 33550\|19\|0 | AU |
| Weighted mean ISOVF in tract corticospinal tract left - imaging visit - value 0 | 0.0037 | 0.66 (0.21, 1.1) | 0.00028 | 33547\|19\|0 | AU |
| Volume of Whole amygdala right hemisphere - imaging visit - value 0 | 0.0037 | 0.62 (0.2, 1) | 0.00029 | 35921\|21\|0 | $mm^3$ |
| Volume of Caudate right hemisphere - imaging visit - value 0 | 0.0038 | 0.61 (0.2, 1) | 0.00029 | 35921\|21\|0 | $mm^3$ |

| Phenotype | P value | Beta (95% CI) | AAF | Cases (RR\|RA\|AA) | Units |
|---|---|---|---|---|---|
| Mean FA in genu of corpus callosum on FA skeleton - imaging visit - value 0 | 0.0038 | -0.65 (-1.1, -0.21) | 0.00028 | 33550\|19\|0 | AU |
| Mean L3 in genu of corpus callosum on FA skeleton - imaging visit - value 0 | 0.0039 | 0.64 (0.21, 1.1) | 0.00028 | 33550\|19\|0 | AU |
| Mean FA in superior longitudinal fasciculus on FA skeleton right - imaging visit - value 0 | 0.0039 | -0.64 (-1.1, -0.21) | 0.00028 | 33550\|19\|0 | AU |
| Mean FA in inferior cerebellar peduncle on FA skeleton left - imaging visit - value 0 | 0.0039 | -0.65 (-1.1, -0.21) | 0.00028 | 33550\|19\|0 | AU |
| Weighted mean L1 in tract acoustic radiation right - imaging visit - value 0 | 0.0041 | 0.65 (0.21, 1.1) | 0.00028 | 33548\|19\|0 | AU |
| Mean L3 in superior longitudinal fasciculus on FA skeleton left - imaging visit - value 0 | 0.0043 | 0.63 (0.2, 1.1) | 0.00028 | 33550\|19\|0 | AU |
| Weighted mean ISOVF in tract anterior thalamic radiation right - imaging visit - value 0 | 0.0043 | 0.65 (0.2, 1.1) | 0.00028 | 33547\|19\|0 | AU |
| Mean ICVF in posterior limb of internal capsule on FA skeleton right - imaging visit - value 0 | 0.0043 | -0.63 (-1.1, -0.2) | 0.00028 | 33548\|19\|0 | AU |
| Mean MD in genu of corpus callosum on FA skeleton - imaging visit - value 0 | 0.0044 | 0.64 (0.2, 1.1) | 0.00028 | 33550\|19\|0 | AU |
| Weighted mean ISOVF in tract corticospinal tract right - imaging visit - value 0 | 0.0045 | 0.64 (0.2, 1.1) | 0.00028 | 33547\|19\|0 | AU |
| Mean L1 in body of corpus callosum on FA skeleton - imaging visit - value 0 | 0.0046 | 0.64 (0.2, 1.1) | 0.00028 | 33550\|19\|0 | AU |
| Weighted mean L1 in tract inferior fronto occipital fasciculus left - imaging visit - value 0 | 0.0046 | 0.64 (0.2, 1.1) | 0.00028 | 33548\|19\|0 | AU |
| Weighted mean MD in tract uncinate fasciculus left - imaging visit - value 0 | 0.0046 | 0.64 (0.2, 1.1) | 0.00028 | 33548\|19\|0 | AU |
| Weighted mean ICVF in tract inferior fronto occipital fasciculus left - imaging visit - value 0 | 0.0047 | -0.62 (-1.1, -0.19) | 0.00028 | 33547\|19\|0 | AU |
| Mean corpuscular haemoglobin - initial visit - mean | 0.0048 | -0.19 (-0.33, -0.059) | 2.00E-04 | 418057\|167\|0 | picograms |
| Mean FA in superior corona radiata on FA skeleton right - imaging visit - value 0 | 0.0049 | -0.63 (-1.1, -0.19) | 0.00028 | 33550\|19\|0 | AU |
| Acceptability of each blow result - initial visit - mean | 0.0049 | 0.2 (0.059, 0.33) | 0.00021 | 392413\|161\|0 | no_registered_unit |
| Weighted mean ISOVF in tract superior longitudinal fasciculus right - imaging visit - value 0 | 0.0049 | 0.63 (0.19, 1.1) | 0.00028 | 33547\|19\|0 | AU |
| Mean L3 in tapetum on FA skeleton right - imaging visit - value 0 | 0.0049 | 0.63 (0.19, 1.1) | 0.00028 | 33550\|19\|0 | AU |
| Area of G S cingul Mid Post right hemisphere - imaging visit - value 0 | 0.005 | 0.61 (0.18, 1) | 0.00029 | 35921\|21\|0 | $mm^2$ |
| Mean corpuscular haemoglobin - mean of all visits | 0.0051 | -0.19 (-0.33, -0.057) | 2.00E-04 | 418702\|167\|0 | picograms |
| Weighted mean MD in tract acoustic radiation right - imaging visit - value 0 | 0.0053 | 0.64 (0.19, 1.1) | 0.00028 | 33548\|19\|0 | AU |
| Mean OD in posterior corona radiata on FA skeleton right - imaging visit - value 0 | 0.0054 | -0.62 (-1.1, -0.18) | 0.00028 | 33548\|19\|0 | AU |
| Mean intensity of Thalamus Proper left hemisphere - imaging visit - value 0 | 0.0054 | -0.6 (-1, -0.18) | 0.00029 | 35920\|21\|0 | AU |
| Weighted mean FA in tract posterior thalamic radiation right - imaging visit - value 0 | 0.0054 | -0.63 (-1.1, -0.18) | 0.00028 | 33548\|19\|0 | AU |
| Volume of CeM right hemisphere - imaging visit - value 0 | 0.0056 | 0.59 (0.17, 1) | 0.00029 | 35921\|21\|0 | $mm^3$ |
| Mean FA in medial lemniscus on FA skeleton left - imaging visit - value 0 | 0.0059 | -0.62 (-1.1, -0.18) | 0.00028 | 33550\|19\|0 | AU |

| Phenotype | P value | Beta (95% CI) | AAF | Cases (RR\|RA\|AA) | Units |
|---|---|---|---|---|---|
| Mean L1 in posterior limb of internal capsule on FA skeleton left - imaging visit - value 0 | 0.0059 | 0.62 (0.18, 1.1) | 0.00028 | 33550\|19\|0 | AU |
| Mean thickness of G parietal sup right hemisphere - imaging visit - value 0 | 0.006 | 0.59 (0.17, 1) | 0.00029 | 35921\|21\|0 | mm |
| Acceptability of each blow result - mean of all visits | 0.0062 | 0.19 (0.054, 0.33) | 2.00E-04 | 394984\|162\|0 | no_registered_unit |
| Weighted mean FA in tract inferior longitudinal fasciculus left - imaging visit - value 0 | 0.0062 | -0.61 (-1, -0.17) | 0.00028 | 33548\|19\|0 | AU |
| Weighted mean L3 in tract superior longitudinal fasciculus left - imaging visit - value 0 | 0.0063 | 0.61 (0.17, 1) | 0.00028 | 33548\|19\|0 | AU |
| Volume of hippocampal fissure left hemisphere - imaging visit - value 0 | 0.0064 | 0.59 (0.17, 1) | 0.00029 | 35921\|21\|0 | $mm^3$ |
| Weighted mean L3 in tract uncinate fasciculus left - imaging visit - value 0 | 0.0064 | 0.61 (0.17, 1.1) | 0.00028 | 33548\|19\|0 | AU |
| Mean OD in superior corona radiata on FA skeleton left - imaging visit - value 0 | 0.0066 | -0.61 (-1.1, -0.17) | 0.00028 | 33548\|19\|0 | AU |
| Mean ICVF in superior longitudinal fasciculus on FA skeleton right - imaging visit - value 0 | 0.0066 | -0.6 (-1, -0.17) | 0.00028 | 33548\|19\|0 | AU |
| Weighted mean FA in tract forceps major - imaging visit - value 0 | 0.0068 | -0.61 (-1.1, -0.17) | 0.00028 | 33548\|19\|0 | AU |
| Mean L2 in fornix cres stria terminalis on FA skeleton left - imaging visit - value 0 | 0.0068 | 0.61 (0.17, 1.1) | 0.00028 | 33550\|19\|0 | AU |
| Mean L3 in inferior cerebellar peduncle on FA skeleton left - imaging visit - value 0 | 0.0069 | 0.61 (0.17, 1.1) | 0.00028 | 33550\|19\|0 | AU |
| Weighted mean FA in tract inferior fronto occipital fasciculus left - imaging visit - value 0 | 0.0071 | -0.6 (-1, -0.16) | 0.00028 | 33548\|19\|0 | AU |
| Mean OD in superior corona radiata on FA skeleton right - imaging visit - value 0 | 0.0072 | -0.6 (-1, -0.16) | 0.00028 | 33548\|19\|0 | AU |
| Mean MD in tapetum on FA skeleton right - imaging visit - value 0 | 0.0074 | 0.6 (0.16, 1) | 0.00028 | 33550\|19\|0 | AU |
| Weighted mean ICVF in tract inferior longitudinal fasciculus left - imaging visit - value 0 | 0.0074 | -0.59 (-1, -0.16) | 0.00028 | 33547\|19\|0 | AU |
| Volume of BA4p left hemisphere - imaging visit - value 0 | 0.0076 | -0.58 (-1, -0.15) | 0.00029 | 35921\|21\|0 | $mm^3$ |
| Mean ICVF in anterior corona radiata on FA skeleton left - imaging visit - value 0 | 0.0076 | -0.59 (-1, -0.16) | 0.00028 | 33548\|19\|0 | AU |
| Volume of MDl right hemisphere - imaging visit - value 0 | 0.0078 | 0.57 (0.15, 1) | 0.00029 | 35921\|21\|0 | $mm^3$ |
| Mean thickness of insula left hemisphere - imaging visit - value 0 | 0.0078 | -0.58 (-1, -0.15) | 0.00029 | 35921\|21\|0 | mm |
| Weighted mean MO in tract anterior thalamic radiation right - imaging visit - value 0 | 0.0078 | 0.6 (0.16, 1) | 0.00028 | 33548\|19\|0 | AU |
| OS Contour 1 to 2 Region T6 Mean | 0.008 | -0.65 (-1.1, -0.17) | 0.00016 | 46458\|15\|0 | NA |
| Fathers age at death - initial visit - mean | 0.0081 | -0.23 (-0.39, -0.059) | 2.00E-04 | 317217\|129\|0 | years |
| Mean L2 in sagittal stratum on FA skeleton left - imaging visit - value 0 | 0.0083 | 0.59 (0.15, 1) | 0.00028 | 33550\|19\|0 | AU |
| Weighted mean FA in tract superior longitudinal fasciculus left - imaging visit - value 0 | 0.0083 | -0.59 (-1, -0.15) | 0.00028 | 33548\|19\|0 | AU |
| Volume of Caudate left hemisphere - imaging visit - value 0 | 0.0086 | 0.55 (0.14, 0.97) | 0.00029 | 35921\|21\|0 | $mm^3$ |
| Mean FA in tapetum on FA skeleton right - imaging visit - value 0 | 0.0086 | -0.59 (-1, -0.15) | 0.00028 | 33550\|19\|0 | AU |

| Phenotype | P value | Beta (95% CI) | AAF | Cases (RR\|RA\|AA) | Units |
|---|---|---|---|---|---|
| Area of posteriorcingulate right hemisphere - imaging visit - value 0 | 0.0088 | 0.57 (0.14, 0.99) | 0.00029 | 35921\|21\|0 | $mm^2$ |
| Volume of BA3a left hemisphere - imaging visit - value 0 | 0.0089 | -0.57 (-0.99, -0.14) | 0.00029 | 35921\|21\|0 | $mm^3$ |
| Mean L1 in posterior thalamic radiation on FA skeleton left - imaging visit - value 0 | 0.0089 | 0.59 (0.15, 1) | 0.00028 | 33550\|19\|0 | AU |
| Mean L2 in tapetum on FA skeleton right - imaging visit - value 0 | 0.009 | 0.59 (0.15, 1) | 0.00028 | 33550\|19\|0 | AU |
| Weighted mean ICVF in tract superior longitudinal fasciculus right - imaging visit - value 0 | 0.0092 | -0.57 (-1, -0.14) | 0.00028 | 33547\|19\|0 | AU |
| Area of S cingul Marginalis right hemisphere - imaging visit - value 0 | 0.0092 | 0.56 (0.14, 0.99) | 0.00029 | 35921\|21\|0 | $mm^2$ |
| Frequency of travelling from home to job workplace - mean of all visits | 0.0094 | 0.22 (0.055, 0.39) | 0.00021 | 241873\|101\|0 | times |
| Mean FA in sagittal stratum on FA skeleton left - imaging visit - value 0 | 0.0095 | -0.58 (-1, -0.14) | 0.00028 | 33550\|19\|0 | AU |
| Volume of Pole temporal left hemisphere - imaging visit - value 0 | 0.0095 | 0.56 (0.14, 0.99) | 0.00029 | 35921\|21\|0 | $mm^3$ |
| Volume of PuA right hemisphere - imaging visit - value 0 | 0.0099 | 0.56 (0.13, 0.98) | 0.00029 | 35921\|21\|0 | $mm^3$ |
| Weighted mean L1 in tract superior longitudinal fasciculus right - imaging visit - value 0 | 0.01 | 0.57 (0.14, 1) | 0.00028 | 33548\|19\|0 | AU |
| Mean FA in body of corpus callosum on FA skeleton - imaging visit - value 0 | 0.01 | -0.57 (-1, -0.14) | 0.00028 | 33550\|19\|0 | AU |
| Mean thickness of Pole occipital right hemisphere - imaging visit - value 0 | 0.01 | 0.55 (0.13, 0.98) | 0.00029 | 35921\|21\|0 | mm |
| Mean MD in sagittal stratum on FA skeleton left - imaging visit - value 0 | 0.01 | 0.57 (0.13, 1) | 0.00028 | 33550\|19\|0 | AU |
| Mean intensity of Putamen left hemisphere - imaging visit - value 0 | 0.01 | -0.55 (-0.96, -0.13) | 0.00029 | 35921\|21\|0 | AU |
| Mean FA in anterior corona radiata on FA skeleton left - imaging visit - value 0 | 0.01 | -0.57 (-1, -0.13) | 0.00028 | 33550\|19\|0 | AU |
| Mean L3 in sagittal stratum on FA skeleton left - imaging visit - value 0 | 0.01 | 0.57 (0.13, 1) | 0.00028 | 33550\|19\|0 | AU |
| Mean L2 in uncinate fasciculus on FA skeleton left - imaging visit - value 0 | 0.011 | 0.58 (0.14, 1) | 0.00028 | 33550\|19\|0 | AU |
| Volume of Basal nucleus right hemisphere - imaging visit - value 0 | 0.011 | 0.55 (0.13, 0.97) | 0.00029 | 35921\|21\|0 | $mm^3$ |
| Volume of Accessory Basal nucleus right hemisphere - imaging visit - value 0 | 0.011 | 0.55 (0.13, 0.97) | 0.00029 | 35921\|21\|0 | $mm^3$ |
| Mean FA in uncinate fasciculus on FA skeleton right - imaging visit - value 0 | 0.011 | -0.57 (-1, -0.13) | 0.00028 | 33550\|19\|0 | AU |
| Mean ICVF in uncinate fasciculus on FA skeleton left - imaging visit - value 0 | 0.011 | -0.57 (-1, -0.13) | 0.00028 | 33548\|19\|0 | AU |
| Mean L3 in fornix cres stria terminalis on FA skeleton left - imaging visit - value 0 | 0.011 | 0.58 (0.13, 1) | 0.00028 | 33550\|19\|0 | AU |
| Volume of S central left hemisphere - imaging visit - value 0 | 0.011 | -0.55 (-0.97, -0.13) | 0.00029 | 35921\|21\|0 | $mm^3$ |
| Volume of MDl left hemisphere - imaging visit - value 0 | 0.011 | 0.54 (0.12, 0.97) | 0.00029 | 35921\|21\|0 | $mm^3$ |
| Area of superiortemporal right hemisphere - imaging visit - value 0 | 0.012 | 0.54 (0.12, 0.96) | 0.00029 | 35921\|21\|0 | $mm^2$ |
| Weighted mean ICVF in tract inferior longitudinal fasciculus right - imaging visit - value 0 | 0.012 | -0.55 (-0.99, -0.12) | 0.00028 | 33547\|19\|0 | AU |

| Phenotype | P value | Beta (95% CI) | AAF | Cases (RR\|RA\|AA) | Units |
|---|---|---|---|---|---|
| Fathers age at death - mean of all visits | 0.012 | -0.21 (-0.38, -0.047) | 2.00E-04 | 323649\|132\|0 | years |
| Grey white contrast in lingual right hemisphere - imaging visit - value 0 | 0.012 | 0.53 (0.12, 0.95) | 0.00029 | 35921\|21\|0 | AU |
| Weighted mean ISOVF in tract superior longitudinal fasciculus left - imaging visit - value 0 | 0.012 | 0.56 (0.12, 1) | 0.00028 | 33547\|19\|0 | AU |
| Weighted mean L1 in tract posterior thalamic radiation right - imaging visit - value 0 | 0.012 | 0.57 (0.12, 1) | 0.00028 | 33548\|19\|0 | AU |
| Diastolic brachial blood pressure - mean of all visits | 0.013 | 0.65 (0.14, 1.2) | 0.00022 | 32350\|14\|0 | mmHg |
| Weighted mean FA in tract uncinate fasciculus left - imaging visit - value 0 | 0.013 | -0.56 (-1, -0.12) | 0.00028 | 33548\|19\|0 | AU |
| Mean signal to noise ratio SNR left - mean of all visits | 0.013 | 0.28 (0.06, 0.51) | 0.00021 | 172716\|72\|0 | no_registered_unit |
| Mean arterial pressure during PWA - mean of all visits | 0.013 | 0.65 (0.14, 1.2) | 0.00022 | 32008\|14\|0 | mmHg |
| Weighted mean ICVF in tract uncinate fasciculus right - imaging visit - value 0 | 0.013 | -0.55 (-0.98, -0.11) | 0.00028 | 33547\|19\|0 | AU |
| Total adipose tissue volume - mean of all visits | 0.013 | 0.9 (0.19, 1.6) | 0.00044 | 8030\|7\|0 | litres |
| Weighted mean L1 in tract inferior longitudinal fasciculus right - imaging visit - value 0 | 0.013 | 0.56 (0.12, 1) | 0.00028 | 33548\|19\|0 | AU |
| Diastolic brachial blood pressure during PWA - mean of all visits | 0.013 | 0.64 (0.13, 1.1) | 0.00022 | 32009\|14\|0 | mmHg |
| Mean thickness of G oc temp med Lingual left hemisphere - imaging visit - value 0 | 0.014 | 0.53 (0.11, 0.96) | 0.00029 | 35921\|21\|0 | mm |
| Maximum carotid IMT intima medial thickness at 150 degrees - mean of all visits | 0.014 | 0.65 (0.13, 1.2) | 0.00026 | 23462\|12\|0 | micrometres |
| Mean L1 in sagittal stratum on FA skeleton right - imaging visit - value 0 | 0.014 | 0.56 (0.11, 1) | 0.00028 | 33550\|19\|0 | AU |
| Mothers age at death - initial visit - mean | 0.014 | -0.23 (-0.41, -0.047) | 2.00E-04 | 254094\|102\|0 | years |
| Weighted mean L1 in tract corticospinal tract left - imaging visit - value 0 | 0.014 | 0.55 (0.11, 1) | 0.00028 | 33548\|19\|0 | AU |
| Mean L2 in middle cerebellar peduncle on FA skeleton - imaging visit - value 0 | 0.014 | 0.55 (0.11, 0.99) | 0.00028 | 33550\|19\|0 | AU |
| Weighted mean FA in tract middle cerebellar peduncle - imaging visit - value 0 | 0.014 | -0.56 (-1, -0.11) | 0.00028 | 33548\|19\|0 | AU |
| Weighted mean ICVF in tract superior longitudinal fasciculus left - imaging visit - value 0 | 0.015 | -0.54 (-0.97, -0.11) | 0.00028 | 33547\|19\|0 | AU |
| Weighted mean L2 in tract acoustic radiation right - imaging visit - value 0 | 0.015 | 0.55 (0.11, 1) | 0.00028 | 33548\|19\|0 | AU |
| Time spend outdoors in summer - mean of all visits | 0.015 | 0.18 (0.034, 0.32) | 2.00E-04 | 393360\|161\|0 | hours/day |
| Volume of Paralaminar nucleus right hemisphere - imaging visit - value 0 | 0.015 | 0.52 (0.1, 0.95) | 0.00029 | 35921\|21\|0 | $mm^3$ |
| Area of superiorfrontal left hemisphere - imaging visit - value 0 | 0.015 | 0.52 (0.1, 0.94) | 0.00029 | 35921\|21\|0 | $mm^2$ |
| Volume of grey matter in Temporal Pole right - imaging visit - value 0 | 0.015 | 0.53 (0.1, 0.97) | 0.00028 | 35591\|20\|0 | $mm^3$ |
| Volume of VA right hemisphere - imaging visit - value 0 | 0.015 | 0.52 (0.099, 0.93) | 0.00029 | 35921\|21\|0 | $mm^3$ |
| Mean ICVF in retrolenticular part of internal capsule on FA skeleton right - imaging visit - value 0 | 0.016 | -0.53 (-0.97, -0.1) | 0.00028 | 33548\|19\|0 | AU |

 

| Phenotype | P value | Beta (95% CI) | AAF | Cases (RR\|RA\|AA) | Units |
|---|---|---|---|---|---|
| Fathers age - mean of all visits | 0.016 | -0.26 (-0.46, -0.048) | 2.00E-04 | 96182\|38\|0 | years |
| Mean MD in uncinate fasciculus on FA skeleton right - imaging visit - value 0 | 0.016 | 0.54 (0.1, 0.99) | 0.00028 | 33550\|19\|0 | AU |
| Mean intensity of VentralDC left hemisphere - imaging visit - value 0 | 0.016 | -0.52 (-0.94, -0.096) | 0.00029 | 35921\|21\|0 | AU |
| Volume of superiorparietal right hemisphere - imaging visit - value 0 | 0.016 | 0.52 (0.095, 0.94) | 0.00029 | 35921\|21\|0 | $mm^3$ |
| Mean L3 in inferior cerebellar peduncle on FA skeleton right - imaging visit - value 0 | 0.016 | 0.54 (0.099, 0.99) | 0.00028 | 33550\|19\|0 | AU |
| Mean ICVF in posterior limb of internal capsule on FA skeleton left - imaging visit - value 0 | 0.016 | -0.53 (-0.97, -0.097) | 0.00028 | 33548\|19\|0 | AU |
| Heel bone mineral density BMD - mean of all visits | 0.017 | -0.21 (-0.38, -0.037) | 0.00021 | 245918\|104\|0 | $g/cm^2$ |
| Heel bone mineral density BMD - initial visit - mean | 0.017 | -0.21 (-0.38, -0.037) | 0.00021 | 245918\|104\|0 | $g/cm^2$ |
| Weighted mean MO in tract uncinate fasciculus left - imaging visit - value 0 | 0.017 | -0.54 (-0.99, -0.098) | 0.00028 | 33548\|19\|0 | AU |
| Average monthly beer plus cider intake - mean of all visits | 0.017 | 0.33 (0.06, 0.61) | 0.00029 | 40945\|24\|0 | pints |
| Mean ISOVF in genu of corpus callosum on FA skeleton - imaging visit - value 0 | 0.017 | 0.54 (0.095, 0.98) | 0.00028 | 33548\|19\|0 | AU |
| Peak expiratory flow PEF - initial visit - mean | 0.018 | -0.15 (-0.27, -0.026) | 0.00021 | 392413\|161\|0 | litres/min |
| Time spend outdoors in summer - initial visit - mean | 0.018 | 0.17 (0.03, 0.31) | 2.00E-04 | 390211\|160\|0 | hours/day |
| Age diabetes diagnosed - mean of all visits | 0.018 | 0.71 (0.12, 1.3) | 0.00017 | 20684\|7\|0 | years |
| Weighted mean L3 in tract forceps minor - imaging visit - value 0 | 0.018 | 0.53 (0.089, 0.96) | 0.00028 | 33548\|19\|0 | AU |
| Mean L1 in anterior limb of internal capsule on FA skeleton left - imaging visit - value 0 | 0.019 | 0.53 (0.088, 0.97) | 0.00028 | 33550\|19\|0 | AU |
| Weighted mean L2 in tract medial lemniscus left - imaging visit - value 0 | 0.019 | 0.53 (0.088, 0.98) | 0.00028 | 33548\|19\|0 | AU |
| Mean FA in tapetum on FA skeleton left - imaging visit - value 0 | 0.019 | -0.53 (-0.97, -0.086) | 0.00028 | 33550\|19\|0 | AU |
| Volume of Corticoamygdaloid transitio right hemisphere - imaging visit - value 0 | 0.019 | 0.51 (0.082, 0.93) | 0.00029 | 35921\|21\|0 | $mm^3$ |
| Mean ISOVF in fornix cres stria terminalis on FA skeleton left - imaging visit - value 0 | 0.019 | 0.53 (0.087, 0.98) | 0.00028 | 33548\|19\|0 | AU |
| Volume of LP left hemisphere - imaging visit - value 0 | 0.019 | 0.5 (0.08, 0.92) | 0.00029 | 35921\|21\|0 | $mm^3$ |
| Grey white contrast in cuneus left hemisphere - imaging visit - value 0 | 0.02 | 0.5 (0.08, 0.91) | 0.00029 | 35921\|21\|0 | AU |
| Area of superiortemporal right hemisphere - imaging visit - value 0 | 0.02 | 0.5 (0.081, 0.92) | 0.00029 | 35921\|21\|0 | $mm^3$ |
| Heel bone mineral density BMD T score automated - mean of all visits | 0.02 | -0.2 (-0.37, -0.032) | 0.00021 | 246031\|104\|0 | Std.Devs |
| Heel bone mineral density BMD T score automated - initial visit - mean | 0.02 | -0.2 (-0.37, -0.032) | 0.00021 | 246031\|104\|0 | Std.Devs |
| Heel quantitative ultrasound index QUI direct entry - mean of all visits | 0.02 | -0.2 (-0.37, -0.032) | 0.00021 | 246031\|104\|0 | no_registered_unit |
| Heel quantitative ultrasound index QUI direct entry - initial visit - mean | 0.02 | -0.2 (-0.37, -0.032) | 0.00021 | 246031\|104\|0 | no_registered_unit |

| Phenotype | P value | Beta (95% CI) | AAF | Cases (RR\|RA\|AA) | Units |
|---|---|---|---|---|---|
| Mean intensity of Pallidum right hemisphere - imaging visit - value 0 | 0.02 | -0.5 (-0.92, -0.079) | 0.00029 | 35921\|21\|0 | AU |
| Mean ISOVF in posterior thalamic radiation on FA skeleton left - imaging visit - value 0 | 0.02 | 0.53 (0.084, 0.97) | 0.00028 | 33548\|19\|0 | AU |
| Volume of L Sg left hemisphere - imaging visit - value 0 | 0.02 | 0.5 (0.078, 0.92) | 0.00029 | 35921\|21\|0 | $mm^3$ |
| Speed of sound through heel - mean of all visits | 0.02 | -0.2 (-0.37, -0.032) | 0.00021 | 246031\|104\|0 | m/s |
| Speed of sound through heel - initial visit - mean | 0.02 | -0.2 (-0.37, -0.032) | 0.00021 | 246031\|104\|0 | m/s |
| Area of insula left hemisphere - imaging visit - value 0 | 0.021 | 0.5 (0.077, 0.92) | 0.00029 | 35921\|21\|0 | $mm^2$ |
| Mean L2 in uncinate fasciculus on FA skeleton right - imaging visit - value 0 | 0.021 | 0.52 (0.08, 0.96) | 0.00028 | 33550\|19\|0 | AU |
| Age at first live birth - mean of all visits | 0.021 | -0.28 (-0.52, -0.043) | 0.00019 | 158561\|59\|0 | years |
| Mean thickness of S central left hemisphere - imaging visit - value 0 | 0.021 | -0.5 (-0.92, -0.076) | 0.00029 | 35921\|21\|0 | mm |
| Weighted mean MD in tract forceps minor - imaging visit - value 0 | 0.021 | 0.52 (0.078, 0.96) | 0.00028 | 33548\|19\|0 | AU |
| Mean L2 in medial lemniscus on FA skeleton left - imaging visit - value 0 | 0.021 | 0.52 (0.079, 0.97) | 0.00028 | 33550\|19\|0 | AU |
| Weighted mean L3 in tract acoustic radiation right - imaging visit - value 0 | 0.021 | 0.52 (0.079, 0.97) | 0.00028 | 33548\|19\|0 | AU |
| Creatinine - mean of all visits | 0.021 | 0.14 (0.02, 0.25) | 2.00E-04 | 411405\|164\|0 | mg/dL |
| Creatinine - initial visit - mean | 0.021 | 0.14 (0.02, 0.25) | 2.00E-04 | 410621\|164\|0 | mg/dL |
| Age at first live birth - initial visit - mean | 0.022 | -0.28 (-0.52, -0.041) | 0.00019 | 158529\|59\|0 | years |
| Mean OD in superior fronto occipital fasciculus on FA skeleton left - imaging visit - value 0 | 0.022 | -0.52 (-0.96, -0.075) | 0.00028 | 33548\|19\|0 | AU |
| Speech reception threshold SRT estimate right - mean of all visits | 0.022 | 0.26 (0.038, 0.48) | 0.00021 | 170294\|72\|0 | no_registered_unit |
| Mean thickness of S circular insula sup left hemisphere - imaging visit - value 0 | 0.022 | -0.49 (-0.92, -0.071) | 0.00029 | 35921\|21\|0 | mm |
| Weighted mean L1 in tract superior longitudinal fasciculus left - imaging visit - value 0 | 0.022 | 0.51 (0.073, 0.94) | 0.00028 | 33548\|19\|0 | AU |
| Fathers age - initial visit - mean | 0.023 | -0.24 (-0.44, -0.033) | 2.00E-04 | 96087\|38\|0 | years |
| Average acceleration 06 00 06 59 - mean of all visits | 0.023 | 0.37 (0.051, 0.7) | 0.00019 | 91659\|34\|0 | milli-gravity |
| Average acceleration 06 00 06 59 - initial visit - mean | 0.023 | 0.37 (0.051, 0.7) | 0.00019 | 91659\|34\|0 | milli-gravity |
| Grey white contrast in insula left hemisphere - imaging visit - value 0 | 0.023 | -0.48 (-0.9, -0.066) | 0.00029 | 35921\|21\|0 | AU |
| Weighted mean L2 in tract forceps minor - imaging visit - value 0 | 0.023 | 0.51 (0.069, 0.95) | 0.00028 | 33548\|19\|0 | AU |
| Baby birth weight - mean of all visits | 0.023 | 0.85 (0.12, 1.6) | 0.00018 | 13632\|5\|0 | g |
| Baby birth weight - initial visit - mean | 0.023 | 0.85 (0.12, 1.6) | 0.00018 | 13632\|5\|0 | g |
| Volume of LP right hemisphere - imaging visit - value 0 | 0.023 | 0.48 (0.066, 0.9) | 0.00029 | 35921\|21\|0 | $mm^3$ |

| Phenotype | P value | Beta (95% CI) | AAF | Cases (RR\|RA\|AA) | Units |
|---|---|---|---|---|---|
| Standing height - initial visit - mean | 0.023 | -0.1 (-0.19, -0.014) | 2.00E-04 | 429531\|172\|0 | cm |
| Standing height - mean of all visits | 0.024 | -0.099 (-0.19, -0.013) | 2.00E-04 | 429571\|172\|0 | cm |
| Area of superiortemporal right hemisphere - imaging visit - value 0 | 0.024 | 0.48 (0.064, 0.9) | 0.00029 | 35921\|21\|0 | $mm^2$ |
| Mean intensity of VentralDC right hemisphere - imaging visit - value 0 | 0.024 | -0.49 (-0.91, -0.063) | 0.00029 | 35921\|21\|0 | AU |
| Mean intensity of Putamen right hemisphere - imaging visit - value 0 | 0.025 | -0.48 (-0.9, -0.061) | 0.00029 | 35921\|21\|0 | AU |
| Area of posteriorcingulate right hemisphere - imaging visit - value 0 | 0.025 | 0.49 (0.062, 0.91) | 0.00029 | 35921\|21\|0 | $mm^2$ |
| Mean ISOVF in superior longitudinal fasciculus on FA skeleton right - imaging visit - value 0 | 0.025 | 0.5 (0.064, 0.95) | 0.00028 | 33548\|19\|0 | AU |
| Mean thickness of V1 right hemisphere - imaging visit - value 0 | 0.025 | 0.48 (0.061, 0.91) | 0.00029 | 35921\|21\|0 | mm |
| Mean ICVF in sagittal stratum on FA skeleton left - imaging visit - value 0 | 0.025 | -0.49 (-0.93, -0.062) | 0.00028 | 33548\|19\|0 | AU |
| Mean FA in inferior cerebellar peduncle on FA skeleton right - imaging visit - value 0 | 0.025 | -0.51 (-0.95, -0.063) | 0.00028 | 33550\|19\|0 | AU |
| 6mm asymmetry index left - initial visit - mean | 0.026 | 0.48 (0.059, 0.9) | 0.00021 | 48878\|21\|0 | no_registered_unit |
| Age high blood pressure diagnosed - initial visit - mean | 0.026 | 0.23 (0.028, 0.44) | 0.00028 | 103405\|58\|0 | years |
| Mean L2 in posterior limb of internal capsule on FA skeleton left - imaging visit - value 0 | 0.026 | 0.5 (0.06, 0.94) | 0.00028 | 33550\|19\|0 | AU |
| Area of paracentral right hemisphere - imaging visit - value 0 | 0.026 | 0.48 (0.057, 0.9) | 0.00029 | 35921\|21\|0 | $mm^2$ |
| Weighted mean ICVF in tract uncinate fasciculus left - imaging visit - value 0 | 0.026 | -0.49 (-0.93, -0.059) | 0.00028 | 33547\|19\|0 | AU |
| Grey white contrast in lateraloccipital right hemisphere - imaging visit - value 0 | 0.026 | 0.47 (0.056, 0.89) | 0.00029 | 35921\|21\|0 | AU |
| Mean L3 in middle cerebellar peduncle on FA skeleton - imaging visit - value 0 | 0.026 | 0.5 (0.059, 0.94) | 0.00028 | 33550\|19\|0 | AU |
| Volume of grey matter in Pallidum right - imaging visit - value 0 | 0.027 | 0.49 (0.057, 0.92) | 0.00028 | 35591\|20\|0 | $mm^3$ |
| Mean thickness of S temporal sup left hemisphere - imaging visit - value 0 | 0.027 | -0.48 (-0.9, -0.055) | 0.00029 | 35921\|21\|0 | mm |
| Mean OD in inferior cerebellar peduncle on FA skeleton left - imaging visit - value 0 | 0.027 | 0.5 (0.057, 0.95) | 0.00028 | 33548\|19\|0 | AU |
| Bread intake - mean of all visits | 0.028 | 0.16 (0.018, 0.3) | 0.00019 | 421644\|164\|0 | slices/week |
| Signal to noise ratio SNR of triplet right - mean of all visits | 0.028 | 0.25 (0.027, 0.47) | 0.00021 | 173060\|73\|0 | no_registered_unit |
| Mean FA in corticospinal tract on FA skeleton right - imaging visit - value 0 | 0.028 | -0.5 (-0.94, -0.055) | 0.00028 | 33550\|19\|0 | AU |
| Mean MD in uncinate fasciculus on FA skeleton left - imaging visit - value 0 | 0.028 | 0.5 (0.055, 0.94) | 0.00028 | 33550\|19\|0 | AU |
| OD Contour 2 to 3 Region I3 Mean | 0.028 | -0.6 (-1.1, -0.065) | 0.00015 | 43616\|13\|0 | NA |
| Mean ISOVF in superior longitudinal fasciculus on FA skeleton left - imaging visit - value 0 | 0.028 | 0.49 (0.054, 0.94) | 0.00028 | 33548\|19\|0 | AU |
| Number of measurements made - initial visit - mean | 0.028 | 0.11 (0.012, 0.21) | 0.00021 | 392413\|161\|0 | no_registered_unit |

| Phenotype | P value | Beta (95% CI) | AAF | Cases (RR\|RA\|AA) | Units |
|---|---|---|---|---|---|
| Mean L1 in superior longitudinal fasciculus on FA skeleton left - imaging visit - value 0 | 0.028 | 0.49 (0.052, 0.92) | 0.00028 | 33550\|19\|0 | AU |
| Volume of CeM left hemisphere - imaging visit - value 0 | 0.028 | 0.47 (0.049, 0.89) | 0.00029 | 35921\|21\|0 | $mm^3$ |
| Area of medialorbitofrontal left hemisphere - imaging visit - value 0 | 0.028 | 0.47 (0.05, 0.9) | 0.00029 | 35921\|21\|0 | $mm^2$ |
| Weighted mean L2 in tract forceps major - imaging visit - value 0 | 0.029 | 0.49 (0.05, 0.93) | 0.00028 | 33548\|19\|0 | AU |
| FEV1 FVC ratio Z score - mean of all visits | 0.029 | -0.18 (-0.34, -0.018) | 2.00E-04 | 342887\|136\|0 | Z |
| FEV1 FVC ratio Z score - initial visit - mean | 0.029 | -0.18 (-0.34, -0.018) | 2.00E-04 | 342887\|136\|0 | Z |
| Peak expiratory flow PEF - mean of all visits | 0.03 | -0.13 (-0.26, -0.013) | 2.00E-04 | 394984\|162\|0 | litres/min |
| Mean FA in posterior limb of internal capsule on FA skeleton left - imaging visit - value 0 | 0.03 | -0.49 (-0.92, -0.048) | 0.00028 | 33550\|19\|0 | AU |
| Mean L3 in cerebral peduncle on FA skeleton left - imaging visit - value 0 | 0.03 | 0.49 (0.048, 0.94) | 0.00028 | 33550\|19\|0 | AU |
| Area of G S paracentral right hemisphere - imaging visit - value 0 | 0.03 | 0.47 (0.045, 0.89) | 0.00029 | 35921\|21\|0 | $mm^2$ |
| Mean tfMRI head motion averaged across space and time points - mean of all visits | 0.03 | 0.68 (0.065, 1.3) | 0.00026 | 17058\|9\|0 | mm |
| Weighted mean MD in tract uncinate fasciculus right - imaging visit - value 0 | 0.031 | 0.48 (0.044, 0.92) | 0.00028 | 33548\|19\|0 | AU |
| Time first key touched - initial visit - mean | 0.031 | -0.58 (-1.1, -0.053) | 0.00013 | 44675\|12\|0 | milliseconds |
| Average acceleration 07 00 07 59 - mean of all visits | 0.031 | 0.35 (0.032, 0.68) | 0.00019 | 91659\|34\|0 | milli-gravity |
| Average acceleration 07 00 07 59 - initial visit - mean | 0.031 | 0.35 (0.032, 0.68) | 0.00019 | 91659\|34\|0 | milli-gravity |
| Weighted mean L3 in tract forceps major - imaging visit - value 0 | 0.031 | 0.49 (0.044, 0.93) | 0.00028 | 33548\|19\|0 | AU |
| Area of paracentral right hemisphere - imaging visit - value 0 | 0.031 | 0.47 (0.042, 0.89) | 0.00029 | 35921\|21\|0 | $mm^2$ |
| Mean L1 in retrolenticular part of internal capsule on FA skeleton right - imaging visit - value 0 | 0.031 | 0.49 (0.044, 0.93) | 0.00028 | 33550\|19\|0 | AU |
| Mean L3 in uncinate fasciculus on FA skeleton left - imaging visit - value 0 | 0.031 | 0.49 (0.043, 0.93) | 0.00028 | 33550\|19\|0 | AU |
| delta1 deconf | 0.032 | 0.44 (0.037, 0.84) | 0.00026 | 28316\|15\|0 | no_registered_unit |
| Diastolic blood pressure automated reading - mean of all visits | 0.032 | 0.16 (0.014, 0.31) | 2.00E-04 | 407743\|160\|0 | mmHg |
| Time spent watching television TV - mean of all visits | 0.032 | 0.15 (0.013, 0.29) | 2.00E-04 | 409778\|166\|0 | hours/day |
| Mean thickness of Lat Fis ant Horizont right hemisphere - imaging visit - value 0 | 0.033 | -0.46 (-0.89, -0.039) | 0.00029 | 35921\|21\|0 | mm |
| Volume of PuL left hemisphere - imaging visit - value 0 | 0.033 | 0.46 (0.038, 0.88) | 0.00029 | 35921\|21\|0 | $mm^3$ |
| Area of entorhinal left hemisphere - imaging visit - value 0 | 0.033 | 0.46 (0.037, 0.88) | 0.00029 | 35921\|21\|0 | $mm^2$ |
| Weighted mean ISOVF in tract posterior thalamic radiation left - imaging visit - value 0 | 0.033 | 0.48 (0.039, 0.93) | 0.00028 | 33547\|19\|0 | AU |
| Weighted mean L3 in tract uncinate fasciculus right - imaging visit - value 0 | 0.034 | 0.48 (0.037, 0.91) | 0.00028 | 33548\|19\|0 | AU |

| Phenotype | P value | Beta (95% CI) | AAF | Cases (RR\|RA\|AA) | Units |
|---|---|---|---|---|---|
| Heel Broadband ultrasound attenuation direct entry - mean of all visits | 0.034 | -0.18 (-0.35, -0.014) | 0.00021 | 246016\|104\|0 | dB/MHz |
| Heel Broadband ultrasound attenuation direct entry - initial visit - mean | 0.034 | -0.18 (-0.35, -0.014) | 0.00021 | 246016\|104\|0 | dB/MHz |
| Mean MO in cingulum hippocampus on FA skeleton left - imaging visit - value 0 | 0.034 | -0.48 (-0.93, -0.036) | 0.00028 | 33550\|19\|0 | AU |
| Scanner transverse Y brain position - mean of all visits | 0.034 | 0.61 (0.045, 1.2) | 0.00024 | 20407\|10\|0 | Units |
| Neutrophill percentage - initial visit - mean | 0.035 | 0.15 (0.011, 0.3) | 2.00E-04 | 417306\|167\|0 | percent |
| Volume of Amygdala right hemisphere - imaging visit - value 0 | 0.035 | 0.45 (0.032, 0.88) | 0.00029 | 35921\|21\|0 | $mm^3$ |
| Mean intensity of CC Mid Posterior whole brain - imaging visit - value 0 | 0.036 | -0.45 (-0.88, -0.03) | 0.00029 | 35921\|21\|0 | AU |
| Weighted mean ICVF in tract forceps major - imaging visit - value 0 | 0.036 | -0.47 (-0.9, -0.031) | 0.00028 | 33547\|19\|0 | AU |
| Volume of S collat transv post left hemisphere - imaging visit - value 0 | 0.037 | 0.45 (0.027, 0.88) | 0.00029 | 35921\|21\|0 | $mm^3$ |
| Year job started - mean of all visits | 0.037 | -0.21 (-0.4, -0.012) | 0.00021 | 107327\|45\|0 | calendar year |
| Year job started - initial visit - mean | 0.037 | -0.21 (-0.4, -0.012) | 0.00021 | 107327\|45\|0 | calendar year |
| Mean signal to noise ratio SNR right - mean of all visits | 0.037 | 0.24 (0.014, 0.46) | 0.00021 | 172703\|73\|0 | no_registered_unit |
| Volume of G parietal sup right hemisphere - imaging visit - value 0 | 0.037 | 0.45 (0.026, 0.88) | 0.00029 | 35921\|21\|0 | $mm^3$ |
| Volume of S oc sup transversal left hemisphere - imaging visit - value 0 | 0.037 | 0.45 (0.026, 0.88) | 0.00029 | 35921\|21\|0 | $mm^3$ |
| Duration visual acuity screen displayed right - initial visit - mean | 0.038 | 0.31 (0.018, 0.61) | 0.00022 | 96537\|42\|0 | seconds |
| Volume of PuL right hemisphere - imaging visit - value 0 | 0.038 | 0.45 (0.025, 0.87) | 0.00029 | 35921\|21\|0 | $mm^3$ |
| Volume of superiorparietal right hemisphere - imaging visit - value 0 | 0.038 | 0.45 (0.025, 0.88) | 0.00029 | 35921\|21\|0 | $mm^3$ |
| Ankle spacing width right - mean of all visits | 0.038 | 0.2 (0.011, 0.4) | 0.00021 | 166147\|69\|0 | mm |
| Grey white contrast in lateraloccipital left hemisphere - imaging visit - value 0 | 0.038 | 0.44 (0.024, 0.85) | 0.00029 | 35921\|21\|0 | AU |
| Time spent watching television TV - initial visit - mean | 0.038 | 0.15 (0.0081, 0.29) | 2.00E-04 | 407184\|165\|0 | hours/day |
| Area of precuneus right hemisphere - imaging visit - value 0 | 0.038 | 0.44 (0.024, 0.86) | 0.00029 | 35921\|21\|0 | $mm^2$ |
| Mean L1 in tapetum on FA skeleton right - imaging visit - value 0 | 0.038 | 0.47 (0.026, 0.92) | 0.00028 | 33550\|19\|0 | AU |
| Weighted mean FA in tract forceps minor - imaging visit - value 0 | 0.038 | -0.46 (-0.9, -0.025) | 0.00028 | 33548\|19\|0 | AU |
| Volume of S temporal transverse right hemisphere - imaging visit - value 0 | 0.038 | 0.45 (0.024, 0.88) | 0.00029 | 35920\|21\|0 | $mm^3$ |
| Weighted mean L2 in tract middle cerebellar peduncle - imaging visit - value 0 | 0.038 | 0.47 (0.025, 0.92) | 0.00028 | 33548\|19\|0 | AU |
| Volume of S cingul Marginalis right hemisphere - imaging visit - value 0 | 0.039 | 0.45 (0.024, 0.88) | 0.00029 | 35921\|21\|0 | $mm^3$ |
| Volume of grey matter in Supracalcarine Cortex left - imaging visit - value 0 | 0.039 | 0.45 (0.024, 0.89) | 0.00028 | 35591\|20\|0 | $mm^3$ |

| Phenotype | P value | Beta (95% CI) | AAF | Cases (RR\|RA\|AA) | Units |
|---|---|---|---|---|---|
| Mean L1 in retrolenticular part of internal capsule on FA skeleton left - imaging visit - value 0 | 0.039 | 0.47 (0.024, 0.91) | 0.00028 | 33550\|19\|0 | AU |
| Duration to entering value - mean of all visits | 0.039 | 0.28 (0.014, 0.55) | 0.00018 | 105503\|39\|0 | milliseconds |
| Duration to entering value - initial visit - mean | 0.039 | 0.28 (0.014, 0.55) | 0.00018 | 105503\|39\|0 | milliseconds |
| Volume of Hippocampal tail left hemisphere - imaging visit - value 0 | 0.039 | 0.44 (0.021, 0.86) | 0.00029 | 35921\|21\|0 | $mm^3$ |
| Area of precuneus right hemisphere - imaging visit - value 0 | 0.04 | 0.44 (0.021, 0.86) | 0.00029 | 35921\|21\|0 | $mm^2$ |
| Grey white contrast in cuneus right hemisphere - imaging visit - value 0 | 0.04 | 0.44 (0.02, 0.85) | 0.00029 | 35921\|21\|0 | AU |
| OD Contour 1 to 2 Region I3 Mean | 0.04 | 0.55 (0.025, 1.1) | 0.00015 | 43616\|13\|0 | NA |
| Ankle spacing width right - initial visit - mean | 0.04 | 0.23 (0.01, 0.45) | 0.00019 | 137273\|53\|0 | mm |
| Area of precuneus right hemisphere - imaging visit - value 0 | 0.04 | 0.44 (0.019, 0.86) | 0.00029 | 35921\|21\|0 | $mm^2$ |
| Mean OD in inferior cerebellar peduncle on FA skeleton right - imaging visit - value 0 | 0.04 | 0.46 (0.02, 0.91) | 0.00028 | 33548\|19\|0 | AU |
| Mean thickness of S suborbital right hemisphere - imaging visit - value 0 | 0.041 | -0.45 (-0.87, -0.019) | 0.00029 | 35921\|21\|0 | mm |
| Systolic brachial blood pressure - mean of all visits | 0.041 | 0.52 (0.022, 1) | 0.00022 | 32348\|14\|0 | mmHg |
| Mean L3 in medial lemniscus on FA skeleton left - imaging visit - value 0 | 0.041 | 0.46 (0.019, 0.91) | 0.00028 | 33550\|19\|0 | AU |
| Area of entorhinal left hemisphere - imaging visit - value 0 | 0.041 | 0.44 (0.018, 0.86) | 0.00029 | 35921\|21\|0 | $mm^2$ |
| Area of S oc sup transversal left hemisphere - imaging visit - value 0 | 0.041 | 0.44 (0.017, 0.87) | 0.00029 | 35921\|21\|0 | $mm^2$ |
| Mean thickness of G S frontomargin right hemisphere - imaging visit - value 0 | 0.041 | -0.44 (-0.87, -0.017) | 0.00029 | 35921\|21\|0 | mm |
| Basophill count - mean of all visits | 0.041 | 0.14 (0.0055, 0.28) | 2.00E-04 | 417965\|167\|0 | $10^9$ cells/Litre |
| Systolic brachial blood pressure during PWA - mean of all visits | 0.042 | 0.52 (0.02, 1) | 0.00022 | 32009\|14\|0 | mmHg |
| Central systolic blood pressure during PWA - mean of all visits | 0.042 | 0.51 (0.019, 1) | 0.00022 | 32008\|14\|0 | mmHg |
| 6mm asymmetry index left - mean of all visits | 0.042 | 0.42 (0.015, 0.82) | 0.00021 | 55432\|23\|0 | no_registered_unit |
| Immature reticulocyte fraction - initial visit - mean | 0.042 | 0.15 (0.0054, 0.3) | 2.00E-04 | 411137\|164\|0 | ratio |
| Mean thickness of G oc temp med Lingual right hemisphere - imaging visit - value 0 | 0.042 | 0.44 (0.015, 0.86) | 0.00029 | 35921\|21\|0 | mm |
| Average monthly beer plus cider intake - initial visit - mean | 0.043 | 0.32 (0.01, 0.62) | 3.00E-04 | 32090\|19\|0 | pints |
| Volume of precuneus right hemisphere - imaging visit - value 0 | 0.043 | 0.43 (0.013, 0.85) | 0.00029 | 35921\|21\|0 | $mm^3$ |
| Bread intake - initial visit - mean | 0.043 | 0.15 (0.0045, 0.29) | 0.00019 | 420880\|164\|0 | slices/week |
| Mean thickness of G pariet inf Supramar left hemisphere - imaging visit - value 0 | 0.043 | -0.44 (-0.86, -0.013) | 0.00029 | 35921\|21\|0 | mm |
| Time first key touched - mean of all visits | 0.043 | -0.39 (-0.78, -0.012) | 0.00017 | 67222\|23\|0 | milliseconds |

| Phenotype | P value | Beta (95% CI) | AAF | Cases (RR\|RA\|AA) | Units |
|---|---|---|---|---|---|
| Volume of L Sg right hemisphere - imaging visit - value 0 | 0.044 | 0.44 (0.012, 0.86) | 0.00029 | 35921\|21\|0 | $mm^3$ |
| QRS duration - mean of all visits | 0.044 | -0.44 (-0.87, -0.012) | 0.00025 | 33775\|17\|0 | ms |
| Weighted mean MO in tract medial lemniscus left - imaging visit - value 0 | 0.045 | -0.45 (-0.9, -0.011) | 0.00028 | 33548\|19\|0 | AU |
| Mean FA in medial lemniscus on FA skeleton right - imaging visit - value 0 | 0.045 | -0.45 (-0.9, -0.011) | 0.00028 | 33550\|19\|0 | AU |
| RGC_CornealAstigmatism_3mm_right | 0.045 | 0.3 (0.0066, 0.59) | 0.00021 | 103404\|44\|0 | NA |
| 3mm cylindrical power right - mean of all visits | 0.045 | -0.3 (-0.59, -0.0066) | 0.00021 | 103404\|44\|0 | diopters |
| Years since last cervical smear test - mean of all visits | 0.045 | 0.21 (0.0047, 0.43) | 0.00019 | 182585\|70\|0 | years |
| Grey white contrast in temporalpole left hemisphere - imaging visit - value 0 | 0.045 | -0.43 (-0.85, -0.0093) | 0.00029 | 35921\|21\|0 | AU |
| Mean thickness of G cuneus right hemisphere - imaging visit - value 0 | 0.045 | 0.43 (0.0093, 0.86) | 0.00029 | 35921\|21\|0 | mm |
| Average weekly beer plus cider intake - initial visit - mean | 0.045 | 0.13 (0.0028, 0.26) | 0.00017 | 304976\|104\|0 | pints |
| Volume of S parieto occipital left hemisphere - imaging visit - value 0 | 0.046 | 0.43 (0.0083, 0.85) | 0.00029 | 35921\|21\|0 | $mm^3$ |
| Diastolic blood pressure automated reading - initial visit - mean | 0.046 | 0.15 (0.0028, 0.3) | 0.00019 | 405855\|158\|0 | mmHg |
| Retinol - initial visit - mean | 0.046 | 0.4 (0.0074, 0.79) | 2.00E-04 | 59316\|24\|0 | ug |
| Area of G Ins lg S cent ins left hemisphere - imaging visit - value 0 | 0.046 | 0.43 (0.0077, 0.86) | 0.00029 | 35921\|21\|0 | $mm^2$ |
| Weighted mean MO in tract parahippocampal part of cingulum left - imaging visit - value 0 | 0.046 | -0.46 (-0.9, -0.0079) | 0.00028 | 33548\|19\|0 | AU |
| Inverted temporal signal to noise ratio in pre processed tfMRI - mean of all visits | 0.046 | 0.63 (0.0099, 1.2) | 0.00026 | 17058\|9\|0 | ratio |
| Mean OD in posterior corona radiata on FA skeleton left - imaging visit - value 0 | 0.047 | -0.45 (-0.89, -0.0068) | 0.00028 | 33548\|19\|0 | AU |
| Volume of Hippocampal tail right hemisphere - imaging visit - value 0 | 0.047 | 0.42 (0.0056, 0.84) | 0.00029 | 35921\|21\|0 | $mm^3$ |
| Weighted mean ISOVF in tract uncinate fasciculus left - imaging visit - value 0 | 0.047 | 0.45 (0.0054, 0.9) | 0.00028 | 33547\|19\|0 | AU |
| Mean OD in superior cerebellar peduncle on FA skeleton left - imaging visit - value 0 | 0.047 | 0.45 (0.0051, 0.89) | 0.00028 | 33548\|19\|0 | AU |
| Weighted mean ISOVF in tract anterior thalamic radiation left - imaging visit - value 0 | 0.047 | 0.45 (0.0052, 0.89) | 0.00028 | 33547\|19\|0 | AU |
| Mean L1 in fornix cres stria terminalis on FA skeleton left - imaging visit - value 0 | 0.048 | 0.45 (0.0036, 0.9) | 0.00028 | 33550\|19\|0 | AU |
| Weighted mean OD in tract middle cerebellar peduncle - imaging visit - value 0 | 0.048 | 0.45 (0.0032, 0.9) | 0.00028 | 33547\|19\|0 | AU |
| Mean MO in sagittal stratum on FA skeleton right - imaging visit - value 0 | 0.049 | -0.45 (-0.89, -0.0024) | 0.00028 | 33550\|19\|0 | AU |
| Mothers age at death - mean of all visits | 0.049 | -0.18 (-0.36, -0.00085) | 0.00021 | 263112\|108\|0 | years |
| Neutrophill percentage - mean of all visits | 0.049 | 0.14 (0.00051, 0.29) | 2.00E-04 | 417970\|167\|0 | percent |

| Phenotype | P value | Beta (95% CI) | AAF | Cases (RR\|RA\|AA) | Units |
|---|---|---|---|---|---|
| Volume of grey matter in Precuneous Cortex right - imaging visit - value 0 | 0.049 | 0.43 (0.0014, 0.86) | 0.00028 | 35591\|20\|0 | $mm^3$ |
| Volume of paracentral right hemisphere - imaging visit - value 0 | 0.049 | 0.43 (0.0014, 0.85) | 0.00029 | 35921\|21\|0 | $mm^3$ |

1. Shown are quantitative phenotypes with p values below 0.05, sorted by increasing p-value. From left-to-right is the phenotype, the effect size (regression beta on RINT scale [11-13] with 95% confidence interval), the p-value, counts of ref/ref genotypes, counts of ref/alt heterozygous genotypes, and units. AU, arbitrary units.
2. Highlight of phenotypes mentioned in the main text

**Supplementary Table 21**. ***NOTCH3* EGFr Domains[1]**.

| Domain | Start | End |
|---|---|---|
| EGFr 1 | 40 | 77 |
| EGFr 2 | 78 | 118 |
| EGFr 3 | 119 | 156 |
| EGFr 4 | 158 | 195 |
| EGFr 5 | 197 | 234 |
| EGFr 6 | 236 | 272 |
| EGFr 7 | 274 | 312 |
| EGFr 8 | 314 | 350 |
| EGFr 9 | 351 | 389 |
| EGFr 10 | 391 | 429 |
| EGFr 11 | 431 | 467 |
| EGFr 12 | 469 | 505 |
| EGFr 13 | 507 | 543 |
| EGFr 14 | 545 | 580 |
| EGFr 15 | 582 | 618 |
| EGFr 16 | 620 | 655 |
| EGFr 17 | 657 | 693 |
| EGFr 18 | 695 | 730 |
| EGFr 19 | 734 | 770 |
| EGFr 20 | 771 | 808 |
| EGFr 21 | 810 | 847 |
| EGFr 22 | 849 | 885 |
| EGFr 23 | 887 | 922 |
| EGFr 24 | 924 | 960 |
| EGFr 25 | 962 | 998 |
| EGFr 26 | 1000 | 1034 |
| EGFr 27 | 1036 | 1082 |
| EGFr 28 | 1084 | 1120 |
| EGFr 29 | 1122 | 1158 |
| EGFr 30 | 1160 | 1203 |
| EGFr 31 | 1205 | 1244 |
| EGFr 32 | 1246 | 1287 |
| EGFr 33 | 1289 | 1325 |
| EGFr 34 | 1335 | 1373 |

1. The start and end positions in the protein sequence for each of 34 EGFr domains was obtained from PROTSITE [14]. Shown is the domain #, codon start residue, and codon end residue.

**Supplementary Table 22. Cys-altering Variants in UKB Included in Burden Test[1].**

| Chrom:pos:ref:alt | Exon | Domain class | EGFr Domain number | HGVS transcript | HGVS protein | Odds Ratio (95 % CI) | Pval | AAF | AAC | Case Counts | Control Counts | UKB AAF | PGR AAF |
|---|---|---|---|---|---|---|---|---|---|---|---|---|---|
| 19:15197537:G:A | Exon 2 | EGFr 1-6 | EGFr 1 | c.160C>T | p.Arg54Cys | NC | NC | 1.16E-06 | 1 | 9143\|0\|0 | 371402\|1\|0 | 1.10E-06 | 0.00E+00 |
| 19:15192449:G:A | Exon 3 | EGFr 1-6 | EGFr 2 | c.268C>T | p.Arg90Cys | NC | NC | 1.16E-06 | 1 | 9142\|1\|0 | 371403\|0\|0 | 1.10E-06 | 0.00E+00 |
| 19:15192389:G:A | Exon 3 | EGFr 1-6 | EGFr 2 | c.328C>T | p.Arg110Cys | NC | NC | 2.32E-06 | 2 | 9142\|1\|0 | 371399\|1\|0 | 2.20E-06 | 0.00E+00 |
| 19:15192218:G:A | Exon 4 | EGFr 1-6 | EGFr 3 | c.421C>T | p.Arg141Cys | NC | NC | 5.80E-06 | 4 | 9141\|2\|0 | 371399\|2\|0 | 5.50E-06 | 0.00E+00 |
| 19:15192182:G:A | Exon 4 | EGFr 1-6 | EGFr 3 | c.457C>T | p.Arg153Cys | NC | NC | 1.16E-06 | 1 | 9142\|1\|0 | 371396\|0\|0 | 1.10E-06 | 0.00E+00 |
| 19:15192134:G:A | Exon 4 | EGFr 1-6 | EGFr 4 | c.505C>T | p.Arg169Cys | NC | NC | 2.32E-06 | 2 | 9143\|0\|0 | 371401\|2\|0 | 2.20E-06 | 6.26E-06 |
| 19:15192095:G:A | Exon 4 | EGFr 1-6 | EGFr 4 | c.544C>T | p.Arg182Cys | NC | NC | 8.12E-06 | 3 | 9142\|1\|0 | 371401\|2\|0 | 7.70E-06 | 0.00E+00 |
| 19:15192020:G:A | Exon 4 | EGFr 1-6 | EGFr 5 | c.619C>T | p.Arg207Cys | NC | NC | 2.32E-06 | 1 | 9143\|0\|0 | 371402\|1\|0 | 2.20E-06 | 1.25E-05 |
| 19:15191529:A:T | Exon 6 | EGFr 7-34 | EGFr 7 | c.931T>A | p.Cys311Ser | NC | NC | 2.32E-06 | 1 | 9143\|0\|0 | 371402\|1\|0 | 2.20E-06 | 0.00E+00 |
| 19:15191507:C:A | Exon 6 | EGFr 7-34 | EGFr 8 | c.953G>T | p.Cys318Phe | NC | NC | 2.32E-06 | 1 | 9143\|0\|0 | 371402\|1\|0 | 2.20E-06 | 0.00E+00 |
| 19:15191466:G:A | Exon 6 | EGFr 7-34 | EGFr 8 | c.994C>T | p.Arg332Cys | NC | NC | 1.16E-06 | 1 | 9143\|0\|0 | 371402\|1\|0 | 1.10E-06 | 0.00E+00 |
| 19:15191450:T:C | Exon 6 | EGFr 7-34 | EGFr 8 | c.1010A>G | p.Tyr337Cys | NC | NC | 1.16E-06 | 1 | 9143\|0\|0 | 371402\|1\|0 | 1.10E-06 | 0.00E+00 |
| 19:15189386:C:T | Exon 7 | EGFr 7-34 | EGFr 9 | c.1079G>A | p.Cys360Tyr | NC | NC | 1.16E-06 | 1 | 9143\|0\|0 | 371402\|1\|0 | 1.10E-06 | 0.00E+00 |
| 19:15189145:A:G | Exon 8 | EGFr 7-34 | EGFr 10 | c.1222T>C | p.Cys408Arg | NC | NC | 1.16E-06 | 1 | 9143\|0\|0 | 371402\|1\|0 | 1.10E-06 | 0.00E+00 |
| 19:15189112:A:T | Exon 8 | EGFr 7-34 | EGFr 10 | c.1255T>A | p.Cys419Ser | NC | NC | 1.16E-06 | 1 | 9143\|0\|0 | 371402\|1\|0 | 1.10E-06 | 0.00E+00 |
| 19:15189106:G:A | Exon 8 | EGFr 7-34 | EGFr 10 | c.1261C>T | p.Arg421Cys | NC | NC | 2.32E-06 | 2 | 9143\|0\|0 | 371401\|2\|0 | 2.20E-06 | 0.00E+00 |
| 19:15189083:A:C | Exon 8 | EGFr 7-34 | EGFr 10 | c.1284T>G | p.Cys428Trp | 0.36 (0,587.18) | 7.88E-01 | 6.57E-06 | 5 | 9143\|0\|0 | 371398\|5\|0 | 5.50E-06 | 0.00E+00 |
| 19:15189003:C:T | Exon 8 | EGFr 7-34 | EGFr 11 | c.1364G>A | p.Cys455Tyr | NC | NC | 1.16E-06 | 1 | 9143\|0\|0 | 371402\|1\|0 | 1.10E-06 | 0.00E+00 |
| 19:15188301:T:A | Exon 9 | EGFr 7-34 | EGFr 12 | c.1426A>T | p.Ser476Cys | NC | NC | 3.48E-06 | 3 | 9143\|0\|0 | 371400\|3\|0 | 3.30E-06 | 0.00E+00 |
| 19:15188286:C:A | Exon 9 | EGFr 7-34 | EGFr 12 | c.1441G>T | p.Gly481Cys | NC | NC | 2.32E-06 | 2 | 9143\|0\|0 | 371399\|2\|0 | 2.20E-06 | 0.00E+00 |
| 19:15187977:A:G | Exon 10 | EGFr 7-34 | EGFr 12 | c.1510T>C | p.Cys504Arg | NC | NC | 1.16E-06 | 1 | 9143\|0\|0 | 371402\|1\|0 | 1.10E-06 | 0.00E+00 |
| 19:15187940:C:A | Exon 10 | EGFr 7-34 | EGFr 13 | c.1547G>T | p.Cys516Phe | 0.36 (0,45.79) | 6.77E-01 | 9.20E-06 | 7 | 9142\|0\|0 | 371396\|7\|0 | 7.70E-06 | 0.00E+00 |
| 19:15187940:C:T | Exon 10 | EGFr 7-34 | EGFr 13 | c.1547G>A | p.Cys516Tyr | NC | NC | 1.16E-06 | 1 | 9142\|1\|0 | 371396\|0\|0 | 1.10E-06 | 0.00E+00 |
| 19:15187922:C:G | Exon 10 | EGFr 7-34 | EGFr 13 | c.1565G>C | p.Cys522Ser | NC | NC | 1.16E-06 | 1 | 9143\|0\|0 | 371402\|1\|0 | 1.10E-06 | 0.00E+00 |
| 19:15187893:G:A | Exon 10 | EGFr 7-34 | EGFr 13 | c.1594C>T | p.Arg532Cys | NC | NC | 4.64E-06 | 4 | 9143\|0\|0 | 371397\|4\|0 | 4.40E-06 | 0.00E+00 |
| 19:15187315:G:A | Exon 11 | EGFr 7-34 | No EGFr domain | c.1630C>T | p.Arg544Cys | NC | NC | 2.32E-06 | 2 | 9142\|1\|0 | 371398\|1\|0 | 1.65E-05 | 2.31E-04 |

| Chrom:pos:ref:alt | Exon | Domain class | EGFr Domain number | HGVS transcript | HGVS protein | Odds Ratio (95 % CI) | Pval | AAF | AAC | Case Counts | Control Counts | UKB AAF | PGR AAF |
|---|---|---|---|---|---|---|---|---|---|---|---|---|---|
| 19:15187284:C:A | Exon 11 | EGFr 7-34 | EGFr 14 | c.1661G>T | p.Cys554Phe | NC | NC | 1.16E-06 | 1 | 9143\|0\|0 | 371402\|1\|0 | 1.10E-06 | 0.00E+00 |
| 19:15187273:G:A | Exon 11 | EGFr 7-34 | EGFr 14 | c.1672C>T | p.Arg558Cys | 0.36 (0,28.51) | 6.45E-01 | 1.31E-05 | 10 | 9137\|0\|0 | 371191\|10\|0 | 4.40E-06 | 6.26E-06 |
| 19:15187213:G:A | Exon 11 | EGFr 7-34 | EGFr 14 | c.1732C>T | p.Arg578Cys | 0.35 (0.04,3.21) | 3.56E-01 | 4.60E-05 | 35 | 9138\|0\|0 | 371246\|35\|0 | 1.43E-05 | 4.38E-05 |
| 19:15187209:C:T | Exon 11 | EGFr 7-34 | EGFr 14 | c.1736G>A | p.Cys579Tyr | NC | NC | 1.16E-06 | 1 | 9143\|0\|0 | 371402\|1\|0 | 2.20E-06 | 0.00E+00 |
| 19:15187186:G:A | Exon 11 | EGFr 7-34 | EGFr 15 | c.1759C>T | p.Arg587Cys | NC | NC | 4.64E-06 | 3 | 9143\|0\|0 | 371381\|3\|0 | 7.70E-06 | 1.25E-05 |
| 19:15187171:G:A | Exon 11 | EGFr 7-34 | EGFr 15 | c.1774C>T | p.Arg592Cys | NC | NC | 1.16E-06 | 1 | 9143\|0\|0 | 371383\|1\|0 | 1.10E-06 | 0.00E+00 |
| 19:15187126:G:A | Exon 11 | EGFr 7-34 | EGFr 15 | c.1819C>T | p.Arg607Cys | 6.7 (1.22,36.78) | 2.86E-02 | 1.97E-05 | 15 | 9136\|2\|0 | 370946\|13\|0 | 7.70E-06 | 0.00E+00 |
| 19:15187121:G:C | Exon 11 | EGFr 7-34 | EGFr 15 | c.1824C>G | p.Cys608Trp | NC | NC | 1.16E-06 | 1 | 9143\|0\|0 | 371401\|1\|0 | 1.10E-06 | 0.00E+00 |
| 19:15186911:G:A | Exon 12 | EGFr 7-34 | EGFr 16 | c.1918C>T | p.Arg640Cys | 14.67 (2.23,96.37) | 5.15E-03 | 1.45E-05 | 11 | 9141\|2\|0 | 371394\|9\|0 | 2.20E-05 | 0.00E+00 |
| 19:15185670:C:T | Exon 13 | EGFr 7-34 | EGFr 16 | c.1961G>A | p.Cys654Tyr | NC | NC | 1.16E-06 | 1 | 9143\|0\|0 | 371402\|1\|0 | 1.10E-06 | 0.00E+00 |
| 19:15185632:C:A | Exon 13 | EGFr 7-34 | EGFr 17 | c.1999G>T | p.Gly667Cys | NC | NC | 2.32E-06 | 1 | 9143\|0\|0 | 371398\|1\|0 | 2.20E-06 | 0.00E+00 |
| 19:15185619:G:C | Exon 13 | EGFr 7-34 | EGFr 17 | c.2012C>G | p.Ser671Cys | NC | NC | 4.64E-06 | 3 | 9143\|0\|0 | 371400\|3\|0 | 4.40E-06 | 0.00E+00 |
| 19:15185616:C:T | Exon 13 | EGFr 7-34 | EGFr 17 | c.2015G>A | p.Cys672Tyr | NC | NC | 1.16E-06 | 1 | 9143\|0\|0 | 371402\|1\|0 | 1.10E-06 | 0.00E+00 |
| 19:15185593:G:A | Exon 13 | EGFr 7-34 | EGFr 17 | c.2038C>T | p.Arg680Cys | NC | NC | 2.32E-06 | 1 | 9142\|1\|0 | 371403\|0\|0 | 3.30E-06 | 2.50E-05 |
| 19:15185404:G:A | Exon 14 | EGFr 7-34 | EGFr 18 | c.2149C>T | p.Arg717Cys | 0.35 (0.03,3.68) | 3.83E-01 | 3.68E-05 | 28 | 9143\|0\|0 | 371375\|28\|0 | 3.19E-05 | 0.00E+00 |
| 19:15185394:C:T | Exon 14 | EGFr 7-34 | EGFr 18 | c.2159G>A | p.Cys720Tyr | NC | NC | 1.16E-06 | 1 | 9143\|0\|0 | 371402\|1\|0 | 1.10E-06 | 0.00E+00 |
| 19:15185371:G:A | Exon 14 | EGFr 7-34 | EGFr 18 | c.2182C>T | p.Arg728Cys | 0.36 (0,43.27) | 6.74E-01 | 1.45E-05 | 11 | 9143\|0\|0 | 371392\|11\|0 | 1.65E-05 | 1.25E-05 |
| 19:15185334:G:C | Exon 14 | EGFr 7-34 | EGFr 19 | c.2219C>G | p.Ser740Cys | NC | NC | 1.16E-06 | 1 | 9143\|0\|0 | 371402\|1\|0 | 1.10E-06 | 0.00E+00 |
| 19:15185280:C:T | Exon 14 | EGFr 7-34 | EGFr 19 | c.2273G>A | p.Cys758Tyr | NC | NC | 1.16E-06 | 1 | 9143\|0\|0 | 371402\|1\|0 | 1.10E-06 | 0.00E+00 |
| 19:15185017:G:A | Exon 15 | EGFr 7-34 | EGFr 19 | c.2299C>T | p.Arg767Cys | 0.35 (0,69) | 6.99E-01 | 1.04E-05 | 7 | 9143\|0\|0 | 371382\|7\|0 | 1.10E-05 | 1.25E-05 |
| 19:15184963:G:A | Exon 15 | EGFr 7-34 | EGFr 20 | c.2353C>T | p.Arg785Cys | 23.74 (0.42,1332.09) | 1.23E-01 | 1.05E-05 | 8 | 9142\|1\|0 | 371396\|7\|0 | 1.32E-05 | 0.00E+00 |
| 19:15184929:C:G | Exon 15 | EGFr 7-34 | EGFr 20 | c.2387G>C | p.Cys796Ser | NC | NC | 1.16E-06 | 1 | 9143\|0\|0 | 371402\|1\|0 | 1.10E-06 | 0.00E+00 |
| 19:15184923:C:T | Exon 15 | EGFr 7-34 | EGFr 20 | c.2393G>A | p.Cys798Tyr | NC | NC | 2.32E-06 | 2 | 9143\|0\|0 | 371401\|2\|0 | 2.20E-06 | 0.00E+00 |
| 19:15184910:C:A | Exon 15 | EGFr 7-34 | EGFr 20 | c.2406G>T | p.Trp802Cys | 21 (1.7,260) | 1.83E-02 | 6.96E-06 | 5 | 9142\|1\|0 | 371398\|4\|0 | 6.60E-06 | 0.00E+00 |
| 19:15181794:G:C | Exon 17 | EGFr 7-34 | EGFr 22 | c.2574C>G | p.Cys858Trp | NC | NC | 1.16E-06 | 1 | 9143\|0\|0 | 371401\|1\|0 | 1.10E-06 | 0.00E+00 |
| 19:15181751:A:G | Exon 17 | EGFr 7-34 | EGFr 22 | c.2617T>C | p.Cys873Arg | NC | NC | 1.16E-06 | 1 | 9143\|0\|0 | 371402\|1\|0 | 1.10E-06 | 0.00E+00 |
| 19:15181750:C:A | Exon 17 | EGFr 7-34 | EGFr 22 | c.2618G>T | p.Cys873Phe | NC | NC | 1.16E-06 | 1 | 9143\|0\|0 | 371402\|1\|0 | 1.10E-06 | 0.00E+00 |

 

| Chrom:pos:ref:alt | Exon | Domain class | EGFr Domain number | HGVS transcript | HGVS protein | Odds Ratio (95 % CI) | Pval | AAF | AAC | Case Counts | Control Counts | UKB AAF | PGR AAF |
|---|---|---|---|---|---|---|---|---|---|---|---|---|---|
| 19:15181640:A:G | Exon 17 | EGFr 7-34 | EGFr 23 | c.2728T>C | p.Cys910Arg | NC | NC | 2.32E-06 | 2 | 9143\|0\|0 | 371401\|2\|0 | 2.20E-06 | 0.00E+00 |
| 19:15181639:C:T | Exon 17 | EGFr 7-34 | EGFr 23 | c.2729G>A | p.Cys910Tyr | NC | NC | 5.80E-06 | 4 | 9143\|0\|0 | 371399\|4\|0 | 5.50E-06 | 0.00E+00 |
| 19:15181633:C:G | Exon 17 | EGFr 7-34 | EGFr 23 | c.2735G>C | p.Cys912Ser | NC | NC | 2.32E-06 | 2 | 9143\|0\|0 | 371401\|2\|0 | 2.20E-06 | 0.00E+00 |
| 19:15181621:T:C | Exon 17 | EGFr 7-34 | EGFr 23 | c.2747A>G | p.Tyr916Cys | 0.35 (0.01,17.32) | 5.99E-01 | 1.18E-05 | 9 | 9143\|0\|0 | 371394\|9\|0 | 1.10E-05 | 0.00E+00 |
| 19:15181139:C:G | Exon 18 | EGFr 7-34 | EGFr 24 | c.2816G>C | p.Cys939Ser | NC | NC | 2.32E-06 | 2 | 9143\|0\|0 | 371401\|2\|0 | 2.20E-06 | 0.00E+00 |
| 19:15181043:C:T | Exon 18 | EGFr 7-34 | EGFr 25 | c.2912G>A | p.Cys971Tyr | NC | NC | 1.16E-06 | 1 | 9143\|0\|0 | 371402\|1\|0 | 1.10E-06 | 0.00E+00 |
| 19:15180999:A:G | Exon 18 | EGFr 7-34 | EGFr 25 | c.2956T>C | p.Cys986Arg | NC | NC | 1.16E-06 | 1 | 9143\|0\|0 | 371402\|1\|0 | 1.10E-06 | 1.25E-05 |
| 19:15180807:G:A | Exon 19 | EGFr 7-34 | EGFr 26 | c.3016C>T | p.Arg1006Cys | NC | NC | 1.16E-06 | 1 | 9143\|0\|0 | 371402\|1\|0 | 1.10E-06 | 0.00E+00 |
| 19:15180780:A:G | Exon 19 | EGFr 7-34 | EGFr 26 | c.3043T>C | p.Cys1015Arg | NC | NC | 1.16E-06 | 1 | 9143\|0\|0 | 371402\|1\|0 | 1.10E-06 | 0.00E+00 |
| 19:15180732:G:A | Exon 19 | EGFr 7-34 | EGFr 26 | c.3091C>T | p.Arg1031Cys | NC | NC | 3.48E-06 | 3 | 9143\|0\|0 | 371400\|3\|0 | 3.30E-06 | 0.00E+00 |
| 19:15180235:C:T | Exon 20 | EGFr 7-34 | EGFr 27 | c.3164G>A | p.Cys1055Tyr | NC | NC | 1.16E-06 | 1 | 9143\|0\|0 | 371402\|1\|0 | 1.10E-06 | 0.00E+00 |
| 19:15180217:C:T | Exon 20 | EGFr 7-34 | EGFr 27 | c.3182G>A | p.Cys1061Tyr | NC | NC | 3.48E-06 | 3 | 9143\|0\|0 | 371400\|3\|0 | 3.30E-06 | 0.00E+00 |
| 19:15180173:G:A | Exon 20 | EGFr 7-34 | EGFr 27 | c.3226C>T | p.Arg1076Cys | NC | NC | 1.16E-06 | 1 | 9143\|0\|0 | 371402\|1\|0 | 1.10E-06 | 1.25E-05 |
| 19:15180101:G:A | Exon 20 | EGFr 7-34 | EGFr 28 | c.3298C>T | p.Arg1100Cys | NC | NC | 2.32E-06 | 2 | 9143\|0\|0 | 371401\|2\|0 | 3.30E-06 | 0.00E+00 |
| 19:15180077:A:G | Exon 20 | EGFr 7-34 | EGFr 28 | c.3322T>C | p.Cys1108Arg | NC | NC | 2.32E-06 | 2 | 9143\|0\|0 | 371401\|2\|0 | 2.20E-06 | 0.00E+00 |
| 19:15179496:A:G | Exon 21 | EGFr 7-34 | EGFr 28 | c.3328T>C | p.Cys1110Arg | NC | NC | 4.64E-06 | 3 | 9143\|0\|0 | 371400\|3\|0 | 4.40E-06 | 0.00E+00 |
| 19:15179468:C:T | Exon 21 | EGFr 7-34 | EGFr 28 | c.3356G>A | p.Cys1119Tyr | 0.36 (0,810.78) | 7.96E-01 | 7.88E-06 | 6 | 9143\|0\|0 | 371397\|6\|0 | 6.60E-06 | 0.00E+00 |
| 19:15179415:A:G | Exon 21 | EGFr 7-34 | EGFr 29 | c.3409T>C | p.Cys1137Arg | NC | NC | 1.16E-06 | 1 | 9143\|0\|0 | 371401\|1\|0 | 1.10E-06 | 0.00E+00 |
| 19:15179397:G:A | Exon 21 | EGFr 7-34 | EGFr 29 | c.3427C>T | p.Arg1143Cys | 2.85 (0.9,9.04) | 7.59E-02 | 1.60E-04 | 122 | 9137\|6\|0 | 371287\|116\|0 | 1.46E-04 | 0.00E+00 |
| 19:15179393:T:C | Exon 21 | EGFr 7-34 | EGFr 29 | c.3431A>G | p.Tyr1144Cys | NC | NC | 2.32E-06 | 2 | 9143\|0\|0 | 371401\|2\|0 | 2.20E-06 | 0.00E+00 |
| 19:15179381:C:A | Exon 21 | EGFr 7-34 | EGFr 29 | c.3443G>T | p.Cys1148Phe | NC | NC | 1.16E-06 | 1 | 9143\|0\|0 | 371402\|1\|0 | 1.10E-06 | 6.26E-06 |
| 19:15179274:A:G | Exon 22 | EGFr 7-34 | EGFr 29 | c.3469T>C | p.Cys1157Arg | NC | NC | 1.16E-06 | 1 | 9143\|0\|0 | 371402\|1\|0 | 1.10E-06 | 0.00E+00 |
| 19:15179250:C:A | Exon 22 | EGFr 7-34 | EGFr 30 | c.3493G>T | p.Gly1165Cys | NC | NC | 3.48E-06 | 3 | 9143\|0\|0 | 371377\|3\|0 | 3.30E-06 | 0.00E+00 |
| 19:15179216:C:T | Exon 22 | EGFr 7-34 | EGFr 30 | c.3527G>A | p.Cys1176Tyr | NC | NC | 1.16E-06 | 1 | 9143\|0\|0 | 371402\|1\|0 | 1.10E-06 | 0.00E+00 |
| 19:15179197:G:C | Exon 22 | EGFr 7-34 | EGFr 30 | c.3546C>G | p.Cys1182Trp | NC | NC | 1.16E-06 | 1 | 9143\|0\|0 | 371386\|1\|0 | 1.10E-06 | 0.00E+00 |
| 19:15179175:G:A | Exon 22 | EGFr 7-34 | EGFr 30 | c.3568C>T | p.Arg1190Cys | 0.36 (0.01,9.43) | 5.36E-01 | 1.97E-05 | 15 | 9143\|0\|0 | 371388\|15\|0 | 2.31E-05 | 1.31E-04 |
| 19:15179165:C:T | Exon 22 | EGFr 7-34 | EGFr 30 | c.3578G>A | p.Cys1193Tyr | NC | NC | 1.16E-06 | 1 | 9143\|0\|0 | 371402\|1\|0 | 1.10E-06 | 0.00E+00 |

| **Chrom:pos:ref:alt** | **Exon** | **Domain class** | **EGFr Domain number** | **HGVS transcript** | **HGVS protein** | **Odds Ratio (95 % CI)** | **Pval** | **AAF** | **AAC** | **Case Counts** | **Control Counts** | **UKB AAF** | **PGR AAF** |
|---|---|---|---|---|---|---|---|---|---|---|---|---|---|
| 19:15179142:G:A | Exon 22 | EGFr 7-34 | EGFr 30 | c.3601C>T | p.Arg1201Cys | 0.36 (0.04,3.3) | 3.64E-01 | 5.26E-05 | 40 | 9143\|0\|0 | 371363\|40\|0 | 5.72E-05 | 6.26E-06 |
| 19:15179115:G:A | Exon 22 | EGFr 7-34 | EGFr 31 | c.3628C>T | p.Arg1210Cys | NC | NC | 3.48E-06 | 3 | 9142\|1\|0 | 371401\|2\|0 | 3.30E-06 | 6.26E-06 |
| 19:15179079:A:C | Exon 22 | EGFr 7-34 | EGFr 31 | c.3664T>G | p.Cys1222Gly | 2.3 (1.13,4.68) | 2.15E-02 | 2.46E-04 | 187 | 9133\|10\|0 | 371225\|177\|0 | 2.31E-04 | 0.00E+00 |
| 19:15179079:A:G | Exon 22 | EGFr 7-34 | EGFr 31 | c.3664T>C | p.Cys1222Arg | NC | NC | 1.16E-06 | 1 | 9133\|0\|0 | 371225\|1\|0 | 1.10E-06 | 0.00E+00 |
| 19:15179052:G:A | Exon 22 | EGFr 7-34 | EGFr 31 | c.3691C>T | p.Arg1231Cys | 3.38 (1.65,6.94) | 8.80E-04 | 1.97E-04 | 150 | 9124\|11\|0 | 370986\|139\|0 | 2.81E-04 | 5.34E-03 |
| 19:15178936:G:A | Exon 23 | EGFr 7-34 | EGFr 31 | c.3724C>T | p.Arg1242Cys | 34.42 (0.59,2022.49) | 8.86E-02 | 1.45E-05 | 10 | 9142\|1\|0 | 371394\|8\|1 | 1.21E-05 | 6.26E-06 |
| 19:15178877:G:C | Exon 23 | EGFr 7-34 | EGFr 32 | c.3783C>G | p.Cys1261Trp | NC | NC | 1.16E-06 | 1 | 9143\|0\|0 | 371402\|1\|0 | 1.10E-06 | 0.00E+00 |
| 19:15178836:C:G | Exon 23 | EGFr 7-34 | EGFr 32 | c.3824G>C | p.Cys1275Ser | NC | NC | 1.16E-06 | 1 | 9142\|1\|0 | 371403\|0\|0 | 1.10E-06 | 0.00E+00 |
| 19:15178081:C:A | Exon 24 | EGFr 7-34 | EGFr 32 | c.3847G>T | p.Gly1283Cys | NC | NC | 1.16E-06 | 1 | 9143\|0\|0 | 371402\|1\|0 | 1.10E-06 | 0.00E+00 |
| 19:15178057:G:A | Exon 24 | EGFr 7-34 | EGFr 33 | c.3871C>T | p.Arg1291Cys | NC | NC | 2.32E-06 | 1 | 9142\|1\|0 | 371402\|0\|0 | 2.20E-06 | 0.00E+00 |
| 19:15178050:C:A | Exon 24 | EGFr 7-34 | EGFr 33 | c.3878G>T | p.Cys1293Phe | NC | NC | 1.16E-06 | 1 | 9143\|0\|0 | 371402\|1\|0 | 1.10E-06 | 0.00E+00 |
| 19:15177990:C:G | Exon 24 | EGFr 7-34 | EGFr 33 | c.3938G>C | p.Cys1313Ser | NC | NC | 1.16E-06 | 1 | 9143\|0\|0 | 371402\|1\|0 | 1.10E-06 | 0.00E+00 |
| 19:15177984:C:A | Exon 24 | EGFr 7-34 | EGFr 33 | c.3944G>T | p.Cys1315Phe | 13.5 (2.14,85.31) | 5.67E-03 | 1.31E-05 | 10 | 9141\|2\|0 | 371395\|8\|0 | 1.10E-05 | 0.00E+00 |
| 19:15177983:G:C | Exon 24 | EGFr 7-34 | EGFr 33 | c.3945C>G | p.Cys1315Trp | NC | NC | 1.16E-06 | 1 | 9143\|0\|0 | 371402\|1\|0 | 1.10E-06 | 0.00E+00 |
| 19:15177958:A:T | Exon 24 | EGFr 7-34 | EGFr 33 | c.3970T>A | p.Cys1324Ser | NC | NC | 2.32E-06 | 2 | 9143\|0\|0 | 371401\|2\|0 | 2.20E-06 | 0.00E+00 |
| 19:15177957:C:T | Exon 24 | EGFr 7-34 | EGFr 33 | c.3971G>A | p.Cys1324Tyr | NC | NC | 1.16E-06 | 1 | 9143\|0\|0 | 371402\|1\|0 | 1.10E-06 | 0.00E+00 |
| 19:15177850:G:A | Exon 24 | EGFr 7-34 | EGFr 34 | c.4078C>T | p.Arg1360Cys | NC | NC | 1.16E-06 | 1 | 9143\|0\|0 | 371402\|1\|0 | 1.10E-06 | 6.26E-06 |
| 19:15177812:G:C | Exon 24 | EGFr 7-34 | EGFr 34 | c.4116C>G | p.Cys1372Trp | NC | NC | 1.16E-06 | 1 | 9143\|0\|0 | 371402\|1\|0 | 1.10E-06 | 0.00E+00 |
| 19:15177763:G:A | Exon 24 | No EGFr | No EGFr domain | c.4165C>T | p.Arg1389Cys | 0.35 (0.02,7.94) | 5.11E-01 | 1.84E-05 | 14 | 9143\|0\|0 | 371388\|14\|0 | 1.98E-05 | 0.00E+00 |
| 19:15177727:A:C | Exon 24 | No EGFr | No EGFr domain | c.4201T>G | p.Cys1401Gly | NC | NC | 3.48E-06 | 2 | 9143\|0\|0 | 371401\|2\|0 | 3.30E-06 | 0.00E+00 |
| 19:15177697:C:A | Exon 24 | No EGFr | No EGFr domain | c.4231G>T | p.Gly1411Cys | NC | NC | 1.16E-06 | 1 | 9143\|0\|0 | 371401\|1\|0 | 1.10E-06 | 0.00E+00 |
| 19:15174368:A:C | Exon 25 | No EGFr | No EGFr domain | c.4436T>G | p.Phe1479Cys | NC | NC | 3.48E-06 | 3 | 9143\|0\|0 | 371400\|3\|0 | 3.30E-06 | 0.00E+00 |
| 19:15174357:G:A | Exon 25 | No EGFr | No EGFr domain | c.4447C>T | p.Arg1483Cys | 0.36 (0.01,19.71) | 6.15E-01 | 1.97E-05 | 15 | 9143\|0\|0 | 371388\|15\|0 | 1.87E-05 | 0.00E+00 |
| 19:15174300:T:A | Exon 25 | No EGFr | No EGFr domain | c.4504A>T | p.Ser1502Cys | NC | NC | 1.16E-06 | 1 | 9143\|0\|0 | 371402\|1\|0 | 1.10E-06 | 0.00E+00 |
| 19:15174228:G:A | Exon 25 | No EGFr | No EGFr domain | c.4576C>T | p.Arg1526Cys | NC | NC | 1.16E-06 | 1 | 9143\|0\|0 | 371402\|1\|0 | 1.10E-06 | 0.00E+00 |
| 19:15174224:G:C | Exon 25 | No EGFr | No EGFr domain | c.4580C>G | p.Ser1527Cys | NC | NC | 2.32E-06 | 2 | 9143\|0\|0 | 371401\|2\|0 | 2.20E-06 | 0.00E+00 |
| 19:15174168:G:A | Exon 25 | No EGFr | No EGFr domain | c.4636C>T | p.Arg1546Cys | 1.77 (0.15,21.26) | 6.54E-01 | 3.68E-05 | 28 | 9142\|1\|0 | 371376\|27\|0 | 3.30E-05 | 0.00E+00 |

| Chrom:pos:ref:alt | Exon | Domain class | EGFr Domain number | HGVS transcript | HGVS protein | Odds Ratio (95 % CI) | Pval | AAF | AAC | Case Counts | Control Counts | UKB AAF | PGR AAF |
|---|---|---|---|---|---|---|---|---|---|---|---|---|---|
| 19:15174153:C:A | Exon 25 | No EGFr | No EGFr domain | c.4651G>T | p.Gly1551Cys | NC | NC | 1.16E-06 | 1 | 9143\|0\|0 | 371402\|1\|0 | 1.10E-06 | 0.00E+00 |
| 19:15174096:G:A | Exon 25 | No EGFr | No EGFr domain | c.4708C>T | p.Arg1570Cys | 0.35 (0,47.28) | 6.77E-01 | 7.88E-06 | 6 | 9143\|0\|0 | 371397\|6\|0 | 8.80E-06 | 0.00E+00 |
| 19:15170707:G:A | Exon 26 | No EGFr | No EGFr domain | c.4855C>T | p.Arg1619Cys | NC | NC | 1.16E-06 | 1 | 9143\|0\|0 | 371401\|1\|0 | 1.10E-06 | 0.00E+00 |
| 19:15167371:C:T | Exon 29 | No EGFr | No EGFr domain | c.5240G>A | p.Cys1747Tyr | NC | NC | 3.48E-06 | 1 | 9143\|0\|0 | 371402\|1\|0 | 4.40E-06 | 0.00E+00 |
| 19:15167369:G:A | Exon 29 | No EGFr | No EGFr domain | c.5242C>T | p.Arg1748Cys | 0.35 (0.01,10.69) | 5.48E-01 | 2.89E-05 | 22 | 9143\|0\|0 | 371381\|22\|0 | 3.63E-05 | 6.26E-06 |
| 19:15167330:G:A | Exon 29 | No EGFr | No EGFr domain | c.5281C>T | p.Arg1761Cys | 0.35 (0,27) | 6.35E-01 | 9.28E-06 | 7 | 9143\|0\|0 | 371396\|7\|0 | 8.80E-06 | 6.26E-06 |
| 19:15167258:G:A | Exon 29 | No EGFr | No EGFr domain | c.5353C>T | p.Arg1785Cys | 0.34 (0,31.47) | 6.40E-01 | 9.20E-06 | 7 | 9143\|0\|0 | 371396\|7\|0 | 1.10E-05 | 6.26E-06 |
| 19:15166058:C:T | Exon 30 | No EGFr | No EGFr domain | c.5396G>A | p.Cys1799Tyr | 0.36 (0.01,11.04) | 5.55E-01 | 1.97E-05 | 15 | 9143\|0\|0 | 371388\|15\|0 | 1.98E-05 | 0.00E+00 |
| 19:15165945:G:A | Exon 30 | No EGFr | No EGFr domain | c.5509C>T | p.Arg1837Cys | 0.36 (0,320.69) | 7.68E-01 | 6.57E-06 | 5 | 9143\|0\|0 | 371372\|5\|0 | 8.80E-06 | 4.38E-05 |
| 19:15165912:G:A | Exon 30 | No EGFr | No EGFr domain | c.5542C>T | p.Arg1848Cys | NC | NC | 1.16E-06 | 1 | 9143\|0\|0 | 371402\|1\|0 | 1.10E-06 | 0.00E+00 |
| 19:15165903:G:A | Exon 30 | No EGFr | No EGFr domain | c.5551C>T | p.Arg1851Cys | NC | NC | 1.16E-06 | 1 | 9143\|0\|0 | 371401\|1\|0 | 1.10E-06 | 6.26E-06 |
| 19:15165837:G:A | Exon 30 | No EGFr | No EGFr domain | c.5617C>T | p.Arg1873Cys | 0.36 (0,78) | 7.07E-01 | 5.80E-06 | 5 | 9143\|0\|0 | 371398\|5\|0 | 5.50E-06 | 6.26E-06 |
| 19:15165500:G:A | Exon 31 | No EGFr | No EGFr domain | c.5683C>T | p.Arg1895Cys | 0.35 (0.01,14.7) | 5.82E-01 | 1.31E-05 | 10 | 9143\|0\|0 | 371393\|10\|0 | 1.10E-05 | 6.26E-06 |
| 19:15165479:G:A | Exon 31 | No EGFr | No EGFr domain | c.5704C>T | p.Arg1902Cys | 11.2 (2.37,52.89) | 2.28E-03 | 1.97E-05 | 15 | 9140\|3\|0 | 371391\|12\|0 | 2.42E-05 | 5.00E-05 |
| 19:15165440:G:A | Exon 31 | No EGFr | No EGFr domain | c.5743C>T | p.Arg1915Cys | NC | NC | 3.48E-06 | 3 | 9143\|0\|0 | 371400\|3\|0 | 3.30E-06 | 6.26E-06 |
| 19:15161674:T:C | Exon 33 | No EGFr | No EGFr domain | c.5954A>G | p.Tyr1985Cys | 0.35 (0.02,5.38) | 4.54E-01 | 3.28E-05 | 25 | 9143\|0\|0 | 371377\|25\|0 | 3.08E-05 | 4.38E-05 |
| 19:15161564:G:A | Exon 33 | No EGFr | No EGFr domain | c.6064C>T | p.Arg2022Cys | NC | NC | 1.16E-06 | 1 | 9143\|0\|0 | 371401\|1\|0 | 2.20E-06 | 6.26E-06 |
| 19:15161546:T:A | Exon 33 | No EGFr | No EGFr domain | c.6082A>T | p.Ser2028Cys | NC | NC | 2.32E-06 | 2 | 9143\|0\|0 | 371401\|2\|0 | 2.20E-06 | 0.00E+00 |
| 19:15161537:G:A | Exon 33 | No EGFr | No EGFr domain | c.6091C>T | p.Arg2031Cys | 5.18 (0.21,128.57) | 3.16E-01 | 1.97E-05 | 15 | 9142\|1\|0 | 371389\|14\|0 | 2.09E-05 | 1.88E-05 |
| 19:15161525:C:A | Exon 33 | No EGFr | No EGFr domain | c.6103G>T | p.Gly2035Cys | NC | NC | 2.32E-06 | 2 | 9143\|0\|0 | 371401\|2\|0 | 2.20E-06 | 1.25E-05 |
| 19:15161365:C:A | Exon 33 | No EGFr | No EGFr domain | c.6263G>T | p.Cys2088Phe | NC | NC | 1.16E-06 | 1 | 9143\|0\|0 | 371402\|1\|0 | 1.10E-06 | 0.00E+00 |
| 19:15161248:T:C | Exon 33 | No EGFr | No EGFr domain | c.6380A>G | p.Tyr2127Cys | NC | NC | 1.16E-06 | 1 | 9143\|0\|0 | 371402\|1\|0 | 2.20E-06 | 1.25E-05 |
| 19:15161221:G:C | Exon 33 | No EGFr | No EGFr domain | c.6407C>G | p.Ser2136Cys | NC | NC | 3.48E-06 | 3 | 9142\|1\|0 | 371401\|2\|0 | 3.30E-06 | 0.00E+00 |
| 19:15161207:C:A | Exon 33 | No EGFr | No EGFr domain | c.6421G>T | p.Gly2141Cys | NC | NC | 1.16E-06 | 1 | 9143\|0\|0 | 371402\|1\|0 | 1.10E-06 | 0.00E+00 |
| 19:15161180:G:A | Exon 33 | No EGFr | No EGFr domain | c.6448C>T | p.Arg2150Cys | 0.36 (0,34.1) | 6.58E-01 | 1.58E-05 | 12 | 9143\|0\|0 | 371391\|12\|0 | 1.32E-05 | 3.75E-05 |
| 19:15160928:G:A | Exon 33 | No EGFr | No EGFr domain | c.6700C>T | p.Arg2234Cys | 0.67 (0.14,3.23) | 6.20E-01 | 8.93E-05 | 68 | 9142\|1\|0 | 371336\|67\|0 | 8.25E-05 | 7.51E-05 |
| 19:15160830:C:G | Exon 33 | No EGFr | No EGFr domain | c.6798G>C | p.Trp2266Cys | NC | NC | 1.16E-06 | 1 | 9143\|0\|0 | 371402\|1\|0 | 1.10E-06 | 0.00E+00 |

1. To test the combined signal of association of rare *NOTCH3* Cys-altering variants with ischemic stroke in the UKB European population, individual variant data was combined into a burden test. Shown is a table of all Cys-altering variants in *NOTCH3* that were observed in UKB Europeans, including variant functional information, and single variant association summary statistics for variants with alternate allele count (AAC) ≥ 5. From left-to-right is variant location (in chromosome:position:reference:alternate format with GRCh38 coordinates); exon number; EGFr domain class in *NOTCH3* (1-6, 7-34, or no EGFr domains); specific EGFr number; impact on ENST00000263388 transcript; impact on the protein sequence; single variant analysis effect size (with 95% confidence interval in parenthesis); single variant analysis p value; alternate variant allele frequency; alternate variant allele count; number of cases (organized by Ref/Ref homozygotes | Ref/Alt heterozygotes | Alt/Alt homozygotes); number of controls; alternate allele frequency in the full UKB 450 K cohort across ancestries; and alternate allele frequency in the full 75 K PGR cohort. “NC” stands for “Not Calculated”, meaning that the case and control counts of the variant was insufficient to calculate a single variant analysis p value and effect size. Table is ordered from start to end of transcript.

**Supplementary Table 23**. **Comparison of Cys-altering and Ser-altering Variants in *NOTCH3* EGFr domains in UKB[1].**

| Domain | Covariate | Variant group | Phenotype | Odds ratio (CI) | Pval | AAF | Case (RR\|RA\|AA) | Case (n) | Control (RR\|RA\|AA) | Control (n) |
|---|---|---|---|---|---|---|---|---|---|---|
| EGFr 1 to 34 | Standard | Cys-alt | Ischemic stroke | 2.86 (2.14,3.82) | 6.29E-10 | 9.99E-04 | 9094\|49\|0 | 9,143 | 370693\|709\|1 | 371,403 |
| EGFr 1 to 34 | Standard+Smoking | Cys-alt | Ischemic stroke | 2.82 (2.11,3.78) | 1.40E-09 | 9.99E-04 | 9034\|48\|0 | 9,082 | 369103\|707\|1 | 369,811 |
| EGFr 1 to 34 | Standard+Smoking+HTN | Cys-alt | Ischemic stroke | 2.72 (2,3.71) | 2.91E-08 | 1.00E-03 | 8298\|43\|0 | 8,341 | 287673\|551\|1 | 288,225 |
| EGFr 1 to 6 | Standard | Cys-alt | Ischemic stroke | 29.51 (10.39,83.82) | 1.37E-07 | 1.97E-05 | 9137\|6\|0 | 9,143 | 371394\|9\|0 | 371,403 |
| EGFr 1 to 6 | Standard+Smoking | Cys-alt | Ischemic stroke | 24.62 (8.18,74.11) | 2.74E-06 | 1.85E-05 | 9077\|5\|0 | 9,082 | 369802\|9\|0 | 369,811 |
| EGFr 1 to 6 | Standard+Smoking+HTN | Cys-alt | Ischemic stroke | 26.91 (7.47,96.98) | 1.83E-05 | 1.69E-05 | 8337\|4\|0 | 8,341 | 288219\|6\|0 | 288,225 |
| EGFr 7 to 34 | Standard | Cys-alt | Ischemic stroke | 2.55 (1.87,3.46) | 1.59E-07 | 9.79E-04 | 9100\|43\|0 | 9,143 | 370702\|700\|1 | 371,403 |
| EGFr 7 to 34 | Standard+Smoking | Cys-alt | Ischemic stroke | 2.56 (1.89,3.49) | 1.30E-07 | 9.80E-04 | 9039\|43\|0 | 9,082 | 369112\|698\|1 | 369,811 |
| EGFr 7 to 34 | Standard+Smoking+HTN | Cys-alt | Ischemic stroke | 2.5 (1.81,3.45) | 1.00E-06 | 9.88E-04 | 8302\|39\|0 | 8,341 | 287679\|545\|1 | 288,225 |
| Not in EGFr | Standard | Cys-alt | Ischemic stroke | 0.97 (0.46,2.03) | 9.30E-01 | 3.93E-04 | 9136\|7\|0 | 9,143 | 371111\|292\|0 | 371,403 |
| Not in EGFr | Standard+Smoking | Cys-alt | Ischemic stroke | 0.97 (0.46,2.03) | 9.37E-01 | 3.93E-04 | 9075\|7\|0 | 9,082 | 369520\|291\|0 | 369,811 |
| Not in EGFr | Standard+Smoking+HTN | Cys-alt | Ischemic stroke | 0.9 (0.42,1.94) | 7.82E-01 | 4.08E-04 | 8335\|6\|0 | 8,341 | 287989\|236\|0 | 288,225 |
| EGFr 1 to 34 | Standard | Ser-alt | Ischemic stroke | 0.98 (0.8,1.2) | 8.35E-01 | 5.45E-03 | 9046\|97\|0 | 9,143 | 367355\|4042\|6 | 371,403 |
| EGFr 1 to 34 | Standard+Smoking | Ser-alt | Ischemic stroke | 0.98 (0.8,1.2) | 8.68E-01 | 5.45E-03 | 8985\|97\|0 | 9,082 | 365781\|4024\|6 | 369,811 |
| EGFr 1 to 34 | Standard+Smoking+HTN | Ser-alt | Ischemic stroke | 1.01 (0.82,1.25) | 9.20E-01 | 5.46E-03 | 8250\|91\|0 | 8,341 | 285080\|3140\|5 | 288,225 |
| EGFr 1 to 6 | Standard | Ser-alt | Ischemic stroke | 0.76 (0.23,2.56) | 6.57E-01 | 1.76E-04 | 9141\|2\|0 | 9,143 | 371271\|132\|0 | 371,403 |
| EGFr 1 to 6 | Standard+Smoking | Ser-alt | Ischemic stroke | 0.77 (0.23,2.63) | 6.78E-01 | 1.76E-04 | 9080\|2\|0 | 9,082 | 369680\|131\|0 | 369,811 |
| EGFr 1 to 6 | Standard+Smoking+HTN | Ser-alt | Ischemic stroke | 0.79 (0.22,2.8) | 7.19E-01 | 1.79E-04 | 8339\|2\|0 | 8,341 | 288121\|104\|0 | 288,225 |
| EGFr 7 to 34 | Standard | Ser-alt | Ischemic stroke | 0.99 (0.8,1.21) | 8.89E-01 | 5.28E-03 | 9048\|95\|0 | 9,143 | 367486\|3911\|6 | 371,403 |
| EGFr 7 to 34 | Standard+Smoking | Ser-alt | Ischemic stroke | 0.99 (0.81,1.22) | 9.19E-01 | 5.28E-03 | 8987\|95\|0 | 9,082 | 365911\|3894\|6 | 369,811 |
| EGFr 7 to 34 | Standard+Smoking+HTN | Ser-alt | Ischemic stroke | 1.02 (0.82,1.26) | 8.71E-01 | 5.29E-03 | 8252\|89\|0 | 8,341 | 285183\|3037\|5 | 288,225 |
| Not in EGFr | Standard | Ser-alt | Ischemic stroke | 0.84 (0.52,1.34) | 4.52E-01 | 1.01E-03 | 9128\|15\|0 | 9,143 | 370656\|744\|3 | 371,403 |
| Not in EGFr | Standard+Smoking | Ser-alt | Ischemic stroke | 0.85 (0.53,1.37) | 5.12E-01 | 1.00E-03 | 9067\|15\|0 | 9,082 | 369069\|739\|3 | 369,811 |
| Not in EGFr | Standard+Smoking+HTN | Ser-alt | Ischemic stroke | 0.9 (0.55,1.46) | 6.60E-01 | 1.02E-03 | 8326\|15\|0 | 8,341 | 287635\|588\|2 | 288,225 |

1. To test the hypothesis that all missense variants contribute to the association between *NOTCH3* and stroke, an analysis was conducted to compare the effects of Cys-altering and Ser-altering variants. Serine was selected because it is one of the most common amino acids added or removed in *NOTCH3*, and at *NOTCH3* codon 1231 a second alternate allele introduces an extremely rare Arg->Ser instead of a Arg->Cys

missense substitution. Shown in this table from left-to-right is a description of the NOTCH3 domains included in the analysis (EGFr domains 1 to 34, 1 to 6, 7 to 34, or the C-terminal region beyond EGFr 34); covariates included; variant group (Cys-altering or Ser-altering); phenotype; effect size (odds ratio with 95% confidence interval in parenthesis); p value; alternate allele frequency; case genotype counts (RR|RA|AA); case total n; control genotype counts (RR|RA|AA); control total n; and a description of the analysis. Results shown for standard covariates.

**Supplementary Table 24**. ***NOTCH3* LoF Variants in UKB Included in Burden Test[1].**

| Chrom:pos:ref:alt | Exon | EGFr Domain | Variant Effect | HGVS transcript | HGVS protein | Odds ratio (95% confidence interval) | P value | AAF | AAC | Cases | Controls | UKB 450K | PGR |
|---|---|---|---|---|---|---|---|---|---|---|---|---|---|
| 19:15200905:T:C | Exon 1 | No EGFr domain | start_lost | c.1A>G | p.Met1? | NC | NC | 1.1E-06 | 1 | 9082\|0\|0 | 368695\|1\|0 | 1.11E-06 | 0 |
| 19:15200904:A:C | Exon 1 | No EGFr domain | start_lost | c.2T>G | p.Met1? | NC | NC | 1.1E-06 | 1 | 9084\|0\|0 | 368727\|1\|0 | 1.11E-06 | 0 |
| 19:15200903:CA:C | Exon 1 | No EGFr domain | frameshift | c.2delT | p.Met1fs | NC | NC | 1.1E-06 | 1 | 9084\|0\|0 | 368727\|1\|0 | 1.11E-06 | 0 |
| 19:15200892:GC:G | Exon 1 | No EGFr domain | frameshift | c.13delG | p.Ala5fs | 13.35 (0.31, 568.42) | 0.176 | 7.9E-06 | 6 | 9097\|1\|0 | 369616\|5\|0 | 6.63E-06 | 0 |
| 19:15200793:GC:G | Exon 1 | No EGFr domain | frameshift | c.112delG | p.Ala38fs | 0.35 (0, 33.83) | 0.656 | 9.2E-06 | 7 | 9143\|0\|0 | 371388\|7\|0 | 7.7E-06 | 0 |
| 19:15197571:A:AG | Exon 2 | EGFr 1 | frameshift | c.125dupC | p.Cys43fs | NC | NC | 1.1E-06 | 1 | 9143\|0\|0 | 371401\|1\|0 | 1.1E-06 | 0 |
| 19:15192410:G:A | Exon 3 | EGFr 2 | stop_gained | c.307C>T | p.Arg103* | NC | NC | 1.1E-06 | 1 | 9142\|0\|0 | 371402\|1\|0 | 1.1E-06 | 0 |
| 19:15192380:G:A | Exon 3 | EGFr 2 | stop_gained | c.337C>T | p.Arg113* | NC | NC | 2.2E-06 | 2 | 9143\|0\|0 | 371401\|2\|0 | 2.2E-06 | 0 |
| 19:15192376:C:G | Exon 3 | No EGFr domain | splice_donor | c.340+1G>C | | NC | NC | 1.1E-06 | 1 | 9143\|0\|0 | 371402\|1\|0 | 1.1E-06 | 0 |
| 19:15192375:A:G | NA | No EGFr domain | splice_donor | c.340+2T>C | | NC | NC | 1.1E-06 | 1 | 9143\|0\|0 | 371402\|1\|0 | 1.1E-06 | 0 |
| 19:15192173:G:A | Exon 4 | EGFr 3 | stop_gained | c.466C>T | p.Arg156* | NC | NC | 2.2E-06 | 2 | 9143\|0\|0 | 371401\|2\|0 | 2.2E-06 | 0 |
| 19:15191827:A:T | Exon 5 | EGFr 6 | stop_gained | c.720T>A | p.Cys240* | NC | NC | 1.1E-06 | 1 | 9143\|0\|0 | 371402\|1\|0 | 1.1E-06 | 0 |
| 19:15191817:G:A | Exon 5 | EGFr 6 | stop_gained | c.730C>T | p.Arg244* | NC | NC | 2.2E-06 | 2 | 9143\|0\|0 | 371401\|2\|0 | 2.2E-06 | 0 |
| 19:15191799:T:TC | Exon 5 | EGFr 6 | frameshift | c.747dupG | p.Thr250fs | NC | NC | 1.1E-06 | 1 | 9143\|0\|0 | 371402\|1\|0 | 1.1E-06 | 0 |
| 19:15191446:A:T | Exon 6 | EGFr 8 | stop_gained | c.1014T>A | p.Cys338* | NC | NC | 1.1E-06 | 1 | 9143\|0\|0 | 371402\|1\|0 | 1.1E-06 | 0 |
| 19:15189368:CAGAT:C | Exon 7 | EGFr 9 | frameshift | c.1093_1096delATCT | p.Ile365fs | NC | NC | 1.1E-06 | 1 | 9143\|0\|0 | 371402\|1\|0 | 1.1E-06 | 0 |
| 19:15189271:A:T | NA | No EGFr domain | splice_donor | c.1192+2T>A | | NC | NC | 1.1E-06 | 1 | 9143\|0\|0 | 371402\|1\|0 | 1.1E-06 | 0 |
| 19:15189013:G:A | Exon 8 | EGFr 11 | stop_gained | c.1354C>T | p.Gln452* | NC | NC | 1.1E-06 | 1 | 9143\|0\|0 | 371401\|1\|0 | 1.1E-06 | 0 |
| 19:15188349:C:T | NA | No EGFr domain | splice_acceptor | c.1379-1G>A | | NC | NC | 1.1E-06 | 1 | 9143\|0\|0 | 371401\|1\|0 | 1.1E-06 | 0 |
| 19:15188271:C:CA | Exon 9 | EGFr 12 | frameshift | c.1455_1456insT | p.Asp486fs | NC | NC | 2.2E-06 | 2 | 9142\|1\|0 | 371402\|1\|0 | 2.2E-06 | 0 |
| 19:15188257:GC:G | Exon 9 | EGFr 12 | frameshift | c.1469delG | p.Gly490fs | NC | NC | 1.1E-06 | 1 | 9142\|0\|0 | 371309\|1\|0 | 1.1E-06 | 0 |
| 19:15187879:A:C | NA | No EGFr domain | splice_donor | c.1606+2T>G | | NC | NC | 1.1E-06 | 1 | 9142\|1\|0 | 371403\|0\|0 | 1.1E-06 | 0 |
| 19:15187245:G:C | Exon 11 | EGFr 14 | stop_gained | c.1700C>G | p.Ser567* | NC | NC | 1.1E-06 | 1 | 9143\|0\|0 | 371402\|1\|0 | 1.1E-06 | 0 |
| 19:15187104:C:A | Exon 11 | No EGFr domain | splice_donor | c.1840+1G>T | | NC | NC | 1.1E-06 | 1 | 9143\|0\|0 | 371402\|1\|0 | 1.1E-06 | 0 |
| 19:15186893:G:A | Exon 12 | EGFr 16 | stop_gained | c.1936C>T | p.Gln646* | NC | NC | 1.1E-06 | 1 | 9143\|0\|0 | 371402\|1\|0 | 1.1E-06 | 0 |
| 19:15185681:T:G | NA | No EGFr domain | splice_acceptor | c.1952-2A>C | | NC | NC | 1.1E-06 | 1 | 9143\|0\|0 | 371402\|1\|0 | 1.1E-06 | 0 |

| Chrom:pos:ref:alt | Exon | EGFr Domain | Variant Effect | HGVS transcript | HGVS protein | Odds ratio (95% confidence interval) | P value | AAF | AAC | Cases | Controls | UKB 450K | PGR |
|---|---|---|---|---|---|---|---|---|---|---|---|---|---|
| 19:15185568:A:T | Exon 13 | EGFr 17 | stop_gained | c.2063T>A | p.Leu688* | NC | NC | 1.1E-06 | 1 | 9143\|0\|0 | 371399\|1\|0 | 1.1E-06 | 0 |
| 19:15185501:A:T | Exon 13 | EGFr 18 | stop_gained | c.2130T>A | p.Tyr710* | NC | NC | 5.5E-06 | 3 | 9143\|0\|0 | 371399\|3\|0 | 5.5E-06 | 0 |
| 19:15185375:GC:G | Exon 14 | EGFr 18 | frameshift | c.2177delG | p.Gly726fs | NC | NC | 2.2E-06 | 2 | 9143\|0\|0 | 371401\|2\|0 | 2.2E-06 | 0 |
| 19:15185350:G:A | Exon 14 | EGFr 19 | stop_gained | c.2203C>T | p.Arg735* | 0.35 (0.03, 4.93) | 0.439 | 3.2E-05 | 24 | 9140\|0\|0 | 371224\|24\|0 | 4.4E-06 | 0 |
| 19:15185256:C:T | Exon 14 | No EGFr domain | splice_donor | c.2296+1G>A | | NC | NC | 1.1E-06 | 1 | 9143\|0\|0 | 371402\|1\|0 | 1.1E-06 | 0 |
| 19:15184922:G:T | Exon 15 | EGFr 20 | stop_gained | c.2394C>A | p.Cys798* | 0.36 (0, 77.63) | 0.707 | 6.6E-06 | 5 | 9143\|0\|0 | 371396\|5\|0 | 6.6E-06 | 0 |
| 19:15184419:A:T | Exon 16 | EGFr 21 | stop_gained | c.2442T>A | p.Cys814* | NC | NC | 1.1E-06 | 1 | 9143\|0\|0 | 371402\|1\|0 | 1.1E-06 | 0 |
| 19:15184382:TG:T | Exon 16 | EGFr 21 | frameshift | c.2478delC | p.Cys826fs | NC | NC | 1.1E-06 | 1 | 9143\|0\|0 | 371402\|1\|0 | 1.1E-06 | 0 |
| 19:15184367:TC:T | Exon 16 | EGFr 21 | frameshift | c.2493delG | p.Ser832fs | NC | NC | 4.4E-06 | 2 | 9143\|0\|0 | 371401\|2\|0 | 4.4E-06 | 0 |
| 19:15184356:GC:G | Exon 16 | EGFr 21 | frameshift | c.2504delG | p.Cys835fs | NC | NC | 2.2E-06 | 2 | 9143\|0\|0 | 371401\|2\|0 | 2.2E-06 | 0 |
| 19:15181668:G:GGT | Exon 17 | EGFr 23 | frameshift | c.2698_2699dupAC | p.Cys901fs | NC | NC | 1.1E-06 | 1 | 9143\|0\|0 | 371402\|1\|0 | 1.1E-06 | 0 |
| 19:15181577:T:TG | Exon 17 | EGFr 24 | frameshift | c.2790dupC | p.Ser931fs | NC | NC | 2.2E-06 | 1 | 9143\|0\|0 | 371402\|1\|0 | 2.2E-06 | 0 |
| 19:15181575:C:T | Exon 17 | No EGFr domain | splice_donor | c.2792+1G>A | | NC | NC | 5.5E-06 | 5 | 9143\|0\|0 | 371398\|5\|0 | 5.5E-06 | 0 |
| 19:15181164:T:C | NA | No EGFr domain | splice_acceptor | c.2793-2A>G | | 0.36 (0, 41.93) | 0.673 | 1.5E-05 | 11 | 9143\|0\|0 | 371392\|11\|0 | 1.54E-05 | 0 |
| 19:15180973:GC:G | Exon 18 | EGFr 25 | frameshift | c.2981delG | p.Gly994fs | NC | NC | 2.2E-06 | 2 | 9143\|0\|0 | 371386\|2\|0 | 2.2E-06 | 0 |
| 19:15180970:C:CACCCGGCCCGTGAA | Exon 18 | EGFr 25 | frameshift | c.2984_2985insTTCACGGGCCGGGT | p.Gln996fs | NC | NC | 1.1E-06 | 1 | 9143\|0\|0 | 371399\|1\|0 | 1.1E-06 | 0 |
| 19:15180970:CG:C | Exon 18 | EGFr 25 | frameshift | c.2984delC | p.Pro995fs | NC | NC | 1.1E-06 | 1 | 9143\|0\|0 | 371399\|1\|0 | 1.1E-06 | 0 |
| 19:15180703:G:T | Exon 19 | EGFr 27 | stop_gained | c.3120C>A | p.Cys1040* | NC | NC | 1.1E-06 | 1 | 9141\|1\|0 | 371394\|0\|0 | 1.1E-06 | 0 |
| 19:15180084:G:GC | Exon 20 | EGFr 28 | frameshift | c.3314dupG | p.Tyr1106fs | NC | NC | 4.4E-06 | 3 | 9143\|0\|0 | 371399\|3\|0 | 4.4E-06 | 0 |
| 19:15180075:A:T | Exon 20 | EGFr 28 | stop_gained | c.3324T>A | p.Cys1108* | NC | NC | 1.1E-06 | 1 | 9143\|0\|0 | 371402\|1\|0 | 1.1E-06 | 0 |
| 19:15180070:A:C | NA | No EGFr domain | splice_donor | c.3327+2T>G | | NC | NC | 1.1E-06 | 1 | 9143\|0\|0 | 371402\|1\|0 | 1.1E-06 | 0 |
| 19:15179456:ACG:A | Exon 21 | EGFr 29 | frameshift | c.3366_3367delCG | p.Val1123fs | NC | NC | 1.1E-06 | 1 | 9143\|0\|0 | 371401\|1\|0 | 1.1E-06 | 0 |
| 19:15179414:CA:C | Exon 21 | EGFr 29 | frameshift | c.3409delT | p.Cys1137fs | NC | NC | 2.2E-06 | 1 | 9143\|0\|0 | 371401\|1\|0 | 2.2E-06 | 0 |
| 19:15179228:G:T | Exon 22 | EGFr 30 | stop_gained | c.3515C>A | p.Ser1172* | NC | NC | 1.1E-06 | 1 | 9143\|0\|0 | 371402\|1\|0 | 1.1E-06 | 0 |
| 19:15178824:TGGGCACA:T | Exon 23 | EGFr 32 | frameshift | c.3829_3835delTGTGCCC | p.Cys1277fs | NC | NC | 1.1E-06 | 1 | 9143\|0\|0 | 371399\|1\|0 | 1.1E-06 | 0 |
| 19:15178092:T:A | NA | No EGFr domain | splice_acceptor | c.3838-2A>T | | NC | NC | 1.1E-06 | 1 | 9143\|0\|0 | 371400\|1\|0 | 1.1E-06 | 0 |
| 19:15177897:CAG:C | Exon 24 | EGFr 34 | frameshift | c.4029_4030delCT | p.Cys1344fs | NC | NC | 3.3E-06 | 3 | 9142\|1\|0 | 371400\|2\|0 | 3.3E-06 | 0 |

| Chrom:pos:ref:alt | Exon | EGFr Domain | Variant Effect | HGVS transcript | HGVS protein | Odds ratio (95% confidence interval) | P value | AAF | AAC | Cases | Controls | UKB 450K | PGR |
|---|---|---|---|---|---|---|---|---|---|---|---|---|---|
| 19:15177835:G:A | Exon 24 | EGFr 34 | stop_gained | c.4093C>T | p.Gln1365* | NC | NC | 1.1E-06 | 1 | 9143\|0\|0 | 371402\|1\|0 | 1.1E-06 | 0 |
| 19:15177752:G:T | Exon 24 | No EGFr domain | stop_gained | c.4176C>A | p.Cys1392* | NC | NC | 1.1E-06 | 1 | 9143\|0\|0 | 371402\|1\|0 | 1.1E-06 | 0 |
| 19:15177739:CG:C | Exon 24 | No EGFr domain | frameshift | c.4188delC | p.Asp1398fs | NC | NC | 1.1E-06 | 1 | 9143\|0\|0 | 371400\|1\|0 | 1.1E-06 | 0 |
| 19:15177713:G:T | Exon 24 | No EGFr domain | stop_gained | c.4215C>A | p.Cys1405* | NC | NC | 1.1E-06 | 1 | 9143\|0\|0 | 371402\|1\|0 | 1.1E-06 | 0 |
| 19:15177655:AG:A | Exon 24 | No EGFr domain | frameshift | c.4272delC | p.Trp1425fs | NC | NC | 1.1E-06 | 1 | 9143\|0\|0 | 371402\|1\|0 | 1.1E-06 | 0 |
| 19:15177637:GCGCCTCGCATTGC:G | Exon 24 | No EGFr domain | frameshift | c.4278_4290delGCAATGCGAGGCG | p.Gln1427fs | NC | NC | 6.6E-06 | 5 | 9143\|0\|0 | 371392\|5\|0 | 6.6E-06 | 0 |
| 19:15177627:C:T | Exon 24 | No EGFr domain | stop_gained | c.4301G>A | p.Trp1434* | NC | NC | 1.1E-06 | 1 | 9143\|0\|0 | 371402\|1\|0 | 1.1E-06 | 0 |
| 19:15174332:TCCGTGTTGCAG:T | Exon 25 | No EGFr domain | frameshift | c.4461_4471delCTGCAACACGG | p.Cys1488fs | NC | NC | 1.1E-06 | 1 | 9143\|0\|0 | 371387\|1\|0 | 1.1E-06 | 0 |
| 19:15174078:C:A | Exon 25 | No EGFr domain | stop_gained | c.4726G>T | p.Glu1576* | NC | NC | 2.2E-06 | 1 | 9143\|0\|0 | 371402\|1\|0 | 2.2E-06 | 0 |
| 19:15170768:A:AT | Exon 26 | No EGFr domain | frameshift | c.4793dupA | p.Asp1598fs | NC | NC | 1.1E-06 | 1 | 9143\|0\|0 | 371402\|1\|0 | 1.1E-06 | 0 |
| 19:15170554:C:T | NA | No EGFr domain | splice_acceptor | c.4892-1G>A | | NC | NC | 2.2E-06 | 1 | 9143\|0\|0 | 371402\|1\|0 | 2.2E-06 | 0 |
| 19:15170162:GC:G | Exon 28 | No EGFr domain | frameshift | c.5122delG | p.Ala1708fs | NC | NC | 1.1E-06 | 1 | 9143\|0\|0 | 371401\|1\|0 | 1.1E-06 | 0 |
| 19:15170139:C:A | Exon 28 | No EGFr domain | stop_gained | c.5146G>T | p.Glu1716* | NC | NC | 1.1E-06 | 1 | 9143\|0\|0 | 371401\|1\|0 | 1.1E-06 | 0 |
| 19:15170138:TC:T | Exon 28 | No EGFr domain | frameshift | c.5146delG | p.Glu1716fs | NC | NC | 1.1E-06 | 1 | 9143\|0\|0 | 371401\|1\|0 | 1.1E-06 | 0 |
| 19:15170112:C:A | Exon 28 | No EGFr domain | stop_gained | c.5173G>T | p.Glu1725* | NC | NC | 1.1E-06 | 1 | 9143\|0\|0 | 371402\|1\|0 | 1.1E-06 | 0 |
| 19:15170103:C:A | Exon 28 | No EGFr domain | stop_gained | c.5182G>T | p.Glu1728* | NC | NC | 3.3E-06 | 2 | 9142\|0\|0 | 371394\|2\|0 | 3.3E-06 | 0 |
| 19:15170085:C:T | Exon 28 | No EGFr domain | splice_donor | c.5199+1G>A | | NC | NC | 1.1E-06 | 1 | 9143\|0\|0 | 371402\|1\|0 | 1.1E-06 | 0 |
| 19:15167297:G:A | Exon 29 | No EGFr domain | stop_gained | c.5314C>T | p.Gln1772* | NC | NC | 1.1E-06 | 1 | 9142\|0\|0 | 371400\|1\|0 | 1.1E-06 | 0 |
| 19:15167248:C:A | Exon 29 | No EGFr domain | splice_donor | c.5362+1G>T | | NC | NC | 1.1E-06 | 1 | 9143\|0\|0 | 371402\|1\|0 | 1.1E-06 | 0 |
| 19:15166049:G:GC | Exon 30 | No EGFr domain | frameshift | c.5404dupG | p.Ala1802fs | NC | NC | 2.2E-06 | 2 | 9143\|0\|0 | 371400\|2\|0 | 2.2E-06 | 0 |
| 19:15165893:G:GC | Exon 30 | No EGFr domain | frameshift | c.5560dupG | p.Ala1854fs | NC | NC | 1.1E-06 | 1 | 9143\|0\|0 | 371402\|1\|0 | 1.1E-06 | 0 |
| 19:15165785:A:C | NA | No EGFr domain | splice_donor | c.5667+2T>G | | NC | NC | 1.1E-06 | 1 | 9143\|0\|0 | 371402\|1\|0 | 1.1E-06 | 0 |
| 19:15165506:G:A | Exon 31 | No EGFr domain | stop_gained | c.5677C>T | p.Arg1893* | NC | NC | 1.1E-06 | 1 | 9143\|0\|0 | 371402\|1\|0 | 1.1E-06 | 2.6E-05 |
| 19:15165492:TG:T | Exon 31 | No EGFr domain | frameshift | c.5690delC | p.Thr1897fs | NC | NC | 1.1E-06 | 1 | 9143\|0\|0 | 371401\|1\|0 | 1.1E-06 | 0 |
| 19:15162525:G:GTTGT | Exon 32 | No EGFr domain | frameshift | c.5849_5852dupACAA | p.Asn1951fs | NC | NC | 1.1E-06 | 1 | 9143\|0\|0 | 371394\|1\|0 | 1.1E-06 | 0 |
| 19:15162464:C:T | Exon 32 | No EGFr domain | splice_donor | c.5913+1G>A | | NC | NC | 1.1E-06 | 1 | 9143\|0\|0 | 371402\|1\|0 | 1.1E-06 | 0 |
| 19:15161701:A:AG | Exon 33 | No EGFr domain | frameshift | c.5926dupC | p.Leu1976fs | 0.36 (0, 447.32) | 0.78 | 9.2E-06 | 7 | 9143\|0\|0 | 371393\|7\|0 | 8.8E-06 | 0 |

| Chrom:pos:ref:alt | Exon | EGFr Domain | Variant Effect | HGVS transcript | HGVS protein | Odds ratio (95% confidence interval) | P value | AAF | AAC | Cases | Controls | UKB 450K | PGR |
|---|---|---|---|---|---|---|---|---|---|---|---|---|---|
| 19:15161512:AG:A | Exon 33 | No EGFr domain | frameshift | c.6115delC | p.Leu2039fs | NC | NC | 1.1E-06 | 1 | 9143\|0\|0 | 371402\|1\|0 | 1.1E-06 | 0 |
| 19:15161247:A:C | Exon 33 | No EGFr domain | stop_gained | c.6381T>G | p.Tyr2127* | NC | NC | 1.1E-06 | 1 | 9143\|0\|0 | 371401\|1\|0 | 1.1E-06 | 0 |
| 19:15161177:G:A | Exon 33 | No EGFr domain | stop_gained | c.6451C>T | p.Gln2151* | NC | NC | 1.1E-06 | 1 | 9143\|0\|0 | 371402\|1\|0 | 1.1E-06 | 0 |
| 19:15161112:C:T | Exon 33 | No EGFr domain | stop_gained | c.6516G>A | p.Trp2172* | NC | NC | 1.1E-06 | 1 | 9143\|0\|0 | 371402\|1\|0 | 1.1E-06 | 0 |
| 19:15160951:GC:G | Exon 33 | No EGFr domain | frameshift | c.6676delG | p.Ala2226fs | NC | NC | 1.1E-06 | 1 | 9142\|0\|0 | 371401\|1\|0 | 1.1E-06 | 0 |
| 19:15160760:G:A | Exon 33 | No EGFr domain | stop_gained | c.6868C>T | p.Gln2290* | NC | NC | 2.2E-06 | 1 | 9143\|0\|0 | 371402\|1\|0 | 2.2E-06 | 0 |
| 19:15160718:G:A | Exon 33 | No EGFr domain | stop_gained | c.6910C>T | p.Gln2304* | NC | NC | 1.1E-06 | 1 | 9143\|0\|0 | 371402\|1\|0 | 1.1E-06 | 0 |
| 19:15160694:C:A | Exon 33 | No EGFr domain | stop_gained | c.6934G>T | p.Glu2312* | NC | NC | 1.1E-06 | 1 | 9143\|0\|0 | 371401\|1\|0 | 1.1E-06 | 0 |
| 19:15160664:A:C | Exon 33 | No EGFr domain | stop_lost | c.6964T>G | p.Ter2322Glyext*? | NC | NC | 1.1E-06 | 1 | 9143\|0\|0 | 371402\|1\|0 | 1.1E-06 | 0 |

1. To test the hypothesis that loss of function (LoF) variants in *NOTCH3* do not contribute to stroke risk in the UKB European population, individual LoF variants were combined into a burden test. Shown is a table of all LoF variants in *NOTCH3* that were observed in UKB Europeans, including single variant association summary statistics for variants with alternate allele count (AAC) $\geq$ 5. From left-to-right is variant location (in chromosome:position:reference:alternate format with GRCh38 coordinates); the exon in the canonical ENST00000263388 transcript; specific EGFr domain number; impact on ENST00000263388 transcript; impact on the protein sequence; single variant analysis effect size (odds ratio with 95% confidence interval in parenthesis); single variant analysis p value; alternate variant allele frequency; alternate variant allele count; number of cases (organized by Ref/Ref homozygotes | Ref/Alt heterozygotes | Alt/Alt homozygotes); number of controls; alternate allele frequency in the full UKB 450 K cohort across ancestries; alternate allele frequency for the variant in 75 K PGR cohort. "NC" stands for "Not Calculated", meaning that the case and control counts of the variant was insufficient to calculate a single variant analysis p value and effect size. "NA" stands for "not applicable". Table is ordered from start to end of transcript.

**Supplementary Figures**

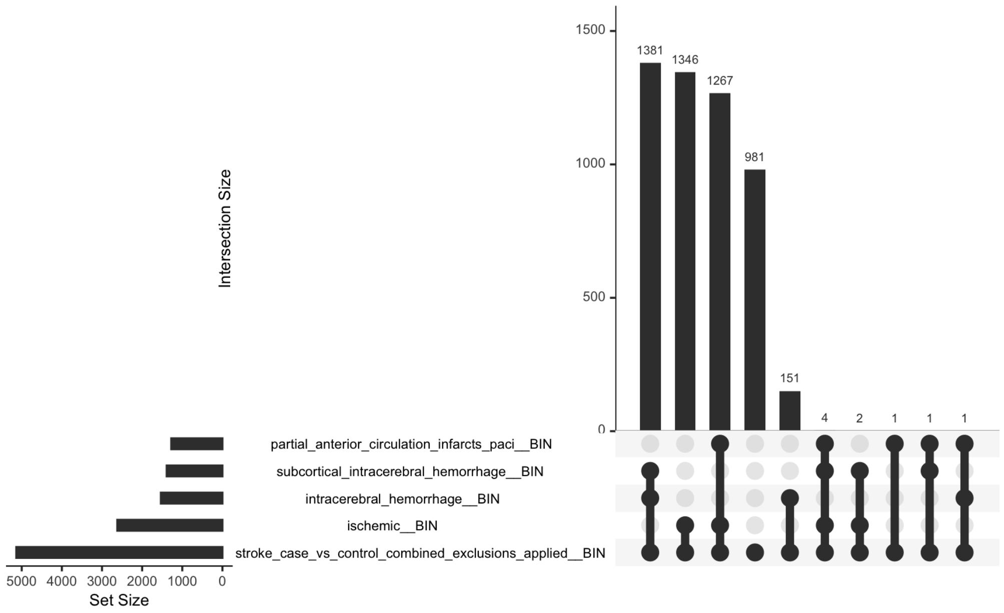


**Supplementary Figure 1. Overlap between Cases across 5 Stroke Phenotypes in PGR Discovery Cohort.** ExWAS was conducted for 5 stroke phenotypes, including the all-inclusive "stroke" (n = 5,135 cases) and 4 sub-phenotypes whose cases are subsets of the "stroke" cases". While all sub-phenotype cases are subsets of the "stroke" meta-phenotype, there was some overlap between cases for the 4 sub-phenotypes. To visualize the extent of overlap between stroke and 4 sub-phenotypes, a UpSetR plot was generated. At the bottom left is a bar plot illustrating the sample size of each of the 5 stroke case phenotypes ordered from smallest to largest sample size (stroke, ischemic, intracerebral hemorrhage, subcortical intracerebral hemorrhage, PACI). In the bottom right are horizontal lines, where each line represents one intersecting set between two or more sets, with a filled black circle showing presence of the phenotype (row) in the overlapping set and an unfilled grey circle showing non-presence in the intersection set. On top of each intersection line is a bar plot of the intersection size, ordered from left to right from largest to smallest.

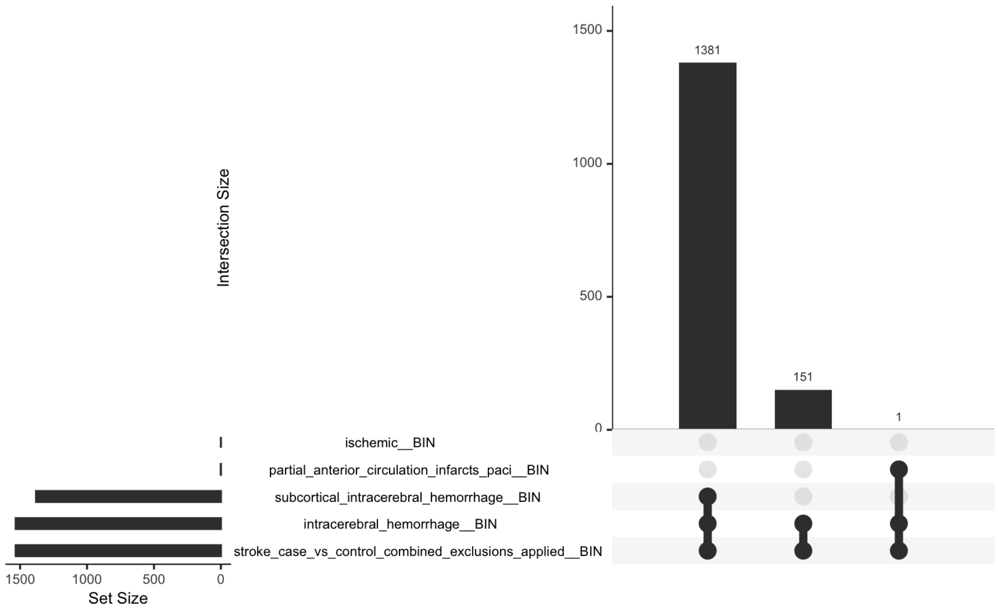


**Supplementary Figure 2. Overlap between Subcortical Stroke Cases across 5 Stroke Sub-Phenotypes in PGR Discovery Cohort.** To visualize the extent of overlap between subcortical stroke cases (n = 1,388) and 4 other stroke sub-phenotypes (including the meta-phenotype “stroke” of which all 4 sub-phenotypes, including subcortical stroke, are subsets of), an UpSetR plot was generated. At the bottom left is a bar plot illustrating the sample size of each of the 5 stroke case sub-phenotypes ordered from smallest to largest overlap sample size. In the bottom right are horizontal lines, where each line represents one intersection set between two or more sets, with a filled black circle showing presence of the phenotype (row) in the overlapping set and an unfilled grey circle showing non-presence in the intersection set. On top of each intersection line is a bar plot of the intersection size, ordered from left to right from largest to smallest.

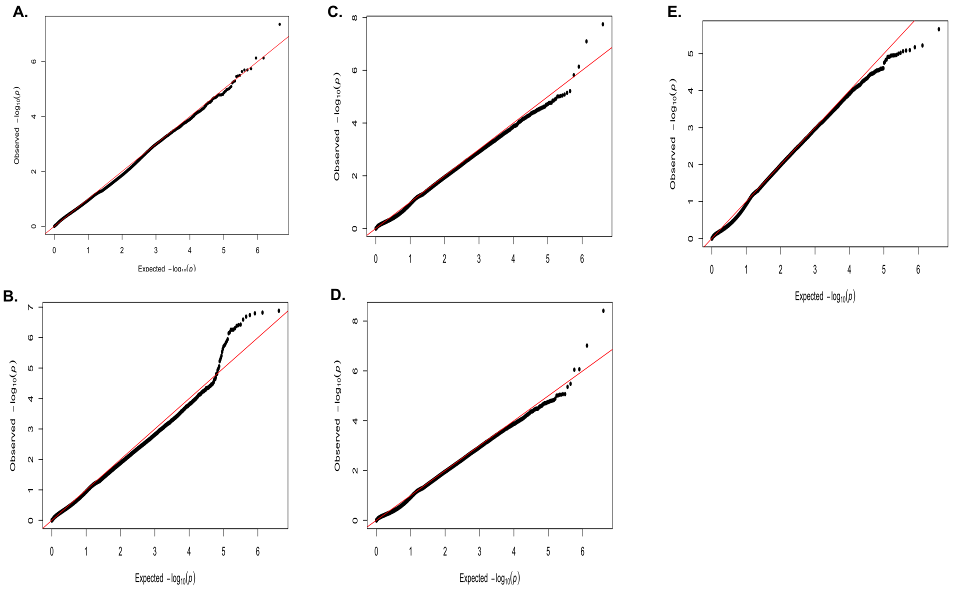


**Supplementary Figure 3. QQ Plots of 5 Stroke ExWASes in PGR Discovery Cohort.** Plot compares observed (y-axis) to expected (x-axis) ExWAS likelihood ratio test -$\log_{10}$ p-values calculated using REGENIE for n= 5,135 stroke cases and n = 26,602 controls. **A**. Stroke; **B.** Ischemic; **C**. Hemorrhagic; **D**. Subcortical, **E**. PACI.

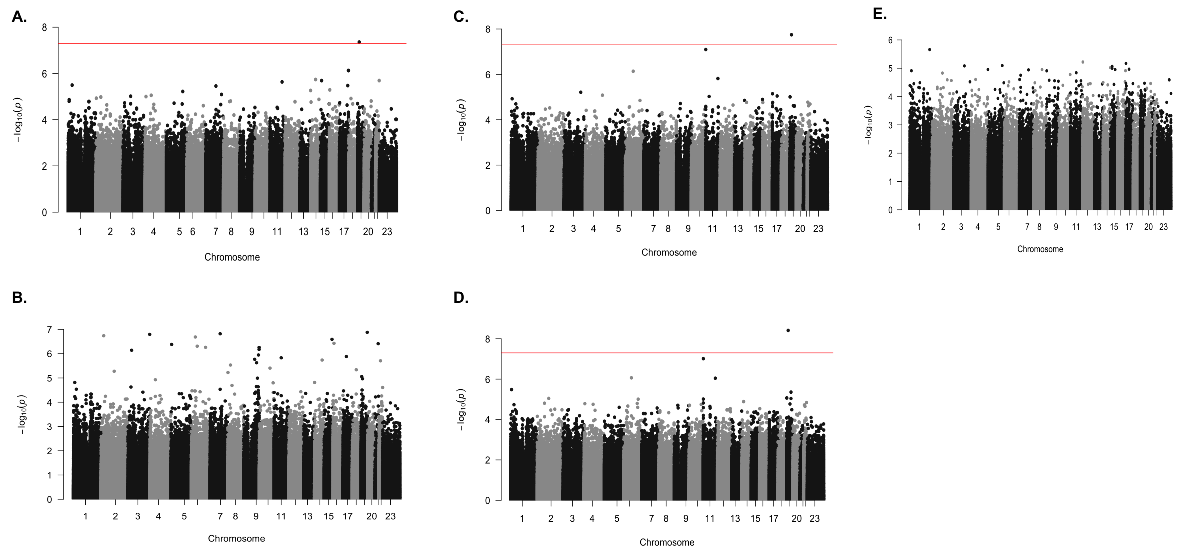


**Supplementary Figure 4. Manhattan Plots of 5 Stroke ExWASes in PGR Discovery Cohort.** Plot shows the ExWAS likelihood ratio test -log$_{10}$ p values calculated using REGENIE for n= 5,135 stroke cases and n = 26,602 controls (y-axis) versus the position in the genome (x-axis). Alternating colors by chromosome from 1 to 23 (ChrX), with a dotted horizontal line showing the genome-wide significance threshold of 5.0x10$^{-8}$ for ExWASes where at least one variant had a -log$_{10}$ p value above threshold. **A**. Stroke; **B.** Ischemic; **C**. Hemorrhagic; **D**. Subcortical, **E**. PACI.

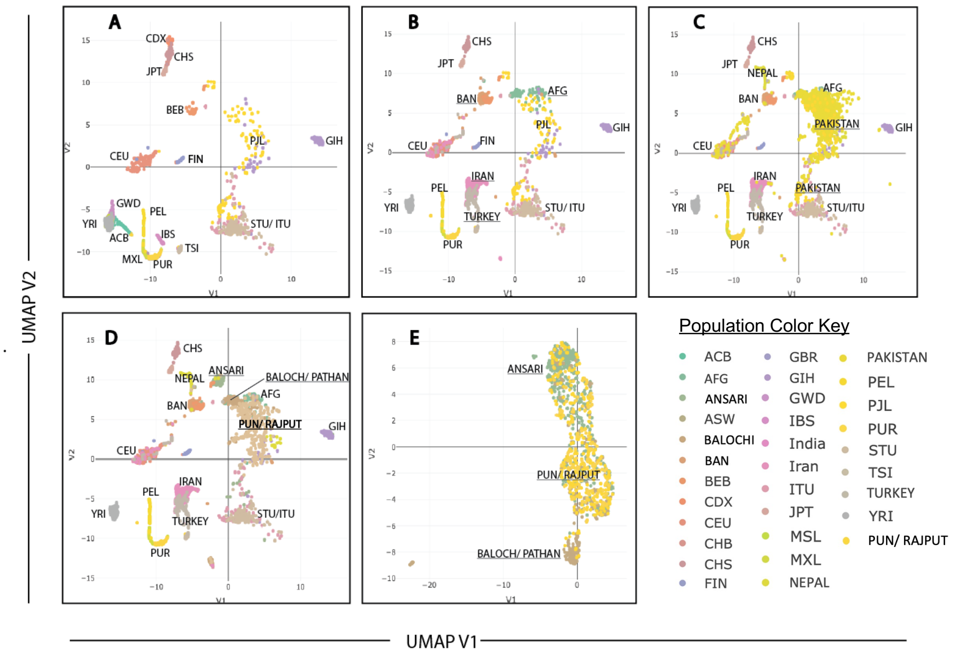


**Supplementary Figure 5. Population Structure of Pakistan** This plot was created using first seven principal components (PCs) computed using genome-wide SNV data from PGR, 1000G and UKB. UMAP is a dimensional reduction technique that can be used to view complex data in two-dimensional space. In this plot the x-axis represents the first dimension and y-axis the second dimension of UMAP. UMAP plot for CNCD data formed three major clusters that were enriched with individuals identifying as Ansari, Punjabi/Rajput and Balochi/Pathan ancestries. **A.** 1000 Genomes Populations. **B.** Same as panel A, with addition of UKB South Asian (SAS) and West Asian (WAS) Populations with n > 100. **C.** Same as panel B, with addition of n = 900 from PGR cohort. **D.** Same as panel C, with Pakistani ancestry clusters labeled (n = 300 Baloch/Pathan, n = 300 Punjabi/Rajput, n = 300 Ansari). **E.** Focus on the PGR to show the major ancestry clusters. Each color on plot represents an individual ancestry and has been labeled accordingly as described in the Population Color Key. The abbreviations of corresponding ancestries as follow: 1000 Genomes Project African Caribbean in Barbados (ACB), 1000 Genomes Project Bengali in Bangladesh (BEB), 1000 Genomes Project Chinese Dai in Xishuangbanna, China (CDX), 1000 Genomes Project Han Chinese South (CHS), 1000 Genomes Project Finnish in Finland (FIN), 1000 Genomes Project Gujarati Indians in Houston, Texas, USA (GIH), 1000 Genomes Project Gambian in Western Division – Mandinka (GWD), 1000 Genomes Project Iberian Populations in Spain (IBS), 1000 Genomes Project Indian Telugu in the U.K. (ITU), 1000 Genomes Project Japanese in Tokyo, Japan (JPT), 1000 Genomes Project Mexican Ancestry in Los Angeles CA USA (MXL), 1000 Genomes Project Peruvian in Lima Peru (PEL), 1000 Genomes Project Puerto Rican in Puerto Rico (PUR), 1000 Genomes Project Punjabi in Lahore, Pakistan (PJL), 1000 Genomes Project Sri Lankan Tamil in the UK (STU]), 1000 Genomes Project Toscani in Italia (TSI), 1000 Genomes Project Yoruba in Ibadan, Nigeria (YRI), UKB Country of birth Afghanistan (AFG) (n = 112), UKB Country of birth Bangladesh (BAN) (n = 246), UKB Country of birth Iran (IRAN) (n = 540), UKB Country of birth Nepal (NEPAL) (n = 161), UKB Country of birth Pakistan (PAKISTAN) (n = 1439), UKB Country of birth Turkey (TURKEY) (n = 182), CNCD Baloch/Pathan (BALOCH/PATHAN) (n = 3,068), CNCD Punjabi/Rajput (PUN/RAJPUT) (n = 20,907), CNCD (ANSARI) (n = 10,402).

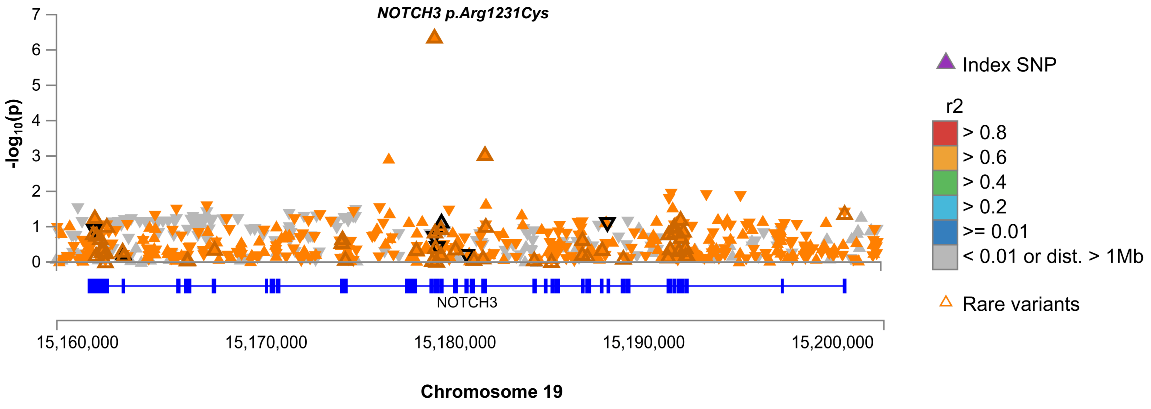


**Supplementary Figure 6. Evidence for Pathological Changes in Brain of p.Arg1231Cys Carriers.** LocusZoom plot of *NOTCH3* for white matter hyperintensity (WMH) in n = 35,344 UKB participants. WMH was one of several brain MRI phenotypes with linear regression test p values below $1.0 \times 10^{-5}$. Shown is a plot of the $-\log_{10}$ p values (y axis) for all *NOTCH3* exome variants, ordered by position (x axis), with p.Arg1231Cys labeled. Variants represented as triangles color-coded by LD with the index SNP.

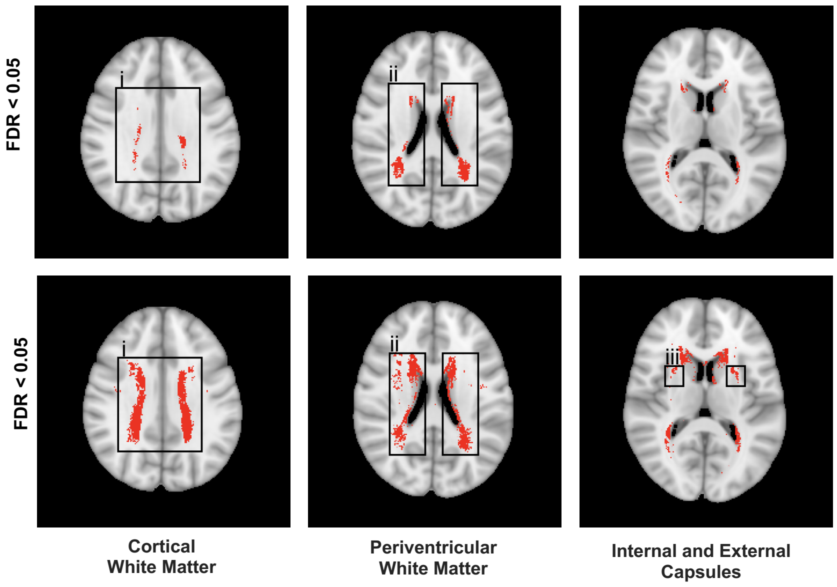


**Supplementary Figure 7. Voxel-wide Heatmap of White Matter Hyperintensity Signal in *NOTCH3* Cys-Altering Mutation Carriers Compared to Controls from UK Biobank**. Shown is the voxel-wide heatmap of white matter hyperintensity (WMH) signal with voxel significant differences calculated by logistic regression in n = 35,344 UKB participants with brain MRI data between (*top*) n = 20 p.Arg1231Cys carriers vs. controls and (*bottom*) n = 86 EGFr Cys-altering variant cariers vs. controls shown in red. (top) Signals observed most strongly in (i) the centrum semiovale (cortical white matter) and (ii) the periventricular white matter, to lesser degree in the internal and external capsule white matter. (bottom) Signals observed appreciably in (i) the centrum semiovale (cortical white matter), (ii) the periventricular white matter, and (iii) the external capsules bilaterally.
